# Disentangling independent and mediated causal relationships between blood metabolites, cognitive factors, and Alzheimer’s Disease

**DOI:** 10.1101/2021.02.12.21251409

**Authors:** Jodie Lord, Rebbeca Green, Shing Wan Choi, Christopher Hübel, Dag Aarsland, Latha Velayudhan, Pak Sham, Cristina Legido-Quigley, Marcus Richards, Richard Dobson, Petra Proitsi, on behalf of the Alzheimer’s Disease Neuroimaging initiative, the GERAD1 Consortium, AddNeuroMed

## Abstract

**Background:** Education and cognition demonstrate consistent inverse associations with Alzheimer’s Disease (AD). The biological underpinnings, however, remain unclear. Blood metabolites can reflect the endpoint of biological processes and are accessible and malleable. Identifying metabolites with aetiological relevance to AD and disentangling how these relate to cognitive factors along the AD causal pathway could, therefore, offer unique insights into underlying causal mechanisms.

**Methods:** Using data from the largest metabolomics genome-wide association study (*N*≈24,925) and three independent AD cohorts (*N*=4,725), cross-trait polygenic scores were generated and meta-analyzed. Metabolites genetically associated with AD were taken forward for causal analyses. Bidirectional two-sample Mendelian randomization (MR) interrogated univariable causal relationships between (i) metabolites and AD, (ii) metabolites, education and cognition (iii) education, cognition and AD, and (iv) education and cognition. Mediating relationships were computed using multivariable MR.

**Results:** Thirty-four metabolites were genetically associated with AD at *p*<0.05. Of these, glutamine and free cholesterol in extra-large high-density lipoproteins (XL.HDL.FC) demonstrated a protective causal effect (Glutamine: 95% CI=0.70-0.92; XL.HDL.FC: 95% CI=0.75-0.92). An AD-protective effect was also observed for education (95% CI=0.61-0.85) and cognition (95% CI=0.60-0.89), with bidirectional mediation evident. Cognition as a mediator of the education-AD relationship was stronger than vice-versa, however. No evidence of mediation via any metabolite was found.

**Conclusions:** Glutamine and XL.HDL.FC show protective causal effects on AD. Education and cognition also demonstrate protection, though education’s effect is almost entirely mediated by cognition. These insights provide key pieces of the AD causal puzzle, important for informing future multi-modal work and progressing towards effective intervention strategies.

## Introduction

Late-onset Alzheimer’s Disease (AD) presents a huge burden to society, affecting over 47 million individuals worldwide (1). Factors such as educational attainment (EA) and cognition demonstrate protective associations with AD (2–4), and these associations may indeed be causal (5–9). The aetiological mechanisms underlying these relationships, however, remain unclear. Understanding the biological basis through which cognitive factors may exert their protective effect on AD, as well as establishing direct markers of disease pathogenesis more generally, could therefore hold special value in advancing treatment and prevention strategies.

Blood metabolites - small molecular compounds such as lipids and amino acids - have the potential to provide vital clues to how education and cognition may indirectly influence AD risk. Concentrations of these analytes represent the crosstalk between genomic encoding together with influences from the surrounding environment (10, 11). Covariance of metabolites with AD relevant risk-factors could therefore be indicative of mediated relationships along the same causal pathway. Further, as metabolites are quantifiable via a simple blood-test and many are of a plausible size to cross the blood-brain-barrier (12), they represent promising candidates as direct targets for treatment intervention. While research has indeed implicated several metabolites, particularly lipids, in AD and cognitive processing (13–16), the weight of evidence derives from observational studies. These remain problematic with respect to informing intervention strategies, as uncaptured confounding and reverse causation risk incorrect causal inferences. Moreover, associative studies allow little opportunity to understand specific pathways into disease endpoints, and as such, mediating relationships have been little explored.

Mendelian Randomization (MR) presents a statistical methodology akin to a randomized control trial which allows researchers to investigate putative causal relationships through use of genetic variants as randomizing instruments (17). Multivariable approaches allow for the addition of covariables which may influence the exposure-outcome relationship. When independent instruments are utilised, this then allows for mediating relationships to be interrogated (18, 19). In a previous study, we utilised MR to investigate causal relationships between nineteen candidate metabolites and AD. A protective effect of extra-large high-density lipoproteins (XL.HDLs), and a risk increasing effect of glycoprotein acetyls (GP) was found (20). Whilst offering preliminary insight, candidate metabolites were restricted to those previously associated with midlife cognition (21), rather than AD specifically, neglecting other causal candidates which may be of relevance. Mediating relationships were also not explored, which if demonstrated, could offer intermediate sources of intervention and provide a richer understanding of the aetiological drivers behind relationships observed.

This study sought to extend our previous findings, this time using cross-trait polygenic risk scoring (PRS) to prioritise AD-specific candidate metabolites; selecting only those most genetically predictive of AD diagnosis. To disentangle causality between these candidate metabolites, AD, and AD-relevant cognitive factors, univariable MR was then utilised to interrogate bidirectional causal relationships. Finally, we introduced a multivariable framework to disentangle independent verses mediating mechanisms through which causal associations arise. In this way, we distinguished the extent to which causal relationships are mutually independent, or whether they reflect a chain of interdependent causal events along the same AD pathway. If so, this could offer insights into direct and indirect sources of AD intervention.

## Methods and Materials

### Data sources

#### Blood Metabolites

Summary statistics from the largest metabolomics genome-wide association study (GWAS) (N≈24,925) (22) were utilised for the generation of PRS and for MR analyses. Here, 123 blood metabolites were quantified using nuclear magnetic resonance (NMR) spectroscopy, including lipid subfractions, amino acids, and glycolysis precursors (Supplementary Dataset S1). All samples were of European ancestry. Ages ranged from 31-61 years (mean=44.6 yrs), and all datasets were adjusted for age, sex, time of last meal, and 10 principal components (PCs).

##### Alzheimer’s Disease

Genotype data across three AD cohorts were obtained for use as the target phenotype within PRS analyses; (i) The Genetic and Environmental Risk in Alzheimer’s Disease (GERAD1) consortium (https://gtr.ukri.org/project/B6C58A7C-3C3E-41CB-AF10-16DB59962C9E/), (ii) The Alzheimer’s Disease Neuroimaging Initiative (ADNI) ((23), http://adni.loni.usc.edu/), and (iii) The AddNeuroMed & Dementia Care Register (ANM) (24, 25). Cohort details can be found within Supplementary Information S3.

For MR, summary data from the largest GWAS of clinically diagnosed late-onset AD was used (26). This study consisted of three stages; (i) a discovery phase (*N*=63,926), (ii) a replication phase (*N*=18,845), and (iii) a post replication phase (*N*=11,666). Datasets were adjusted for sex, age, and PCs and yielded twenty-five genome-wide significant hits (GSH).

#### Educational attainment

GWAS data from Lee et al. (27) represented education (*GSH=*1,271, *N*=1.1million); defined as the highest education level obtained by age thirty. Age, sex, and PCs were adjusted for, and individuals were of European ancestry.

#### Cognition

Cognition was measured using meta-analysed GWAS data from Savage et al. (28) (*GSH*=205, *N*=269,867). Different measures of cognition were mapped to a common latent “*g”* factor, representing multiple dimensions of cognitive functioning. Cohorts ranged from children (min: age 5 yrs) to older adults (max: age 98 yrs), with older adults screened for cognitive decline to exclude possible dementia. All cohorts were adjusted for age, sex and PCs.

### Polygenic Score preparation

Quality control (QC) was conducted across AD cohorts separately, each following the same pipeline (Fig.1) using a combination of PLINK 1.90 (29) and R 3.6.1. PCA was performed using EIGENSOFT 6.1.4. (https://www.hsph.harvard.edu/alkes-price/software/), and genotyped data were imputed via the Sanger Imputation Service (31). Individuals of non-white European ancestry, or whose most recent diagnosis was MCI or non-AD dementia were excluded. Minimum sample ages within ADNI, ANM, and GERAD1 cohorts were 54, 53, and 43 years respectively. As AD has a long symptom-free prodromal phase, cases and controls were age matched to a conservative minimum of 70 years to avoid contamination of pre-clinical cases within the control group. SNPs within 750kb of the ApoE genomic region were removed. Following imputation, separately typed platforms within the ADNI cohort were merged, resulting in three AD datasets taken forward for PRS (1xADNI, 1xGERAD, 1xANM).

**Figure 1.**
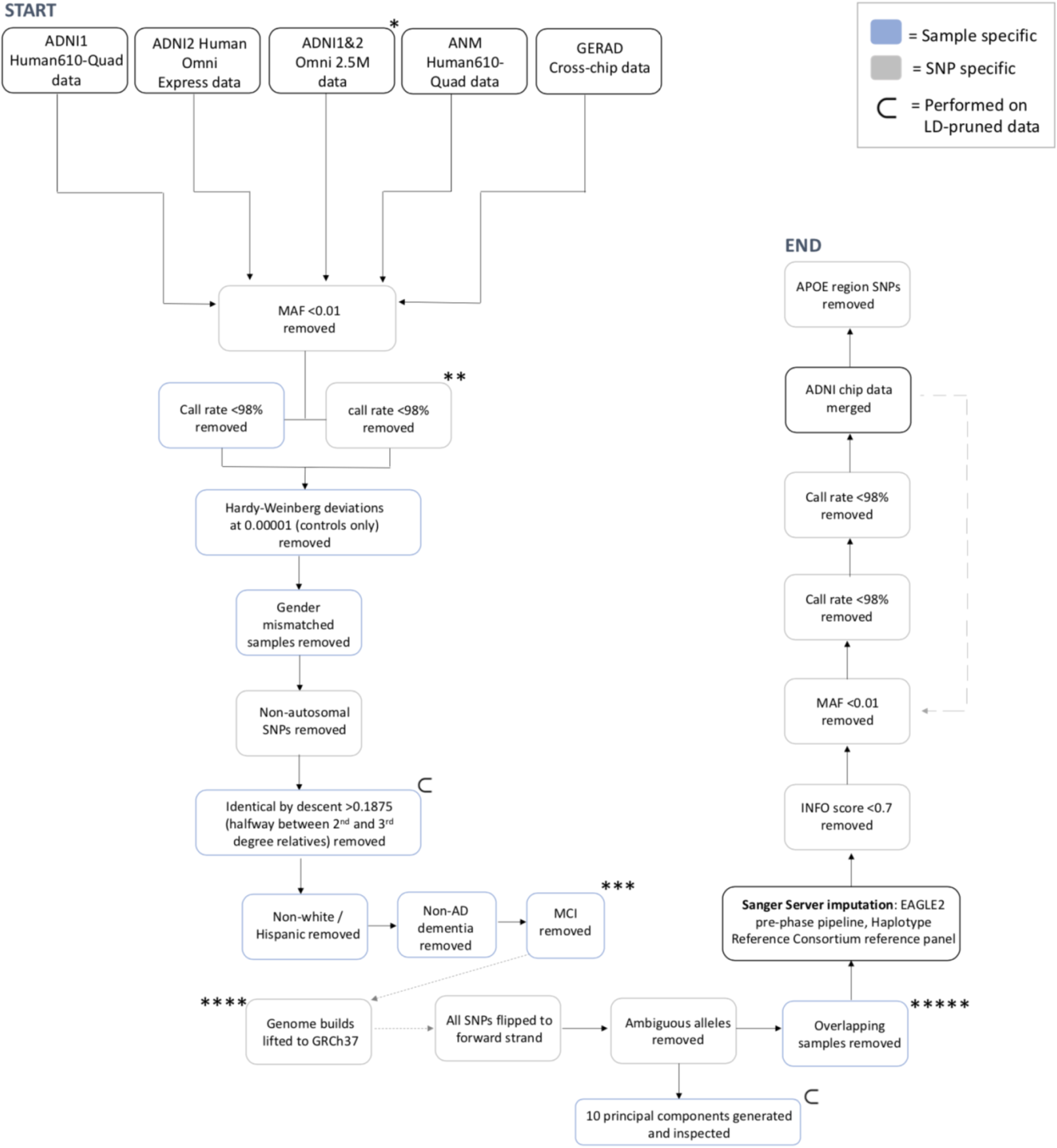
Flow chart overviewing quality control pipelines applied to all AD genotype datasets (separately) prior to PRS analyses. * The illumina omni2.5 microarray chip includes samples across ADNI1 and 2, whole genome sequenced at high coverage, and subsequently genotyped on the high coverage illumina chip. ** Missingness for both SNPs and samples were inspected iteratively, from 90-98%, iterating between SNPs and samples in steps of 1%. *** Latest diagnosis was used to classify samples into cases and controls. Late stage MCI, with MCI due to probable AD, and clinician confidence score of 3-4 (indicating high confidence) remained in analyses as cases. **** Required for Human610-Quad platform only. ***** For overlaps between ADN1 chip data (Human610-Quad) and Omni 2.5M, duplicates in Omni 2.5M removed and Human610-Quad retained. For overlaps between ADNI1 chip data (HumanOmniExpress) and Omni 2.5M, duplicates in HumanOmniExpress removed and Omni 2.5M retained. No overlaps observed between Human610-Quad and HumanOmniExpress.

To quantify signal power within metabolite datasets, estimates of SNP-based heritability (h^2^_SNP_) were computed using LD score regression (30). h^2^*Z* scores (h^2^*Z* = h^2^_SNP_/*se*) were subsequently computed (31). Datasets with h^2^*Z*<2 or with out-of-range h^2^_SNP_ (0<>1) were excluded.

### Mendelian randomization preparation

To maximise instrumental variable (IV) validity, SNPs were selected only if they (i) were associated with their exposure at genome-wide significance or below (*p*<5×10^- 8^), (ii) demonstrated a computed F>10 (32), and (iii) were not within 750kb of the ApoE genomic region due to known pleiotropy (33, 34). To ensure instrument independence, SNPs were clumped using an *r*^2^ threshold of 0.001 for AD, EA, and intelligence (Supplementary Dataset S2-S4). For metabolites, instruments were selected from a set of pre-curated Kettunen (22) metabolite quantitative trait loci (mQTLs) available within MRBase; no additional clumping was required for these (Supplementary Dataset S5). Finally, all datasets were harmonised and any SNPs with non-inferable palindromic SNPs (minor allele frequency (MAF)>0.40) or with MAF<0.01 were excluded. Metabolite MAFs were used to infer AD allele frequencies, as these were unavailable. All data extraction, pre-processing and analyses were performed within R.3.6.1. using the MRBase package (v.0.4.25) (35). A Document further detailing scope in-line with MR reporting guidelines (36) is provided in Supplementary Information S2.

### Primary statistical analyses

#### Genetic associations using polygenic scoring

PRS models were generated using PRSice-2 (37). Each metabolite was set as the model generating “base” dataset which was then used to predict status within each AD dataset separately. For each metabolite, models were generated across ten pre-defined p-value threshold (*P_T_*): 5×10^-08^, 1×10^-05^, 1×10^-04^, 0.001, 0.01, 0.05, 0.1, 0.2, 0.5, 1. SNPs with *p*-value<*P_T_* were weighted by their effect size, aggregated within the PRS of the corresponding model and regressed on AD genotypes separately. SNPs with r^2^>0.1 were clumped to account for linkage disequilibrium (LD). Sex, age, and seven PCs were adjusted for and all models were standardised (*mean*=0, *sd*=1).

##### PRS meta-analyses

PRSs for each metabolite at every *P_T_* were meta-analysed across the three AD datasets using REML within the metafor R package (38). For each metabolite, the ten meta-analysed results (one per *P_T_*) were ranked by their *p*-value to obtain the metabolite most significantly predictive of AD status (Supplementary Fig S1). Pseudo-R2s were backcomputed using the meta-analysed regression coefficients (Supplementary Information S4). I^2^ and Cochran’s-Q (*Q*) were used to assess cross-study heterogeneity, indicated by I2>0.5 and *Q-p*-value<0.05. To reflect multiple testing, an adjusted alpha of α=0.0002 was computed using the navigome_independent_tests package in python 3.7 (https://github.com/hagax8/independent_tests<u>. Supplementary Information S5</u>). Metabolites genetically associated with AD status were selected for MR analyses.

#### Bidirectional causal analyses

Inverse variance weighted (IVW) two-sample univariable MR (UVMR) was subsequently conducted; a method analogous to a fixed-effect meta-analysis, with the intercept constrained to zero to reflect MR’s exclusion restriction assumption (34). Two-sample reflects the use of two independent datasets – (i) exposure and (ii) outcome. To compute the total causal effect of each metabolite on AD, metabolite_1…*j*_ was set as the exposure *(x)* in turn, and AD the outcome *(y)*. To interrogate reverse causation, *x* and *y* were then reversed. This bidirectional procedure was repeated for metabolites and education, metabolites and cognition, education and AD, cognition and AD, and education and cognition. While the temporal order of education (highest grade at 30 years) and AD make reverse causation implausible, bidirectional analyses were undertaken as a negative control. To align orders of magnitude, all results were computed in standard deviation (SD) units.

#### Causal mediation

Multivariable MR (MVMR) was employed to interrogate mediation. This followed the same framework as UVMR but with the addition of a second “mediating” variable (*m*), allowing for a causal estimate of *x* on *y* whilst holding *m* constant. A difference between the total estimate (*c*) derived from UVMR, and the direct estimate (*c’*) derived from MVMR, was used to signify a mediating effect of *m* on the *x-y* relationship (Fig. 2) (19). To ensure direct estimates were not biased by confounding of *m*, datasets were cross-checked to ensure no IV overlap between *x* and *m* datasets (Supplementary Datasets S2-S5). In this way, any *m* to *x* confounding would not enter into *c’* estimate. Thus any *c*-*c’* difference could be concluded a consequence of downstream mediation (18).

**Figure 2.**
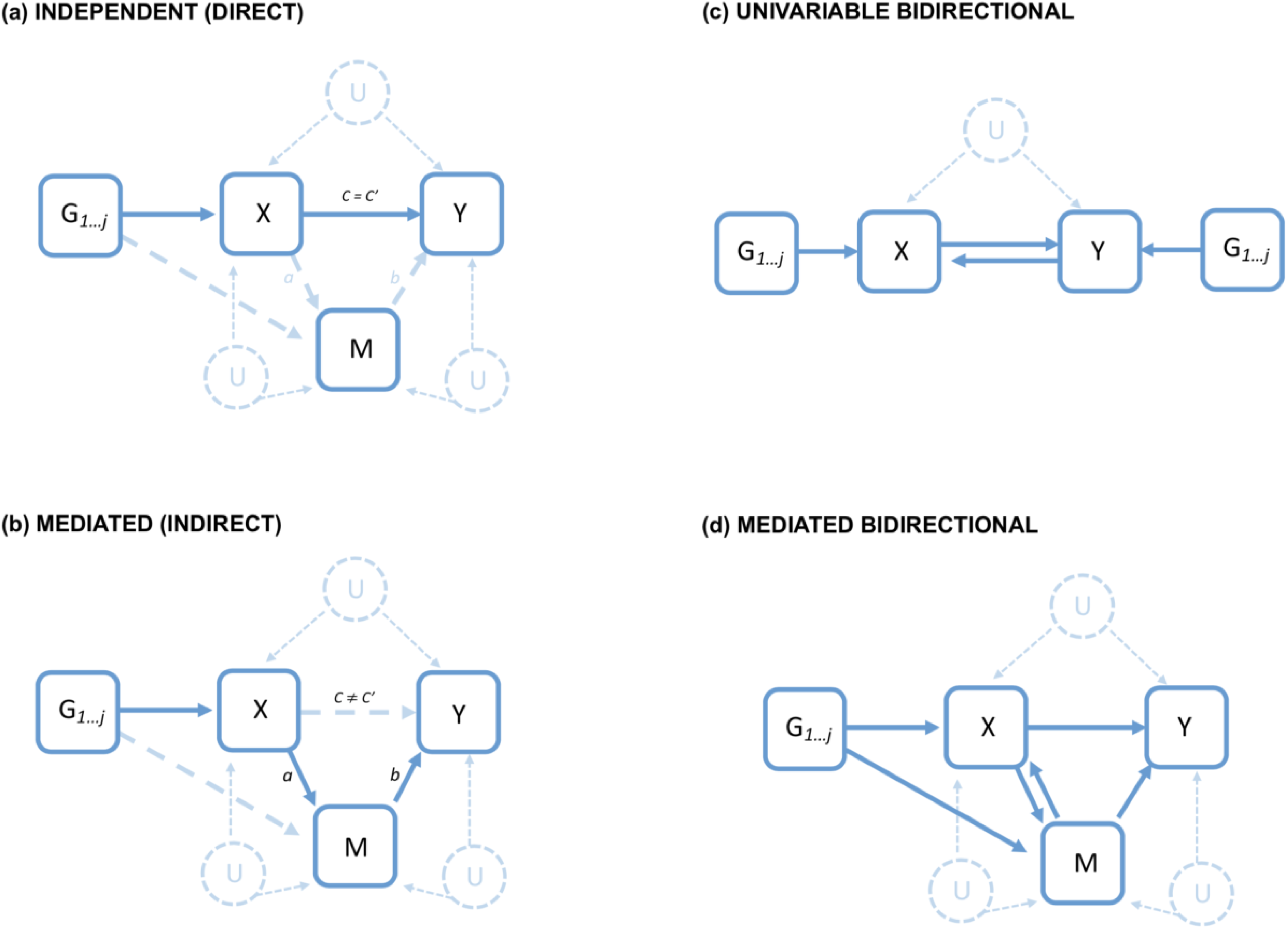
Diagrammatic illustration of causal paths identifiable using univariable and multivariable Mendelian randomization, assuming MR assumptions are satisfied. *G1…j*=Genetic instruments used as proxy for random treatment assignment. X=exposure of interest. Y=outcome of interest. M=mediator of interest. U=potential unmeasured confounding. C=total causal estimates (univariable estimate when mediator unaccounted for). C’=direct causal estimate (multivariable estimate when holding the mediator constant). a-path (X to M) * b-path (M to Y)=mediated path of X to Y via M. Solid arrows represent observed causal effects in the direction depicted by the arrow-head. Dashed arrows represent potential directed relationships not fully observed. For (a) dashed arrows from X to M and M to Y represent the possibility of either an active a-path or an active b-path, but no significant effect of both. This would be expected if there was no difference between univariable (C) or multivariable (C’) estimates of X. For (b) a dashed arrow from X to M represents an inactivation of the C’ path when M is introduced via multivariable models. C’ may be partially inactivated (reduced magnitude of C’ relative to C but a significant relationship maintained) or fully inactivated (complete loss of causal signal in C’ estimate) by the presence of M. (c) and (d) depict bidirectional relationships. For (c) a significant causal effect is observed when switching X and Y as the exposure of interest. For (d) a significant causal effect is observed when switching X and M as the mediator of interest. All models allow for unmeasured confounding between X,M and Y. Core MR assumptions are assumed to hold in all models ((i) no G_1…j_ to X confounding, (ii) no direct G_1…j_ to Y relationship, (iii) a robust relationship between G_1…j_ and X).

For variables to be candidate mediators, evidence of an association from *x* to *m* (when *m=y*), and from *m* to *y* (when *m=x*) was first required within UVMR. A total causal effect of *x* to *y* was not, however, necessary due to potential suppression through *m* (39). Univariable results were therefore used to inform variable selection for MVMR. Any exposure with a univariable effect on *y* but no univariable effect on a potential *m* (when *m*=y) was automatically deemed to be acting independently of *m* and not selected for MVMR. As with UVMR, all variables were measured in SD units.

### Sensitivity analyses

For both UVMR and MVMR analyses, causal effects were re-estimated using MR-Egger; a conservative method comparable to IVW with the intercept constraint removed. An intercept significantly deviating from 0 is indicative of pleiotropy, as are large discrepancies between Egger and IVW estimates (40). For UVMR, several additional sensitivity were undertaken, including; (i) weighted-median MR which, like Egger, re-computes causal estimates with assumptions regarding pleiotropy relaxed (34); (ii) leave-one-out to investigate the presence of influential points; (iii) Cochran’s-*Q* to examine instrument heterogeneity (36); and (iv) Bayesian model averaging (MR-BMA) to re-assess metabolite-AD causal estimates whilst accounting for known correlation amongst metabolites (41). Further information can be found within Supplementary Information S6.

### Post hoc analyses

#### Validity of glutamine as a causal analyte

Of metabolites showing evidence of a causal association with AD in primary analyses, one – glutamine – was observed to have an IV outlier. The removal of this resulted in a non-significant result (see Results). To further explore the validity of glutamine as a metabolite on the causal pathway to AD, two post-hoc analyses were therefore undertaken.

##### Single SNP MR

For MR instruments to be valid they must act as a robust proxy for exposure randomization (34). In a two-sample setting, it is conventional to include multiple instruments to strengthen the signal. However, if a SNP has a particularly robust association *and* strong biological relevance, this SNP alone may be a plausible instrument (36). Outlier SNP rs2657879 demonstrated a statistically significant association with Glutamine that was notably stronger than any other Glutamine instruments (*p*=3×10^-70^), indicating a particularly robust association. It is also located within the genomic region of glutaminase-2 (GLS2), a protein-encoding gene for the enzyme responsible for converting glutamine to glutamate as part of the glutamine-glutamate cycle (Supplementary Fig. S2-S3) (42–44). Moreover, it was the only reported GSH for glutamine in an earlier GWAS (*p*=6*10^-18^), whereby glutamine was quantified using liquid-chromatography mass spectrometry (LCMS) in a sample of *N=*7,825 (45). We therefore carried out a post-hoc single-SNP MR using this independent dataset (45), setting only rs2657879 as the IV. The Wald-ratio was used to assess causality (46).

##### Sub-threshold MR

At the opposite extreme, we investigated how increasing SNP IV numbers might improve power in the absence of rs2657879. Relaxing *p*-value thresholds risks introducing pleiotropic instruments. However, we used knowledge from our PRS analyses to inform the sub-threshold selected, relaxing this to independent SNPs significant at *p*<0.0001, with rs2657879 excluded. Data preparation followed that outlined for primary analyses.

Information on further post-hoc analyses can be found within Supplementary Information S8.

## Results

### Polygenic association between 34 metabolites and Alzheimer’s Disease

Following data preparation, 106 metabolites (Supplementary Dataset S1) and 4,725 AD case-control samples (GERAD1=3,191, ADNI=886, ANM=648. Supplementary Dataset S8) were taken forward for PRS. Thirty-four metabolites were? genetically associated with AD at *p*<0.05 when PRS were meta-analysed (82% lipoprotein subunits, 6% lipid extracts, and 12% amino or fatty acids (Fig. 3. Supplementary Dataset S7). No significant heterogeneity was evident (*I*^2^<0.5, *Q*-p>0.05) (Supplementary Dataset S7). No PRS reached adjusted significance (α=0.0002), but glutamine (Gln) came close at *p*=0.0009 (*se*=0.03, *P_T_* =0.0001) (Supplementary Dataset S7). Therefore, 34 metabolites significant at *p*<0.05 were taken forward for causal analyses.

**Figure 3.**
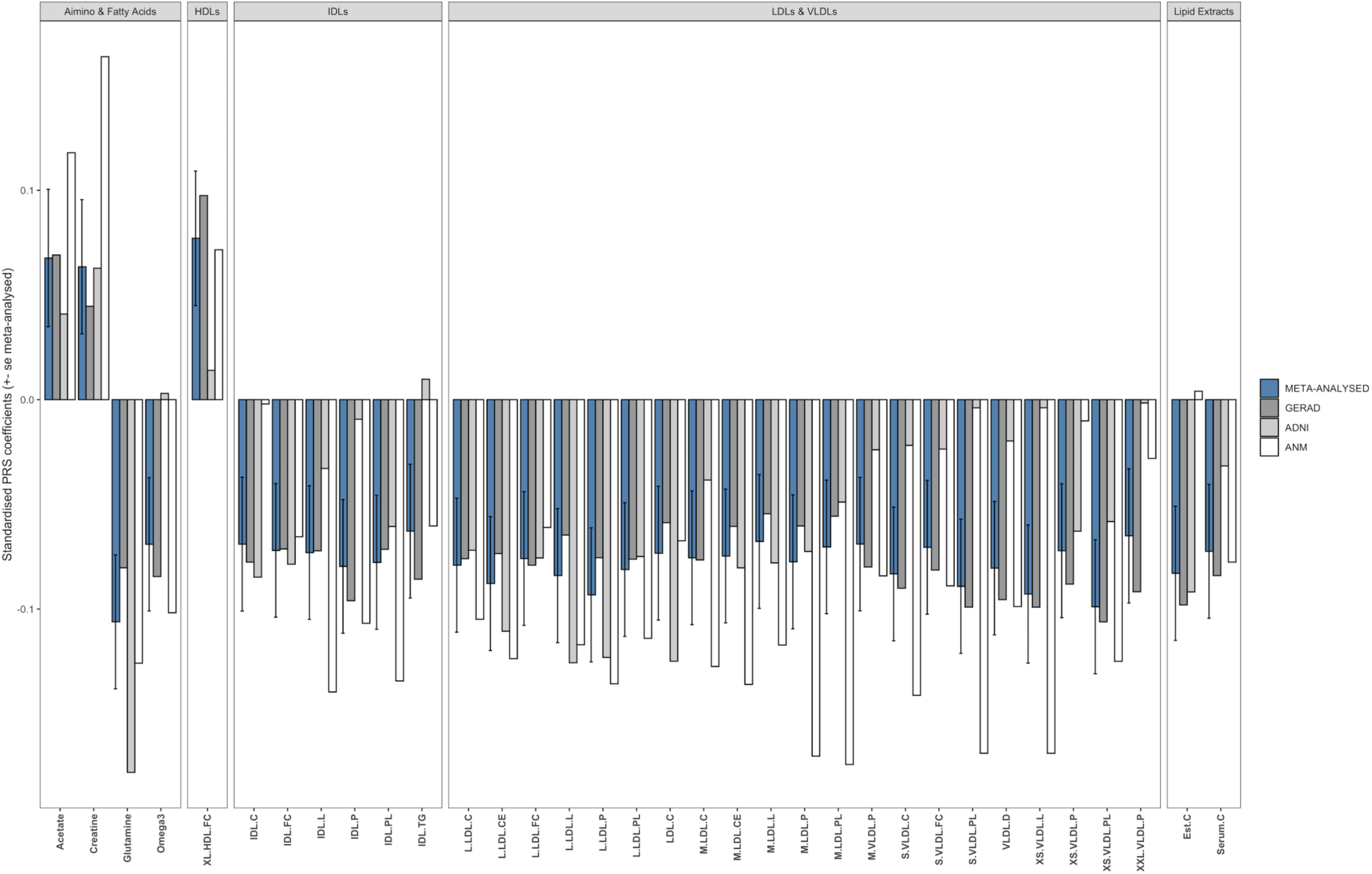
Bar chart demonstrating the magnitude of effect and meta-analysed standard errors for metabolite-AD cross-trait polygenic score associations. Results are displayed for meta-analysed associations significant at p<0.05. Meta-analysed coefficients are represented by blue bars. Non-meta-analysed coefficients are displayed in grey hues for comparison. Metabolites are grouped into metabolite-families. HDLs=High density lipoproteins, IDLs=Intermediate density lipoproteins, LDLs=Low density lipoproteins, VLDLs=Very low density lipoproteins, XL=extra large, L=large, M=medium, S=small, XS=extra small, XXL=extra extra large, C=total cholesterol, CE=cholesterol esters, FC=free cholesterol, L=lipids, PL=phospholipids, P=Concentration of particles, TG=triglycerides, D=mean diameter, Est.C=free cholesterol to esterified cholesterol ratio.

### Protective causal effect of glutamine and XL.HDL.FC on Alzheimer’s Disease

To account for multiple testing, an adjusted α=0.004 was computed for MR analyses (Supplementary Information S5). Two metabolites – glutamine and free cholesterol in XL.HDL (XL.HDL.FC) – demonstrated evidence of a small protective causal effect on AD (glutamine: IVW-OR=0.80, *p*=0.002; XL.HDL.FC: IVW-OR=0.83, *p*=0.001) (Fig. 4). Sensitivity analyses demonstrated consistent directionality for both Egger and weighted-median, though lower precision of Egger resulted in wider confidence intervals crossing the null (Fig. 4a). No pleiotropy was evidenced by the Egger-intercept, nor was significant heterogeneity apparent (Supplementary Dataset S9). MR-BMA also corroborated, ranking XL.HDL.FC and glutamine with the highest marginal inclusion probability, indicative of being the strongest “true causal” candidates of those analysed (Supplementary Dataset S19). Both also represented the most frequent metabolites within group models (Supplementary Information S9. Supplementary Dataset S20). Leave-one-out did, however, identify one influential instrument for glutamine (rs2657879), the removal of which resulted in effect attenuation and confidence intervals crossing the null (IVW-OR=0.82, 95% CI=0.62-1.10. Supplementary Figure S12). No other metabolite demonstrated evidence of a causal effect on AD (Supplementary Dataset S9, Supplementary Figure S4a-S4g). When causation was investigated in the opposite direction, no evidence was found for a causal effect of AD on metabolites (Supplementary Dataset S10, Supplementary Figure S5a-S4h).

**Figure 4.**
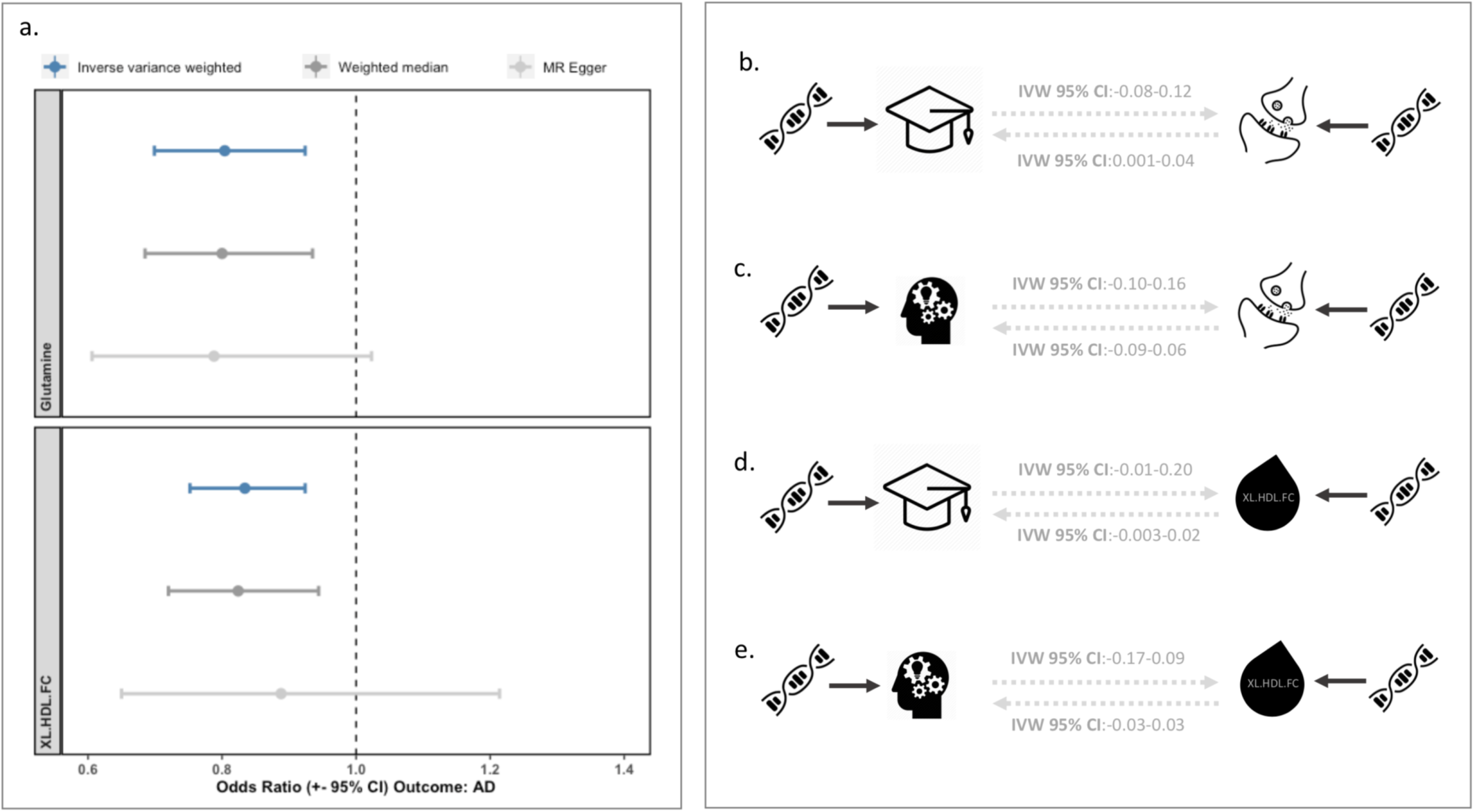
Causal effects of Glutamine and XL.HDL.FC. (a) Forest plot illustrating odds ratio point estimates and 95% confidence intervals for the effect of glutamine (top panel) and XL.HDL.FC (bottom panel) on AD in primary univariable analyses (blue bars) and secondary analyses (Weighted median=dark grey hue, Egger=light grey hue). (b) causal diagram and 95% confidence intervals from primary univariable MR of the causal effect of education on glutamine (top arrows), and of glutamine on education (bottom arrow). (c) causal diagram and 95% confidence intervals from primary univariable MR of the causal effect of cognition on glutamine (top arrows), and of glutamine on cognition (bottom arrow). (d) causal diagram and 95% confidence intervals from primary univariable MR of the causal effect of education on XL.HDL.FC (top arrows), and of XL.HDL.FC on education (bottom arrow). (e) causal diagram and 95% confidence intervals from primary univariable MR of the causal effect of cognition on XL.HDL.FC (top arrows), and of XL.HDL.FC on cognition (bottom arrow). Double helix icons represent genetic instrumental variables used in MR analyses. Causal arrows and confidence intervals are displayed in greyed hue to represent the non-significant causal relationship observed across these variables, indicating no evidence of shared causal pathways.

### Negative causal effect of cognition on lipid-related metabolites

A nominally significant causal effect of cognition on eighteen lipid-related metabolites was observed. These metabolites primarily belonged to the low- or very low- density lipoprotein family (*N*=11). All associations were in the negative direction (Supplementary Figure S6), indicating that higher cognition may result in lowered levels of these metabolites. However, no estimate survived multiple test corrections, and inconsistent directionality was seen for seventeen of the eighteen metabolites in Egger sensitivity analyses (Supplementary Dataset S11). No evidence of a causal effect in the opposite direction from metabolite to cognition was found (Supplementary Dataset S12).

### Causal association between educational attainment and lipid-related metabolites

A nominally significant causal effect of education on nine lipid-related metabolites (Supplementary Figure S7. Supplementary Dataset S13) was found. One metabolite, triglycerides in intermediate density lipoproteins (IDL.TG), remained significant at the adjusted level (*p*=0.002). This effect was in the negative direction (IVW-β=-0.18, 95% CI=-0.29 - -0.07), indicating that higher education results in lower levels of IDL.TG. Directionality was consistent for both Egger and weighted-median and no pleiotropy was evidenced by the Egger-intercept (Supplementary Dataset S13).

When causality was investigated in the opposite direction, from metabolite to education attainment, omega-3 (FAω3) was the only metabolite to demonstrate evidence of a causal relationship with education. This was in the positive direction, though the magnitude of effect was small, and significance was retained at the nominal level only (IVW-β=0.02, 95% CI=0.01-0.46, p=0.04) (Supplementary Dataset S14). FAω3 was also one of nine metabolites associated with education in the opposite direction (IVW-β=0.16, 95% CI=0.02-0.30, p=0.03). However, instrument heterogeneity was evident (Q-*p*=0.01), and inconsistent directionality was observed for Egger (Supplementary Dataset S13).

### Protective causal effect of cognition and educational attainment on Alzheimer’s Disease, with bi-directional mediation

Education and cognition both demonstrated evidence of a negative causal association with AD (Supplementary Figure S8-S9). These were both significant at the adjusted level, indicating a protective effect (education: IVW-OR=0.72, 95% CI=0.61-0.84, p=7.34×10^-05^; cognition: IVW-OR=0.73, 95% CI=0.60-0.90, p=0.002. Supplementary Dataset S15). Sensitivity analyses demonstrated consistent directionality for both Egger and weighted-median. Wider confidence intervals were observed for Egger estimates, though no pleiotropy was indicated by the Egger-intercept (Supplementary Dataset S15). Some instrument heterogeneity was evident for cognition (*Q-p*=0.01), though leave-one-out identified no significant estimate change following per-instrument removal (Supplementary Figure S14-S15). No evidence of a causal effect in the opposite direction was found (Supplementary Dataset S16, Supplementary Figure S10-S11).

A bidirectional positive causal relationship between cognition and education was also observed (Fig. 5a). Effect magnitude was, however, larger in the direction of education to cognition (education to cognition: IVW-β=0.67, 95% CI=0.63-0.71; cognition to education: IVW-β=0.30, 95% CI=0.27-0.33. Supplementary Dataset S17).

**Figure 5.**
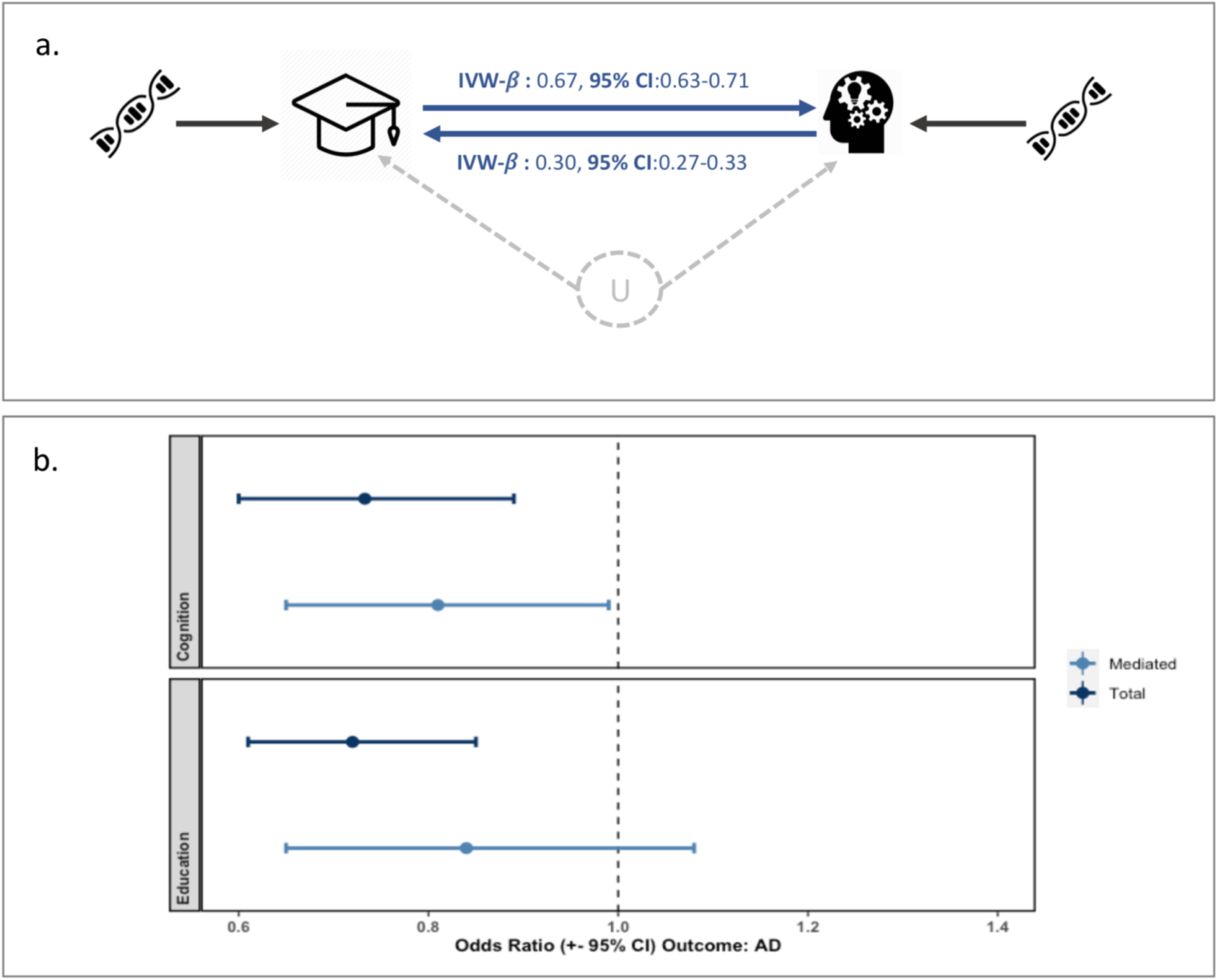
Bidirectional effects of intelligence and educational attainment. (a) Causal diagram illustrating the univariable bidirectional relationship between education (left) and cognition (right). Double helix icons on the far left and far right represent genetic instrumental variables(_1…j_)(IV). U represents unobserved confounding (allowable provided no IV to U relationship). The darker hue arrow confirms the causal estimate when x=education and y=cognition. The lighter hue arrow confirms the causal estimate when x=cognition and y=education. (b) Forest plot confirming the total (univariable) causal estimate of education and cognition on AD (darker hue), and the mediated causal estimate of education on AD when controlling for cognition, and cognition on AD when controlling for education (lighter hue).

MVMR also demonstrated evidence of bidirectional mediation between cognition and education with respect to their effect on AD. Mediation via cognition was, however, stronger than via education. More specifically, when cognition was introduced into education-AD models, the causal estimate of education on AD became smaller in magnitude (MV-IVW-OR=0.84) and was no longer significant (p=0.17), indicating total mediation via cognition (Fig. 5b. Supplementary Dataset S18). While the magnitude of the cognition-AD relationship also dropped when introducing education (MV-IVW-OR=0.81), the direct effect of cognition retained nominal significance, indicating only partial mediation via education (p=0.049).

### No evidence of mediation between metabolites and cognitive factors on Alzheimer’s Disease

Neither glutamine nor XL.HDL.FC demonstrated evidence of causally effecting education or cognition (Fig. 4b-4e. Supplementary Dataset S12, S14). Thus, the effect of both these metabolites on AD was deemed independent of education and cognition, with no multivariable model necessary. Similarly, while a number of suggestive associations between cognition and metabolites were observed (see *Causal effect of cognition on lipid-related metabolites*), these were in the direction of cognition to metabolite. As none of these metabolites demonstrated evidence of causally affecting AD (Supplementary Dataset S9), a significant b-path required for mediation was absent, and the effect of cognition on AD deemed independent of these metabolites. There was one metabolite – FAω3 – which demonstrated suggestive evidence of contributing to increased EA (Supplementary Dataset S14). Investigating EA as a mediator on the causal pathway from FAω3 to AD, however, demonstrated no evidence of a mediating effect (Supplementary Dataset S18).

For a further breakdown of results from sensitivity analyses across all MR models, see Supplementary Information S9.

### Post-hoc analyses

The Wald-ratio was used to re-estimate the causal association between glutamine and AD using only the influential SNP rs2657879 in an independent dataset (45) (Supplementary Dataset S21). Results corroborated primary analyses with notably greater magnitude of effect, though lower precision (OR=0.035, 95% CI=0.003- 0.381, p=0.006. Supplementary Dataset S22, Fig. 6).

**Figure 6.**
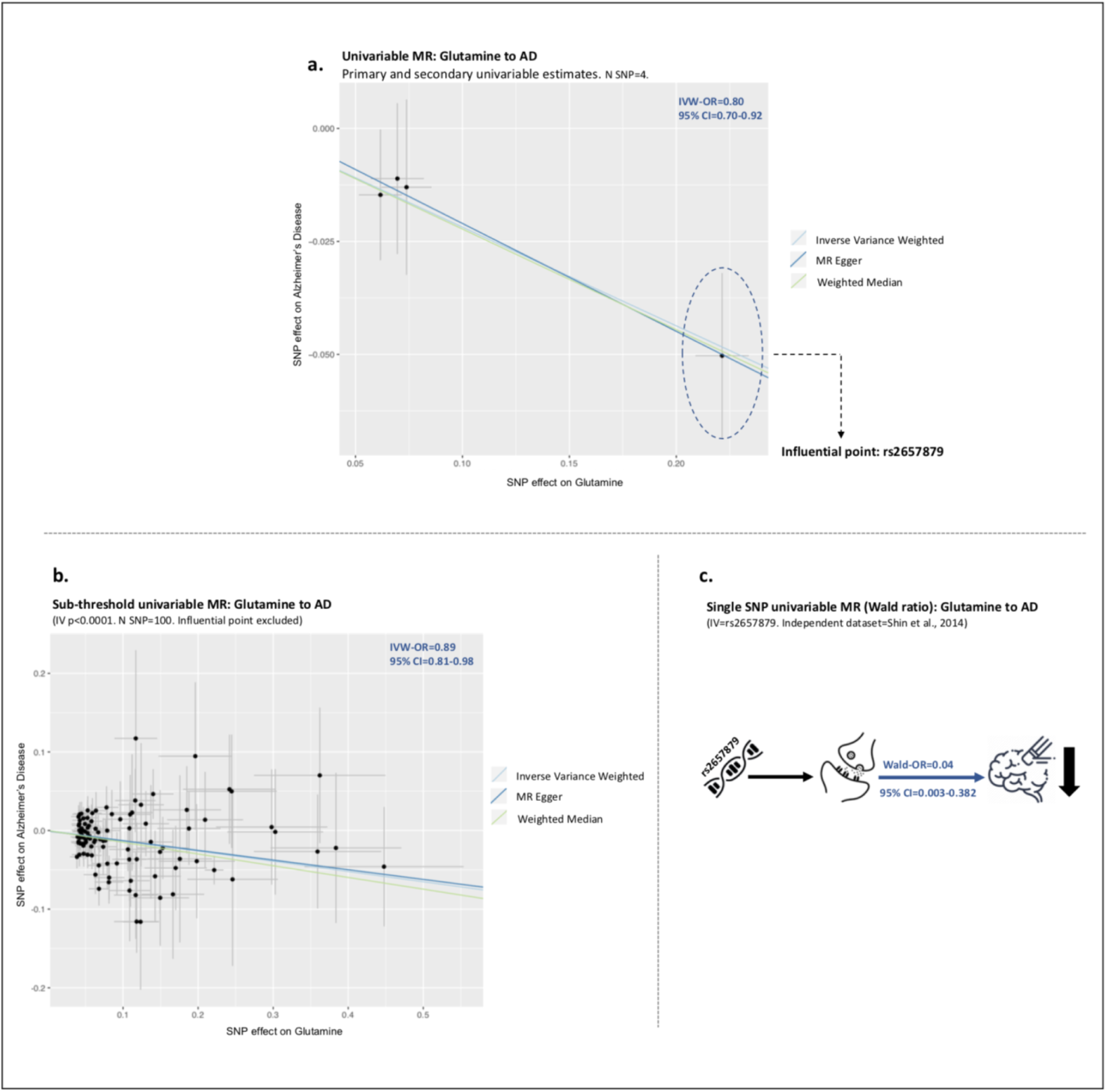
Corroboration between primary, secondary and post-hoc estimates of glutamine on AD. (a) Scatter plot illustrating pooled per-instrument estimates of the effect of glutamine on AD within primary inverse variance weighted (IVW) analyses (light blue slope), and secondary MR-Egger (dark blue slope) and weighted median (green slope) analyses. For each instrument, 95% confidence intervals are displayed for (i) SNP-outcome estimates (vertical bars) and (ii) SNP exposure estimates (horizontal bars). Slopes across the three MR estimates indicate comparable protective causal estimates. Influential point rs2657879 is highlighted (far right). (b) Scatter plot illustrating pooled per-instrument estimates for the effect of glutamine on AD within post-hoc sub-threshold analyses, with independent instruments (R2=0) associated with glutamine at p<0.0001 included in analyses (most predictive cross-trait PRS threshold when glutamine scoring models used to predict AD). Slopes for IVW, Egger and weighted median sub-threshold estimates exclude influential SNP rs2657879. (c) Directed acyclic graph (DAG) and corresponding causal estimate and 95% confidence intervals for the effect of glutamine on AD within post-hoc analyses using influential point rs2657879 as a single SNP IV estimated using the Wald ratio, and in an independent dataset from Sin et al., 2014. Far left icon represents genetic instrumental IV, middle icon represents glutamine as X, far right icon represents AD as Y, with downward arrow signifying a reduced effect of AD given higher levels of glutamine.

We also investigated the effect of increasing the number of SNP IVs (rs2657879 excluded) on glutamine’s causal estimates. Using PRS to inform the sub-threshold selected, independent instruments (r^2^=0) at *p*<0.0001 were included (IV-N=100) (Supplementary Dataset S23). Evidence of a protective causal effect was observed with a comparable magnitude of effect as primary results (OR=0.89, 95% CI=0.81-0.98, p=0.0009. Fig. 6. Supplementary Dataset S22).

## Discussion

To our knowledge, this is the first study to triangulate knowledge across polygenic scoring, univariable, and multivariable Mendelian randomization (MR) to disentangle causal relationships between blood metabolites, cognitive factors, and AD. Polygenic scores allowed us to identify, from a wider set of available metabolites, those demonstrating plausible genetic overlap with AD. MR then allowed us to interrogate both direct and indirect causality, identifying two metabolites - glutamine and free cholesterol in extra-large high-density lipoproteins (XL.HDL.FC) to have direct AD-protective effects, independent of educational attainment or cognition. An AD protective effect was also confirmed for both education and cognition, though no evidence of mediation via any of our metabolites was evident. There was, however, bidirectional mediation confirmed between education and cognition, with the mediating effect of cognition being strongest. In other words, education’s protective effect appears to work almost entirely through its positive effect on cognition and this in turn reduces AD risk.

### Glutamine and Alzheimer’s Disease

Glutamine demonstrated the strongest genetic overlap with AD in cross-trait polygenic scoring. It was also one of only two metabolites demonstrating a causal association with AD in MR. Though sensitivity analyses indicated that glutamine’s causal estimate was primarily driven by influential SNP rs2657879, post-hoc analyses added weight to initial conclusions, with single-SNP replication as well as relaxed IV inclusion (with rs2657879 excluded) corroborating primary findings. Glutamine also has particular biological relevance as it is critically implicated in neuronal transmission as part of the glutamine-glutamate cycle (42, 44) (Supplementary Figure. S3). The leading view in wider literature is that glutamine is indeed AD-protective (47, 48), though some positive associations, both with AD and lower cognition have also been reported (47, 49). The small but statistically significant protective effect we found adds weight to the former. Given glutamine’s intrinsic link to glutamate, findings also implicitly implicate glutamate in AD aetiology. This assertion would align with wider literature, which has indeed found a link between this metabolite and AD pathology (47). Unfortunately, glutamate was unable to be interrogated directly within our study due to its non-availability in Kettunen et al. datasets (22), and while it was available within a smaller GWAS (45), no genome-wide significant SNPs were identified. It will be an important endeavour to incorporate this metabolite into analyses once instruments become available.

### XL.HDL.FC and Alzheimer’s Disease

As with our previous study (20), XL.HDL.FC demonstrated a protective causal effect on AD. This aligns with wider literature which consistently regards HDLs as health promoting, while LDLs and IDLs are their risk-increasing counterparts (50–52). HDLs are implicated in reduced cognitive decline and AD more specifically (47, 53), and hold biological relevance having shown evidence of protection against neuroinflammation and cerebral-amyloid-angiopathy (53).

Three additional HDL sub-fractions, as well as one marker of inflammation – glycoprotein acetyls – demonstrated evidence of causal association with AD in our previous study (20). These were not estimated here due to their lack of polygenic association with AD. This highlights the importance of adopting an expanded repertoire of screening methods for identifying causal candidates, with previous selection based on phenotypic associations with midlife cognition (20), while selection here concentrated on genetic overlap with AD-specific diagnosis.

### Cognitive factors and Alzheimer’s Disease

A strong bi-directional causal relationship was observed between education and cognition, indicating a causal feedback-loop. In line with this, both education and cognition demonstrated a total (non-mediated) protective effect on AD, with similar magnitudes of effect observed. Additionally, when the direct effect for each of these factors was measured with consideration of the other, evidence of bi-directional mediation was present. However, while cognition was only partially mediated by education, the independent effect of education was entirely attenuated by education, suggesting that the protective effect of education on AD is due to its positive effect on cognition. These results mirror findings from a recent study (5) using smaller scaled data from (i) the IGAP consortium for AD (54), (ii) Okbay et al. for education (55), and (iii) Hill et al. for cognition (56). Here too, a bi-directional relationship between education and cognition, and a mediating effect of cognition on the education-AD relationship was found (5). To our knowledge, ours is the first study to successfully replicate these 2020 findings on larger, independent data. Such results also have implications for policy, indicating that those aimed at increasing levels of cognition by prolonging access to education may have beneficial consequences for later AD incidence.

### Cognitive factors and metabolites

Evidence was found for education leading to lower levels of triglycerides in intermediate density lipoproteins (IDL.TG). This was the only causal association to survive multiple test correction when investigating cognitive factors and metabolites. IDLs lie between low and very low density lipoproteins, and therefore most closely resemble LDLs, a family of metabolites consistently associated with adverse vascular outcomes (57–60). Similarly, higher triglyceride levels have been implicated in poor neurocognitive outcomes (61–63) and have previously shown associations with dementia (64). Interestingly, while our results indicated that education may reduce levels of this potentially harmful metabolite, there was no convincing evidence that this translated through to cognition; indicated by a non-significant relationship with the latter. There was also no evidence of causal links to AD. There may be several reasons for this. First, taking into account the magnitude of education’s effect on IDL.TG, it may be that while education has a small effect on this metabolite, this is not of large enough magnitude to translate through to a detectable indirect effect on subsequent cognition. As the effect of education on AD is totally mediated by cognition, this could then mean that any indirect effect of IDL.TG on AD would not be observed due to the absent path from IDL.TG to cognition. Alternatively, results could indicate two independent pathways from education, one leading to lowered IDL.TG, and one leading to improved cognition and downstream AD protection. The third possibility relates to power. Many more instruments were available for both education (*N*=277) and cognition (*N*=133) relative to metabolites (max.*N*=15, IDL.TG *N*=12). For lipid-related metabolites such as IDL.TG, a large proportion of genetic signal was also removed through exclusion of APOE-related instruments. This was necessary due to known pleiotropy (34). However, coupled with a low number of IVs to begin with, this likely attenuated detectable causal signals. Inferences regarding the extent of IDL.TG’s impact on cognition and AD should remain conservative until larger powered samples become available.

Several additional causal associations were observed, but which failed to survive multiple testing. We advise caution in overinterpreting these, and thus refrain from discussing them here. A brief interpretation can be found within Supplementary Information S10.

### Limitations

Metabolites studied here were limited to those available within the GWAS literature. As a result, only 123 were available for investigation. The human metabolome is estimated to contain over 250,000 metabolites (65). The extent to which we have captured all relevant metabolic mechanisms within our study therefore remains doubtful. Moreover, of those which were studied, quantification relied on NMR spectroscopy, a method with limited specificity relative to alternative methods such as mass spectrometry (MS). It is worth noting that GWAS data for a larger number of MS-quantified metabolites (N≈400) were available at the time of study (45). However, sample power (N≈7,824) lagged considerable behind data used within our study (N≈24,925), resulting in fewer available IVs. Kettunen (22) was selected over Shin (45) for this reason, although Shin provided an accessible independent dataset for post-hoc interrogation of an influential point identified for glutamine. Moreover, while breadth and specificity were sub-optimal in our selected study-data (22), key metabolites previously implicated in cognition, cognitive decline and dementia were indeed present.

For PRS and MR, AD was also quantified using a binary measure of AD diagnosis. However, AD clinical manifestations become apparent only after a long symptom-free prodromal phase (66). It remains plausible, therefore, that our clinical phenotype contains noise, with prodromal AD cases incorrectly classified amongst controls. It is worth noting that an attempt to avoid such noise was made throughout. For PRS, a conservative age-matched cut-off of 70 years for both cases and controls was implemented to avoid prodromal cases contaminating control samples where possible. For MR, our smaller GWAS of clinically diagnosed individuals was selected over a larger alternative containing additional “AD-by-proxy” samples, where diagnosis was derived from self-reported parental dementia (8). Nonetheless, quantification of the AD phenotype, rather than relying on potentially erroneous diagnostic boundaries, should be sought in future studies to improve signal to noise ratio. This could be achieved using an endophenotype approach, or through use of existing imaging or CSF biomarkers (67–69) as biological AD proxies.

### Summary

Combining knowledge across polygenic scores, multivariable and univariable MR, our results identified two blood metabolites – glutamine and XL.HDL.FC – with evidence of contributary protective effects on AD. The biological mechanisms underpinning the relationship between education, cognition and AD remain elusive, with no evidence of mediation via any of our metabolites. However, the effect of education on AD was shown to be almost entirely driven by positive changes that education has on cognition, implying that methods aimed at increasing cognition either indirectly through education, or directly via brain training, could hold protective utility against AD risk. Disentangling wider, multi-modal risk factors and understanding how these connect along the AD causal pathway will be an important future endeavour if we hope to appropriately inform treatment strategies. This study provides some important, initial pieces to this AD causal puzzle, offering biological and non-biological sources of insight to feed into this wider multi-modal work.

## Data Availability

Raw genomic data utilised within this study is available, on request, from the corresponding study consortia. Summary level data is publicly available to download via the web. The output following all statistical analyses performed within this paper can be found either within the results section of the main manuscript body, or via supplementary tables provided.

## Acknowledgements

We would like to thank members of the Statistical Genetics Unit at the Social, Genetic, and Developmental Psychiatry Centre (SGDP), King’s College London, for their continued support and discussion on methodological aspects of this study. This work was also made possible only through generous funding from key funding bodies. JL is funded by the van Geest endowment fund and PP is funded by Alzheimer’s Research UK. MR is supported by the Medical Research Council (MC_UU_12019/3). This study represents independent research additionally funded by the National Institute for Health Research (NIHR) Biomedical Research Centre at South London and Maudsley NHS Foundation Trust and King’s College London. The views expressed are those of the author(s) and not necessarily those of the NHS, the NIHR or the Department of Health and Social Care.

ADNI: A proportion of data collection and sharing for this project was also funded by the Alzheimer’s Disease Neuroimaging Initiative (ADNI) (National Institutes of Health Grant U01 AG024904). ADNI is funded by the National Institute on Aging, the National Institute of Biomedical Imaging and Bioengineering, and through generous contributions from the following: Abbott; Alzheimer’s Association; Alzheimer’s Drug Discovery Foundation; Amorfix Life Sciences Ltd.; AstraZeneca; Bayer HealthCare; BioClinica, Inc.; Biogen Idec Inc.; Bristol-Myers Squibb Company; Eisai Inc.; Elan

Pharmaceuticals Inc.; Eli Lilly and Company; F. Hoffmann-La Roche Ltd and its affiliated company Genentech, Inc.; GE Healthcare; Innogenetics, N.V.; Janssen Alzheimer Immunotherapy Research & Development, LLC.; Johnson & Johnson Pharmaceutical Research & Development LLC.; Medpace, Inc.; Merck & Co., Inc.; Meso Scale Diagnostics, LLC.; Novartis Pharmaceuticals Corporation; Pfizer Inc.; Servier; Synarc Inc.; and Takeda Pharmaceutical Company. The Canadian Institutes of Health Research is providing funds to support ADNI clinical sites in Canada. Private sector contributions are facilitated by the Foundation for the National Institutes of Health (www.fnih.org). The grantee organization is the Northern California Institute for Research and Education, and the study is coordinated by the Alzheimer’s Disease Cooperative Study at the University of California, San Diego. ADNI data are disseminated by the Laboratory of Neuro Imaging at the University of California, Los Angeles.

GERAD1: Cardiff University was supported by the Wellcome Trust, Medical Research Council (MRC), Alzheimer’s Research UK (ARUK) and the Welsh Assembly Government. Cambridge University and Kings College London acknowledge support from the MRC. ARUK supported sample collections at the South West Dementia Bank and the Universities of Nottingham, Manchester and Belfast. The Belfast group acknowledges support from the Alzheimer’s Society, Ulster Garden Villages, N.Ireland R&D Office and the Royal College of Physicians/Dunhill Medical Trust. The MRC and Mercer’s Institute for Research on Ageing supported the Trinity College group. The South West Dementia Brain Bank acknowledges support from Bristol Research into Alzheimer’s and Care of the Elderly. The Charles Wolfson Charitable Trust supported the OPTIMA group.

Washington University was funded by NIH grants, Barnes Jewish Foundation and the Charles and Joanne Knight Alzheimer’s Research Initiative. Patient recruitment for the MRC Prion Unit/UCL Department of Neurodegenerative Disease collection was supported by the UCLH/UCL Biomedical Centre and NIHR Queen Square Dementia Biomedical Research Unit. LASER-AD was funded by Lundbeck SA. The Bonn group was supported by the German Federal Ministry of Education and Research (BMBF), Competence Network Dementia and Competence Network Degenerative Dementia, and by the Alfried Krupp von Bohlen und Halbach-Stiftung. The GERAD1 Consortium also used samples ascertained by the NIMH AD Genetics Initiative.

## Author Contributions

JL performed all data pre-processing, statistical analyses, and manuscript writing. PP supervised the study, provided statistical input, and reviewed the manuscript. RG provided statistical input and reviewed the manuscript. SWC aided PRS statistical insight and back-conversion of R2 statistics and reviewed the manuscript. CH aided in establishing an appropriate tool for correlated multiple test corrections and reviewed the manuscript. DA and LV provided ANM data and reviewed the manuscript. MR, PC, CLQ & RD provided background support and reviewed the manuscript.

## References

1. Prince M, Comas-Herrera A, Knapp M, Guerchet M, Karagiannidou M. World Alzheimer report 2016: improving healthcare for people living with dementia: coverage, quality and costs now and in the future [Internet]. London, UK: Alzheimer’s Disease International (ADI); 2016 [cited 2020 Nov 3]. Available from: http://www.alz.co.uk/

2. Sharp ES, Gatz M. Relationship between education and dementia: an updated systematic review. Alzheimer Dis Assoc Disord. 2011 Dec;25(4):289–304.

3. Yeo RA, Arden R, Jung RE. Alzheimer’s disease and intelligence. Curr Alzheimer Res. 2011 Jun;8(4):345–53.

4. Livingston G, Huntley J, Sommerlad A, Ames D, Ballard C, Banerjee S, et al. Dementia prevention, intervention, and care: 2020 report of the Lancet Commission. The Lancet. 2020 Aug 8;396(10248):413–46.

5. Anderson EL, Howe LD, Wade KH, Ben-Shlomo Y, Hill WD, Deary IJ, et al. Education, intelligence and Alzheimer’s disease: evidence from a multivariable two-sample Mendelian randomization study. Int J Epidemiol. 2020 Jan 31;

6. Anderson EL, Wade KH, Hemani G, Bowden J, Korologou-Linden R, Smith GD, et al. The causal effect of educational attainment on Alzheimer’s disease: A two-sample Mendelian randomization study. bioRxiv. 2017 Apr 17;127993.

7. Larsson SC, Traylor M, Malik R, Dichgans M, Burgess S, Markus HS. Modifiable pathways in Alzheimer’s disease: Mendelian randomisation analysis. BMJ [Internet]. 2017 Dec 7 [cited 2020 Feb 28];359. Available from: https://www.bmj.com/content/359/bmj.j5375

8. Jansen IE, Savage JE, Watanabe K, Bryois J, Williams DM, Steinberg S, et al. Genome-wide meta-analysis identifies new loci and functional pathways influencing Alzheimer’s disease risk. Nat Genet. 2019;51(3):404–13.

9. Raghavan NS, Vardarajan B, Mayeux R. Genomic variation in educational attainment modifies Alzheimer disease risk. Neurol Genet. 2019 Apr;5(2):e310.

10. Enche Ady CNA, Lim SM, Teh LK, Salleh MZ, Chin A-V, Tan MP, et al. Metabolomic-guided discovery of Alzheimer’s disease biomarkers from body fluid. J Neurosci Res. 2017;95(10):2005–24.

11. Brink-Jensen K, Bak S, Jørgensen K, Ekstrøm CT. Integrative analysis of metabolomics and transcriptomics data: a unified model framework to identify underlying system pathways. PloS One. 2013;8(9):e72116.

12. Snowden S, Dahlén S-E, Wheelock CE. Application of metabolomics approaches to the study of respiratory diseases. Bioanalysis. 2012 Sep;4(18):2265–90.

13. Whiley L, Sen A, Heaton J, Proitsi P, García-Gómez D, Leung R, et al. Evidence of altered phosphatidylcholine metabolism in Alzheimer’s disease. Neurobiol Aging. 2014 Feb 1;35(2):271–8.

14. Proitsi P, Kim M, Whiley L, Simmons A, Sattlecker M, Velayudhan L, et al. Association of blood lipids with Alzheimer’s disease: A comprehensive lipidomics analysis. Alzheimers Dement. 2017 Feb 1;13(2):140–51.

15. Wang G, Zhou Y, Huang F-J, Tang H-D, Xu X-H, Liu J-J, et al. Plasma metabolite profiles of Alzheimer’s disease and mild cognitive impairment. J Proteome Res. 2014 May 2;13(5):2649–58.

16. González-Domínguez R, García-Barrera T, Gómez-Ariza JL. Metabolite profiling for the identification of altered metabolic pathways in Alzheimer’s disease. J Pharm Biomed Anal. 2015 Mar 25;107:75–81.

17. Smith GD, Ebrahim S. ‘Mendelian randomization’: can genetic epidemiology contribute to understanding environmental determinants of disease? Int J Epidemiol. 2003 Feb;32(1):1–22.

18. Sanderson E, Davey Smith G, Windmeijer F, Bowden J. An examination of multivariable Mendelian randomization in the single-sample and two-sample summary data settings. Int J Epidemiol. 2019 Jun 1;48(3):713–27.

19. Burgess S, Thompson DJ, Rees JMB, Day FR, Perry JR, Ong KK. Dissecting Causal Pathways Using Mendelian Randomization with Summarized Genetic Data: Application to Age at Menarche and Risk of Breast Cancer. Genetics. 2017;207(2):481–7.

20. Lord J, Jermy B, Green R, Wong A, Xu J, Legido-Quigley C, et al. Deciphering the causal relationship between blood metabolites and Alzheimers Disease: a Mendelian Randomization study. medRxiv. 2020 Apr 30;2020.04.28.20083253.

21. Proitsi P, Kuh D, Wong A, Maddock J, Bendayan R, Wulaningsih W, et al. Lifetime cognition and late midlife blood metabolites: findings from a British birth cohort. Transl Psychiatry. 2018 Sep 26;8(1):1–11.

22. Kettunen J, Demirkan A, Würtz P, Draisma HHM, Haller T, Rawal R, et al. Genome-wide study for circulating metabolites identifies 62 loci and reveals novel systemic effects of LPA. Nat Commun. 2016 Mar 23;7:11122.

23. Weiner MW, Aisen PS, Jack CR, Jagust WJ, Trojanowski JQ, Shaw L, et al. The Alzheimer’s disease neuroimaging initiative: progress report and future plans. Alzheimers Dement J Alzheimers Assoc. 2010 May;6(3):202–211.e7.

24. Lovestone S, Francis P, Kloszewska I, Mecocci P, Simmons A, Soininen H, et al. AddNeuroMed--the European collaboration for the discovery of novel biomarkers for Alzheimer’s disease. Ann N Y Acad Sci. 2009 Oct;1180:36–46.

25. Sattlecker M, Kiddle SJ, Newhouse S, Proitsi P, Nelson S, Williams S, et al. Alzheimer’s disease biomarker discovery using SOMAscan multiplexed protein technology. Alzheimers Dement J Alzheimers Assoc. 2014 Nov;10(6):724–34.

26. Kunkle BW, Grenier-Boley B, Sims R, Bis JC, Damotte V, Naj AC, et al. Genetic meta-analysis of diagnosed Alzheimer’s disease identifies new risk loci and implicates Aβ, tau, immunity and lipid processing. Nat Genet. 2019 Mar;51(3):414–30.

27. Lee JJ, Wedow R, Okbay A, Kong E, Maghzian O, Zacher M, et al. Gene discovery and polygenic prediction from a 1.1-million-person GWAS of educational attainment. Nat Genet. 2018 Aug;50(8):1112–21.

28. Savage JE, Jansen PR, Stringer S, Watanabe K, Bryois J, de Leeuw CA, et al. Genome-wide association meta-analysis in 269,867 individuals identifies new genetic and functional links to intelligence. Nat Genet. 2018 Jul;50(7):912–9.

29. Chang CC, Chow CC, Tellier LC, Vattikuti S, Purcell SM, Lee JJ. Second-generation PLINK: rising to the challenge of larger and richer datasets. GigaScience. 2015;4:7.

30. Bulik-Sullivan BK, Loh P-R, Finucane HK, Ripke S, Yang J, Patterson N, et al. LD Score regression distinguishes confounding from polygenicity in genome-wide association studies. Nat Genet. 2015 Mar;47(3):291–5.

31. Hill WD, Davies G, Harris SE, Hagenaars SP, Liewald DC, Penke L, et al. Molecular genetic aetiology of general cognitive function is enriched in evolutionarily conserved regions. Transl Psychiatry. 2016 Dec;6(12):e980–e980.

32. Burgess S, Butterworth A, Thompson SG. Mendelian randomization analysis with multiple genetic variants using summarized data. Genet Epidemiol. 2013 Nov;37(7):658–65.

33. van der Harst P, Verweij N. Identification of 64 Novel Genetic Loci Provides an Expanded View on the Genetic Architecture of Coronary Artery Disease. Circ Res. 2018 02;122(3):433–43.

34. Bowden J, Davey Smith G, Haycock PC, Burgess S. Consistent Estimation in Mendelian Randomization with Some Invalid Instruments Using a Weighted Median Estimator. Genet Epidemiol. 2016 May;40(4):304–14.

35. Hemani G, Zheng J, Elsworth B, Wade KH, Haberland V, Baird D, et al. The MR-Base platform supports systematic causal inference across the human phenome. Loos R, editor. eLife. 2018 May 30;7:e34408.

36. Burgess S, Davey Smith G, Davies NM, Dudbridge F, Gill D, Glymour MM, et al. Guidelines for performing Mendelian randomization investigations. Wellcome Open Res. 2019;4:186.

37. Choi SW, O’Reilly PF. PRSice-2: Polygenic Risk Score software for biobank-scale data. GigaScience. 2019 01;8(7).

38. Viechtbauer W. Conducting meta-analyses in R with the metafor package. J Stat Softw. 2010 Aug;36(3):1–48.

39. MacKinnon DP, Lockwood CM, Hoffman JM, West SG, Sheets V. A comparison of methods to test mediation and other intervening variable effects. Psychol Methods. 2002;7(1):83–104.

40. Bowden J, Davey Smith G, Burgess S. Mendelian randomization with invalid instruments: effect estimation and bias detection through Egger regression. Int J Epidemiol. 2015 Apr;44(2):512–25.

41. Zuber V, Colijn JM, Klaver C, Burgess S. Selecting likely causal risk factors from high-throughput experiments using multivariable Mendelian randomization. Nat Commun. 2020 Jan 7;11(1):29.

42. Stobart JL, Anderson CM. Multifunctional role of astrocytes as gatekeepers of neuronal energy supply. Front Cell Neurosci [Internet]. 2013 [cited 2020 Nov 2];7. Available from: https://www.frontiersin.org/articles/10.3389/fncel.2013.00038/full

43. Tsepilov YA, Shin S-Y, Soranzo N, Spector TD, Prehn C, Adamski J, et al. Nonadditive Effects of Genes in Human Metabolomics. Genetics. 2015 Jul;200(3):707–18.

44. Scott WK, Medie FM, Ruffin F, Sharma-Kuinkel BK, Cyr DD, Guo S, et al. Human genetic variation in GLS2 is associated with development of complicated Staphylococcus aureus bacteremia. PLoS Genet. 2018;14(10):e1007667.

45. Shin S-Y, Fauman EB, Petersen A-K, Krumsiek J, Santos R, Huang J, et al. An atlas of genetic influences on human blood metabolites. Nat Genet. 2014 Jun;46(6):543–50.

46. Teumer A. Common Methods for Performing Mendelian Randomization. Front Cardiovasc Med [Internet]. 2018 May 28 [cited 2020 Nov 2];5. Available from: https://www.ncbi.nlm.nih.gov/pmc/articles/PMC5985452/

47. van der Lee SJ, Teunissen CE, Pool R, Shipley MJ, Teumer A, Chouraki V, et al. Circulating metabolites and general cognitive ability and dementia: Evidence from 11 cohort studies. Alzheimers Dement J Alzheimers Assoc. 2018;14(6):707–22.

48. Chen J, Herrup K. Chapter 70 - Glutamine as a Potential Neuroprotectant in Alzheimer’s Disease. In: Martin CR, Preedy VR, editors. Diet and Nutrition in Dementia and Cognitive Decline [Internet]. San Diego: Academic Press; 2015 [cited 2020 Nov 8]. p. 761–71. Available from: http://www.sciencedirect.com/science/article/pii/B9780124078246000707

49. Madeira C, Vargas-Lopes C, Brandão CO, Reis T, Laks J, Panizzutti R, et al. Elevated Glutamate and Glutamine Levels in the Cerebrospinal Fluid of Patients With Probable Alzheimer’s Disease and Depression. Front Psychiatry. 2018;9:561.

50. Wilson PW, Garrison RJ, Castelli WP, Feinleib M, McNamara PM, Kannel WB. Prevalence of coronary heart disease in the framingham offspring study: Role of lipoprotein cholesterols. Am J Cardiol. 1980 Oct 1;46(4):649–54.

51. Ouimet Mireille, Barrett Tessa J., Fisher Edward A. HDL and Reverse Cholesterol Transport. Circ Res. 2019 May 10;124(10):1505–18.

52. Bardagjy AS, Steinberg FM. Relationship Between HDL Functional Characteristics and Cardiovascular Health and Potential Impact of Dietary Patterns: A Narrative Review. Nutrients. 2019 Jun;11(6):1231.

53. Button EB, Robert J, Caffrey TM, Fan J, Zhao W, Wellington CL. HDL from an Alzheimer’s disease perspective. Curr Opin Lipidol. 2019 Jun;30(3):224–34.

54. Lambert JC, Ibrahim-Verbaas CA, Harold D, Naj AC, Sims R, Bellenguez C, et al. Meta-analysis of 74,046 individuals identifies 11 new susceptibility loci for Alzheimer’s disease. Nat Genet. 2013 Dec;45(12):1452–8.

55. Okbay A, Beauchamp JP, Fontana MA, Lee JJ, Pers TH, Rietveld CA, et al. Genome-wide association study identifies 74 loci associated with educational attainment. Nature. 2016 26;533(7604):539–42.

56. Hill WD, Marioni RE, Maghzian O, Ritchie SJ, Hagenaars SP, McIntosh AM, et al. A combined analysis of genetically correlated traits identifies 187 loci and a role for neurogenesis and myelination in intelligence. Mol Psychiatry. 2019;24(2):169–81.

57. Zeljkovic A, Vekic J, Spasojevic-Kalimanovska V, Jelic-Ivanovic Z, Bogavac-Stanojevic N, Gulan B, et al. LDL and HDL subclasses in acute ischemic stroke: prediction of risk and short-term mortality. Atherosclerosis. 2010 Jun;210(2):548–54.

58. de Oliveira J, Hort MA, Moreira ELG, Glaser V, Ribeiro-do-Valle RM, Prediger RD, et al. Positive correlation between elevated plasma cholesterol levels and cognitive impairments in LDL receptor knockout mice: relevance of cortico-cerebral mitochondrial dysfunction and oxidative stress. Neuroscience. 2011 Dec 1;197:99–106.

59. Kovács KR, Bajkó Z, Szekeres CC, Csapó K, Oláh L, Magyar MT, et al. Elevated LDL-C combined with hypertension worsens subclinical vascular impairment and cognitive function. J Am Soc Hypertens JASH. 2014 Aug;8(8):550–60.

60. Imamura T, Doi Y, Arima H, Yonemoto K, Hata J, Kubo M, et al. LDL cholesterol and the development of stroke subtypes and coronary heart disease in a general Japanese population: the Hisayama study. Stroke. 2009 Feb;40(2):382–8.

61. Parthasarathy V, Frazier DT, Bettcher BM, Jastrzab L, Chao L, Reed B, et al. Triglycerides are negatively correlated with cognitive function in nondemented aging adults. Neuropsychology. 2017;31(6):682–8.

62. Farr SA, Yamada KA, Butterfield DA, Abdul HM, Xu L, Miller NE, et al. Obesity and hypertriglyceridemia produce cognitive impairment. Endocrinology. 2008 May;149(5):2628–36.

63. Morley JE, Banks WA. Lipids and Cognition. J Alzheimers Dis. 2010 Jan 1;20(3):737– 47.

64. Bernath MM, Bhattacharyya S, Nho K, Barupal DK, Fiehn O, Baillie R, et al. Serum triglycerides in Alzheimer disease: Relation to neuroimaging and CSF biomarkers. Neurology. 2020 19;94(20):e2088–98.

65. Wishart DS, Feunang YD, Marcu A, Guo AC, Liang K, Vázquez-Fresno R, et al. HMDB 4.0: the human metabolome database for 2018. Nucleic Acids Res. 2018 04;46(D1):D608–17.

66. Bature F, Guinn B-A, Pang D, Pappas Y. Signs and symptoms preceding the diagnosis of Alzheimer’s disease: a systematic scoping review of literature from 1937 to 2016. BMJ Open. 2017 Aug 28;7(8):e015746.

67. Dhiman K, Blennow K, Zetterberg H, Martins RN, Gupta VB. Cerebrospinal fluid biomarkers for understanding multiple aspects of Alzheimer’s disease pathogenesis. Cell Mol Life Sci CMLS. 2019 May;76(10):1833–63.

68. Zetterberg H, Skillbäck T, Mattsson N, Trojanowski JQ, Portelius E, Shaw LM, et al. Association of Cerebrospinal Fluid Neurofilament Light Concentration With Alzheimer Disease Progression. JAMA Neurol. 2016 Jan;73(1):60–7.

69. Bloudek LM, Spackman DE, Blankenburg M, Sullivan SD. Review and meta-analysis of biomarkers and diagnostic imaging in Alzheimer’s disease. J Alzheimers Dis JAD. 2011;26(4):627–45.

