## Supplementary Information for "Disentangling independent and mediated causal relationships between blood metabolites, cognitive factors, and Alzheimer’s Disease"

### Document Contents:


- Supplementary Information S1-S10
- Supplementary Figures S1-S16

**Supplementary Information S1. Genetic and Environmental Risk for Alzheimer's disease Consortium (GERAD1) Collaborators**

**Names**

Denise Harold[1], Rebecca Sims[1], Amy Gerrish[1], Jade Chapman[1], Valentina Escott-Price[1], Nandini Badarinarayan[1], Richard Abraham[1], Paul Hollingworth[1], Marian Hamshere[1], Jaspreet Singh Pahwa[1], Kimberley Dowzell[1], Amy Williams[1], Nicola Jones[1], Charlene Thomas[1], Alexandra Stretton[1], Angharad Morgan[1], Kate Williams[1], Sarah Taylor[1], Simon Lovestone[2], John Powell[3], Petroula Proitsi[3], Michelle K Lupton[3], Carol Brayne[4], David C. Rubinsztein[5], Michael Gill[6], Brian Lawlor[6], Aoibhinn Lynch[6], Kevin Morgan[7], Kristelle Brown[7], Peter Passmore[8], David Craig[8], Bernadette McGuinness[8], Janet A Johnston[8], Stephen Todd[8], Clive Holmes[9], David Mann[10], A. David Smith[11], Seth Love[12], Patrick G. Kehoe[12], John Hardy[13], Rita Guerreiro[14,15], Andrew Singleton[14], Simon Mead[16], Nick Fox[17], Martin Rossor[17], John Collinge[16], Wolfgang Maier[18], Frank Jessen[18], Reiner Heun[18], Britta Schürmann[18,19], Alfredo Ramirez[18], Tim Becker[20], Christine Herold[20], André Lacour[20], Dmitriy Drichel[20], Hendrik van den Bussche[21], Isabella Heuser[22], Johannes Kornhuber[23], Jens Wiltfang[24], Martin Dichgans[25,26], Lutz Frölich[27], Harald Hampel[28], Michael Hüll[29], Dan Rujescu[30], Alison Goate[31], John S.K. Kauwe[32], Carlos Cruchaga[33], Petra Nowotny[33], John C. Morris[33], Kevin Mayo[33], Gill Livingston[34], Nicholas J. Bass[34], Hugh Gurling[34], Andrew McQuillin[34], Rhian Gwilliam[35], Panagiotis Deloukas[35], Markus M. Nöthen[20], Peter Holmans[1], Michael O'Donovan[1], Michael J.Owen[1], Julie Williams[1].

**Affiliations**

[1]Medical Research Council (MRC) Centre for Neuropsychiatric Genetics and Genomics, Neurosciences and Mental Health Research Institute, Department of Psychological Medicine and Neurology, School of Medicine, Cardiff University, Cardiff, UK. [2]Department of Psychiatry, Medical Sciences Division, University of Oxford, Oxford, UK. [3]Kings College London, Institute of Psychiatry, Department of Neuroscience, De Crespigny Park, Denmark Hill, London, UK. [4]Institute of Public Health, University of Cambridge, Cambridge, UK. [5]Cambridge Institute for Medical Research, University of Cambridge, Cambridge, UK. [6]Mercers Institute for Research on Aging, St. James Hospital and Trinity College, Dublin, Ireland. [7]Institute of Genetics, Queens Medical Centre, University of Nottingham, UK. [8]Ageing Group, Centre for Public Health, School of Medicine, Dentistry and Biomedical Sciences, Queens University Belfast, UK. [9]Division of Clinical Neurosciences, School of Medicine, University of Southampton, Southampton, UK. [10]Clinical Neuroscience Research Group, Greater Manchester Neurosciences Centre, University of Manchester, Salford, UK. [11]Oxford Project to Investigate Memory and Ageing (OPTIMA), University of Oxford, Department of Pharmacology, Mansfield Road, Oxford, UK. [12]University of Bristol Institute of Clinical Neurosciences, School of Clinical Sciences, Frenchay Hospital, Bristol, UK. [13]Department of Molecular Neuroscience and Reta Lilla Weston Laboratories, Institute of Neurology, UCL, London, UK. [14]Laboratory of Neurogenetics, National Institute on Aging, National Institutes of Health, Bethesda, Maryland, United States of America. [15]Department of Molecular Neuroscience, Institute of Neurology, University College London, Queen Square, London WC1N 3BG, UK. [16]MRC Prion Unit, Department of Neurodegenerative Disease, UCL Institute of Neurology, London, UK. [17]Dementia Research Centre, Department of Neurodegenerative Diseases, University College London, Institute of Neurology, London, UK. [18]Department of Psychiatry, University of Bonn, Sigmund-

Freud-Straβe 25, 53105 Bonn, Germany. [19]Institute for Molecular Psychiatry, University of Bonn, Bonn, Germany. [20]Department of Genomics, Life & Brain Center, University of Bonn, Bonn, Germany. [21]Institute of Primary Medical Care, University Medical Center Hamburg-Eppendorf, Germany. [22]Department of Psychiatry, Charité Berlin, Germany. [23]Department of Psychiatry, Friedrich-Alexander-University Erlangen-Nürnberg, Germany. [24]Department of Psychiatry and Psychotherapy, University Medical Center (UMG), Georg-August-University, Göttingen, Germany. [25]Institute for Stroke and Dementia Research, Klinikum der Universität München, Marchioninistr. 15, 81377, Munich, Germany. [26]Department of Neurology, Klinikum der Universität München, Marchioninistr. 15, 81377, Munich, Germany. [27]Central Institute of Mental Health, Medical Faculty Mannheim, University of Heidelberg, Germany. [28]Institute for Memory and Alzheimer's Disease & INSERM, Sorbonne Universities, Pierre and Marie Curie University, Paris, France; Institute for Brain and Spinal Cord Disorders (ICM), Department of Neurology, Hospital of Pitié-Salpétrière, Paris, France. [29]Centre for Geriatric Medicine and Section of Gerontopsychiatry and Neuropsychology, Medical School, University of Freiburg, Germany. [30]Department of Psychiatry, University of Halle, Halle, Germany. [31]Neuroscience Department, Icahn School of Medicine at Mount Sinai, New York, US. [32]Department of Biology, Brigham Young University, Provo, UT, 84602, USA. [33]Departments of Psychiatry, Neurology and Genetics, Washington University School of Medicine, St Louis, MO 63110, US. [34]Department of Mental Health Sciences, University College London, UK. [35]The Wellcome Trust Sanger Institute, Wellcome Trust Genome Campus, Hinxton, Cambridge, UK.

**Supplementary Information S2. Mendelian Randomization scope**
In-line with core MR checklist outlined by Burgess et al. (1).
*Note: this document includes scope for planned analyses only – and does not extend to post-hoc analyses.*

### 1. Motivation and scope

#### a. The primary hypotheses of interest

i. Metabolites which genetically predict AD have a causal relationship with the disease.
ii. Metabolites which genetically predict AD have a causal relationship with earlier protective factors such as education and cognition.
iii. Factors such as education and cognition have a protective causal effect on Alzheimer's Disease (AD).
iv. Education and cognition exert some of their protective effect on AD through changes to levels of genetically predictive metabolites.
v. A causally relevant feedback loop exists between education and cognition which bidirectionally reinforces their protective effect on AD.

### b. Motivation for using Mendelian Randomization

Metabolites which genetically predict Alzheimer's Disease status are indictive of possessing some degree of shared aetiology. However, association does not necessarily equal causation, and in the instance where causation *is* present, directionality of effect cannot be concluded. If metabolites are to contribute to effective sources of targeted intervention, a causal relationship must be demonstrated. MR offers a statistical technique which – in the absence of a randomized control trial – can help to tease out, from a wider set of metabolites demonstrating genetic overlap with AD, which ones may be on the causal pathway to the disease. Directionality can also be established to rule out potential reverse causation.

Motivation for incorporating multivariable analyses, lies in the knowledge that, whilst factors such as education and cognition are established risk factors for AD, the aetiological underpinnings of this relationship remain elusive. Multivariable MR allows for mediating relationships to be interrogated, which if demonstrated, could offer insight into the underlying forces driving the overarching relationship observed, and offering both direct and indirect sources of intervention.

### c. Study scope

i. Harness the use of polygenic risk scores to tease out, from a wider set of blood metabolites, those demonstrating evidence of genetically predicting clinical AD status.

ii. Of those blood metabolites demonstrating predictive genetic association with AD, utilise univariable MR to:
    1. Investigate bidirectional causal associations with each metabolite and AD; both metabolite → AD, and AD→ metabolite, to investigate the presence of reverse causation.
    2. Investigate bi-directional causal associations between each metabolite and education and each metabolite and cognition – two established AD-related risk factors.

iii. Utilise bi-directional univariable MR to re-confirm previous reports of a causal association between education and AD, and of cognition and AD.

iv. Utilise bi-directional univariable MR to confirm the causal relationship between education and cognition.

v. Utilise multivariable MR to investigate:
    1. Metabolites which may mediate the causal relationship between education and AD.

2. Metabolites which may mediate the causal relationship between cognition and AD.
3. Whether there is evidence of education mediating causal relationships between each metabolites and AD.
4. Whether there is evidence of cognition mediating causal relationships between each metabolites and AD.
5. Whether there is evidence of education mediating causal relationships between cognition and AD.
6. Whether there is evidence of cognition mediating causal relationships between education and AD.

**d. Primary MR analyses (what and how any)**

i. Univariable bidirectional MR to test the following causal relationships (Note: metabolite N is determined by the number of metabolites found to genetically predict AD case/control status in prior cross-trait polygenic score analyses):
   1. Metabolite (1…N) ←→ AD.
   2. Metabolite (1…N) ←→Education.
   3. Metabolite (1…N) ←→ Cognition.
   4. Education ←→ AD.
   5. Cognition ←→ AD.
   6. Education ←→ Cognition.

ii. Multivariable mendelian randomization to investigate:
   1. Metabolites (1 at a time) as mediators of education → AD causal associations.
   2. Metabolites (1 at a time) as mediators of cognition → AD causal associations.
   3. Education as a mediator of metabolite (1 at a time) → AD causal associations.
   4. Cognition as a mediator of metabolite (1 at a time) → AD causal associations.
   5. Education as a mediator of cognition → AD causal associations.
   6. Cognition as a mediator of education → AD causal associations

Note: For justification of multivariable analyses, univariable analyses are required to show a direct causal association between X→M (*a* path), *and* between M→Y (*b* path). For X to be included in multivariable analysis, it must show a univariable X→M relationship, but is not required to show a significant univariable X→Y (*c* path) causal association, as this may be non-significant due to suppression via M.

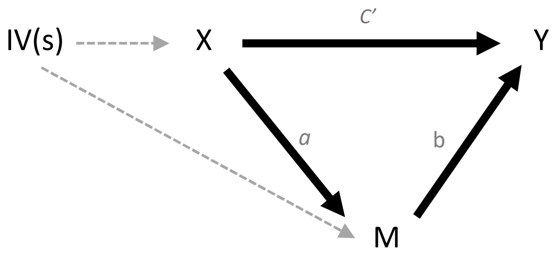


### 2. MR data sources

Summary-level GWAS data were used for all MR analyses:

i. **Metabolites:** Kettunen et al. (2).
N = ~24,925 individuals of European decent. 123 circulating blood metabolites measured using nuclear magnetic resonance imaging spectroscopy.
Participant ages range from 31-61 years (mean=44.6 years).
Of the 123 metabolites available, a thirty-four were selected for causal analyses based on those evidence of polygenic association with AD case/control status.

ii. **Alzheimer's Disease:** Kunkle et al. (3).
N = 94,437 individuals of European decent.
Cases include clinically diagnosed late-onset (>65 years) AD only.

iii. **Education:** Lee et al. (4).
N = 1.1million individuals of European decent.
Attainment = highest level of education at age 30.

iv. **Cognition:** Savage et al. (5).
N = 269,867 individuals of European decent.
Cognition was measured using a test battery. Scores were combined into a common latent cognition ('g') factor.
Cohorts ranged from children to older adults (range=5-98 years), with older adults screened for cognitive decline to exclude prodromal dementia.

To the best of the authors knowledge, no sample overlap existed across studies.

### 3. Selection of Genetic variants

#### a. GWAS selection (including p-value thresholding / clumping)

GWAS summary statistics were utilised to select instrumental variables using a genome-wide approach. As in our previous MR study (6), this approach was favoured over that of a candidate gene-region(s) strategy due to the polygenic nature of all phenotypes of interest. With the exception of APOE, per-SNP effect sizes remain small, making a pooled IV approach most appropriate.

To ensure independence amongst instrumental variables, instruments were clumped using an r2 threshold of 0.001 for AD, education, and cognition exposures. This was not required for metabolites as instruments were selected using a list of pre-curated metabolite quantitative trait loci (mQTLs) extracted from Kettunen et al (2016) (readily available within the MR-Base catalogue).

#### b. Exclusion criteria

Univariable analyses:

i. SNPs with an F statistic <10

Univariable and mediation analyses:

ii. Exposures with cross-trait PRS p>0.05 (metabolites only)
iii. SNPs significant with both exposure and mediator at $p>5*10^{-08}$
iv. SNPs in linkage disequilibrium >r2=0.001
v. SNPs associated with the outcome at $p<5*10^{-08}$
vi. SNPs with known pleiotropy (ApoE)

Mediation analyses:

vii. Exposures with no significant univariable association with mediator
viii. Mediator(s) with no significant univariable association with outcome

#### c. Assessment of instrument validity

i. Assessment of per-instrument F statistic
ii. Sensitivity analyses: leave-one-out, MR-Egger, weighted median, Cochran's Q.

### 4. Harmonization procedure

For all MR analyses, inferable SNPs were aligned across the exposure and outcome datasets, and any non-inferable palindromic SNPs were removed.

### 5. Primary analyses: multiple testing

Multiple testing was corrected for using a Bonferroni corrected adjustment which takes into consideration correlation amongst variables. The procedure was as follows:

i. A metabolite*metabolite genetic correlation matrix was computed using LD score regression (7).
ii. The correlation matrix was passed through an "independent_tests" package within Python (https://github.com/hagax8/independent_tests) to compute the number of independent metabolites, given their correlation structure (Supplementary Information S3).
iii. The number of independent tests (*T*) was computed as the number of independent metabolites plus additional tests for education and cognition.
iv. A final Bonferroni corrected alpha was then computed by dividing *T* by 0.05.

### 6. Sensitivity analyses

A number of sensitivity analyses were conducted to scrutinise the validity of primary results:

**a.** For univariable analyses:

i. Robust methods: MR-Egger and weighted median
ii. Cochran's Q-statistic
iii. Bayesian Model Averaging MR

**b.** For multivariable analyses:

i. MR-Egger

### 7. Data presentation

All MR output is provided either within the main manuscript or supplementary material for the scrutiny of readers. Results which are statistically significant as well as those which do not reach significance are reported across all analyses in order to provide complete transparency and avoid reporting bias. Charting visualisations for primary results as well as for sensitivity measures such as leave-one-out, Egger and weighted median are provided for maximum transparency and interpretability of results. All results are also available in tabular form for use in further analyses where required.

*Interpretation of results can be found within the results and discussion section of the main manuscript.*

**Supplementary Information S3**

1. **ADNI**

ADNI is a USA based longitudinal initiative, beginning in 2004 and following participants through multiple study phases. Currently in the 3rd iteration, genotype data was available for the first two, typed across three separate genotype chips: ADNI1 (Illumina Human610-Quad BeadChip), ADNI2 (Illumina HumanOmniExpress BeadChip), and OMNI (a combination of phase 1 and 2 participants typed on a high coverage Illumina chip - Omni 2.5M). ADNI2 and OMNI were aligned to the Genome Reference Consortium genome build 37 (GRCh37), whilst ADNI1 was aligned to an older build (GRCh36) requiring lifting over to GRCh37. Pre-QCd data across the three genotype chips (overlaps excluded) consisted of 1,674 AD, mildly cognitively impaired (MCI), and healthy individuals.

2. **ANM-DCR**

The AddNeuroMed and Dementia Case Register is a European based study, initiated in 2008 with the aim to establish biomarkers for early AD prediction. DCR is a follow-up of ANM, with UK subjects recruited from the Maudsley and King's Healthcare Partners Dementia Case Register (8). In addition to full clinical and demographic data, genotype data were available for 1063 samples pre-QC. Of these, 397=AD, 235=MCI, 414=Controls (17=Other or unknown). 442=males, 616=females, 5=unknown. Median age=79years (Q1=73, Q3=83). As with ADNI1, ANM was typed on the Illumina Human610-Quad BeadChip.

3. **GERAD1**

Data used in the preparation of this article was obtained from the Genetic and Environmental Risk for Alzheimer's disease (GERAD1) Consortium (Harold et al. 2009). A number of biological, psychosocial, and cognitive variables have been collected for this cohort, with our study having access to genotype, age, sex, and APOE information only. Samples were genotyped at the Sanger Institute on the Illumina 610-quad chip. These samples were recruited by the Medical Research Council (MRC) Genetic Resource for AD (Cardiff University; Kings College London; Cambridge University; Trinity College Dublin), the Alzheimer's Research UK (ARUK) Collaboration (University of Nottingham; University of Manchester; University of Southampton; University of Bristol; Queen's University Belfast; the Oxford Project to Investigate Memory and Ageing (OPTIMA), Oxford University); Washington University, St Louis, United States; MRC PRION Unit, University College London; London and the South East Region AD project (LASER-AD), University College London; Competence Network of Dementia (CND) and Department of Psychiatry, University of Bonn, Germany and the National Institute of Mental Health (NIMH) AD Genetics Initiative. All AD cases met criteria for either probable (NINCDS-ADRDA, DSM-IV) or definite (CERAD) AD. All elderly controls were screened for dementia using the MMSE or ADAS-cog, were determined to be free from dementia at neuropathological examination or had a Braak score of 2.5 or lower. Prior to quality control, 3,292 cases and 1,223 controls were available (1,645=males, 2,870=female). Median age across samples was 80 years (Q1=68,Q3=80).

### Supplementary Information S4

Pseudo-R2 statistics for PRS analyses were retrospectively computed using meta-analysed regression coefficients by computing the meta-analysed z-statistic, and applying the following equation: $R^2 = (z/\sqrt{(n - 2 + z^2)})^2$; where $n$ = the sample size.

### Supplementary Information S5

Aided by an “independent_tests” package in Python (https://github.com/hagax8/independent_tests), together with a squared genetic correlation matrix (computed using LD score regression (7)) for Kettunen et al. (2) metabolites analysed in PRS analyses, the number of independent tests and adjusted alpha were calculated as follows:

1. The number of principal components explaining 99.5% of the variance *(N)* within the squared correlated matrix was computed *(N=28).*
2. *N* of individual tests (*T*) within the squared correlation matrix was then calculated using formula: *T=(N*N-N)/2 (T=378).*
3. The square root of T was then used to establish the final number of individual metabolites (*m*) to correct for ($m=\sqrt{T}$ (m=*19.4*)).
4. *M* multiplied by 10 to account for the number of p-value thresholds (*pt*) run for each metabolite in cross-trait PRS analyses against metabolites *(f=m*pt)* (*f*=194).
5. An adjusted alpha ($\alpha$) was then computed by dividing 0.05 by *f* ($\alpha$=0.0002).

Computed alpha for MR analyses (aided by the “independent_tests” package in Python):

1. *N=11*
2. *T=55*
3. *m=7.42*.
4. *m* multiplied by two to reflect outcome phenotypes (1*AD, 1*cognitive). Note: education and intelligent counted as non-independent due to high rg. *f=14.*
5. Adjusted $\alpha$=0.004.

### Supplementary Information S6

For both univariable and multivariable analyses, causal effects were re-estimated using MR-Egger. Egger methodology mirrors IVW but with the intercept constraint removed. As the IVW constraint reflects the assumption of no independent effect of G on Y, an intercept value which significantly deviates from 0 is indicative of pleiotropy (9). Large discrepancies between Egger and IVW estimates are also warning signs for pleiotropy, though wider confidence intervals for Egger are to be expected (1). For univariable analyses, causal estimates were also re-estimated using weighted median. Like Egger, this method provides an addition MR estimate in the presence of pleiotropy. Here, per-instrument estimates are ordered and weighted by their causal association, with the median then taken as the causal estimate (10). Influential points were investigated using leave-one-out. For this, per-SNP instruments are removed from models one at a time and causal effects re-estimated to identify single SNP instruments which may be driving causal estimates. Cochran's Q also tested for heterogeneity amongst instruments, indicated by Q-$p<0.05$ (1).

For metabolite-AD models only, Bayesian model averaging MR (MR-BMA) was also employed to re-assess causal estimates whilst accounting for known correlation amongst metabolites. Full details of MR-BMA can be found elsewhere (11). Briefly, metabolites genetically correlated (*rg*) up to a threshold of 0.95 (estimated using LD score regression (6,7) (Supplementary Dataset S6, Supplementary Information S6) were included within a single MR-BMA model. Marginal inclusion probabilities (MIP) for each metabolite were then computed, representing the posterior probability (*pp*) of metabolite *j* appearing within the true causal model given 10,000 iterations (prior probability=0.1, prior variance (σ2)=0.25). A model averaged causal effect (MACE) was also estimated, representing the estimated direct effect of metabolite *j* on outcome *y*, averaged across each of these *pp*s. Metabolites showing the strongest evidence for causation in MR-BMA were expected to corroborate those metabolites identified within univariable analyses.

**Supplementary Information S7**

Bayesian Model Averaging MR (BMA-MR) adopts a multivariable framework, whereby multiple exposures can be included within the model, provided a) they are each robustly associated with a least one SNP-instrument used within the model, and b) they do not induce multi-collinearity. To meet criterion b), pairwise genetic correlations (rg) were computed for metabolites using LD Score regressions (LDSC) (7). Metabolites with rg>0.95 were then pruned according to the following procedure:

i) For metabolite pairs with *rg*>0.95, if one metabolite demonstrated a greater number of *rg*>0.95 with other metabolites, that metabolite was removed.
ii) For metabolite pairs with *rg*>0.95 *and* an equal number of *rg*>0.95 with other metabolites, the metabolite with the greater number of correlations at adjusted significance was removed.
iii) For metabolite pairs with *rg*>0.95 and which also had an equal number of metabolite pairwise *rg*>0.95 at adjusted significance, the metabolite with the greatest number of nominally significant *rg*s (*p*<0.05) was removed.

**Supplementary Information S8**

*Post-hoc analyses: APOE related polygenic association*

For all primary analyses, SNPs within the APOE genomic region were removed. For PRS, APOE was removed due to its unusually large effect size for both Alzheimer's Disease (3) and lipid-related metabolites (2), risking dominating and drowning out wider polygenic signal. For lipid-related metabolites associated with AD at $p$<0.05 in PRS analyses however, results were in the unexpected direction, with positive AD-associations observed for HDLs, and negative AD-associations observed for LDLs, IDLs, and VLDLs (Figure 3 (main manuscript), Supplementary Dataset S7). This result runs counter to the direction of associations observed for these lipid subfractions within the wider literature (6,12–15). To investigate whether this reflected APOE removal, PRS meta-analyses were thus re-computed for all lipid-related metabolites significantly associated with AD at $p$<0.05 in primary analyses, but with SNPs restricted to the APOE genomic region only (chr19, bp 45786555-45037368). The QC pipeline followed that outlined for primary analyses in Fig. 1 of the main manuscript, with the exception of "APOE region SNPs removed". PRS analyses were then re-computed, restricting p-value thresholds to those identified as each metabolite's best threshold within primary analyses to ensure a like-for like comparison of results (Supplementary Dataset S7).

Across all post-hoc APOE-region PRS analyses, the direction of association between each metabolite and AD matched that which would be expected for these metabolites based on wider literature, with a negative association observed for XL.HDL.FC, and positive associations for LDLs, VLDLs, and IDLs. Associations were also highly significant, ranging from p=0.001 (XL.HDL.FC) to $p=2 \times 10^{-59}$ (S.VLDL.FC) (Supplementary Dataset S24). This indicates that primary PRS results may in-part reflect the removal of APOE related SNPs. This removal was necessary due to the risk of APOE dominating and drowning out wider polygenic signal as a result of the unusually large effect size that this genetic region has. However, these post-hoc analyses highlight the importance of consideration for how such removal may impact conclusions made from the data if not carefully thought through. Evidence from our results here suggest that because the genetic signal from APOE largely dominates the genetic relationship between lipid-subfractions and AD, its removal in polygenic score analyses may bring to bear counterintuitive associations which work in the opposite direction to that expected based on wider associations which do not consider the effect of APOE. This may be particularly pertinent for impacting polygenic score analyses, as opposed to MR, due to the explicit assumptions of polygenic scores relating to polygenicity which can result in signal bias when these assumptions do not hold due to the presence of unconventionally large effects (see (16). Indeed, this may explain why this phenomenon appeared to impact PRS results, whilst MR maintained consistency despite APOE removal. Whilst outside the scope of the current study, it will be interesting to expand on the genetic associations observed here and further investigate the polygenic relationship between these metabolites and AD on the basis of, and interactions with, APOE status.

**Supplementary Information S9**

***MR-Egger***
Both univariable and multivariable models were re-estimated using MR-Egger – a conservative estimate which is sensitive to pleiotropy. The causal effect of both glutamine and XL.HDL.FC on AD retained consistent directionality. However, lower precision of the Egger estimate resulted in wider confidence intervals equating to a loss of significance for both of these metabolites (Supplementary Dataset S9. Fig. 4a). This was also the case for all education-metabolite and cognition-metabolite estimates of at least nominal significance (Supplementary Dataset S11,S13), and for the causal effect of both education and cognition on AD (Supplementary Dataset S15). Direction of effects did, however, corroborate across primary and Egger estimates for both education and cognition on AD, and for all but one (FAw3) of the causal associations between education and metabolites (Supplementary Dataset S13, S15). The effect of cognition on metabolites was more problematic however, with non-corroborated directionality for seventeen of the eighteen nominally significant metabolites (Supplementary Dataset S11). The Egger intercept – an additional test of pleiotropy – indicated no significant deviations from 0, however, indicating no substantial pleiotropy.

***Weighted Median***
Weighted median was conducted as an additional measure robust to pleiotropy. As with MR-egger, estimates for nominally significant results were attenuated for weighted median, though all estimates remained in the agreed direction (Supplementary Dataset S9,S11,S13-S15). For the effects of glutamine, XL.HDL.FC, and education on AD, adjusted significance was also retained (Supplementary Dataset S9, S15. Fig. 4a.). This was also the case for bidirectional estimates between education and cognition (Supplementary Dataset S17).

***Cochran's Q and Leave-one-out***
Significant evidence of heterogeneity was observed in bi-directional tests between education and cognition (*Q-pval*<0.05. Supplementary Dataset S17). Heterogeneity was also observed for the effect of education on FAw3 (*Q*-pval=0.013), and for a number causal estimates for cognition on metabolites (Supplementary Dataset S11). No heterogeneity was evident for either Glutamine or XL.HDL.FC on AD (Supplementary Dataset S9), nor for education on AD (Supplementary Dataset S15). There was, however, significant heterogeneity when investigating the effect of cognition on AD (*Q-pval*=0.01) (Supplementary Dataset S15).

Leave-one-out identified one influential instrument for glutamine (rs2657879). Instrument removal resulted in attenuation of the causal estimate though directionality remained consistent (OR=0.82, 95% CI=0.62-1.10. Supplementary Figure S12). Removal of rs1532085 resulted in the largest attenuation for XL.HDL.FC on AD (Supplementary Figure S13), though nominal significance was retained (OR=0.87, 95% CI=0.76-0.98). No notable outliers were observed for either education of cognition (Supplementary Figure S14-S17).

***Bayesian Model Averaging***
Following the pruning of metabolites with genetic correlations >95%, nineteen metabolites were jointly analyzed (Supplementary Dataset S6). Results corroborated

primary analyses, with XL.HDL.FC and glutamine ranked with the highest MIP; indicative of being the strongest “true causal” candidates of those analysed (Supplementary Dataset S19). MACE, reflecting the average direct causal effect was also in agreement with primary analyses, with a negative (protective) relationship estimated for both metabolites (XL.HDL.FC: *MACE*=-0.043, Glutamine: *MACE*=-0.045). When metabolite combinations were assessed, glutamine and XL.HDL.FC were also the most frequent metabolites to appear within the top causal models (Supplementary Dataset S20).

**Supplementary Information S10**

A number of causal associations significant at $p<0.05$ were observed when investigating causal relationships between blood metabolites and either education or cognition. Whilst caution should be taken in the interpretation of such results due to their failure to retain significance when accounting for multiple testing, they nonetheless warrant brief discussion as they may represent relationships which hold promise in future replications. Firstly, a bidirectional causal relationship in the positive direction was observed between Omega3 fatty acids (FAw3) and education. This was stronger in the direction of education increasing levels of FAw3, rather than the other way around. However, directionality of point estimates for sensitivity measures were more consistent in the direction of FAw3 increasing education, with less heterogeneity amongst instruments also present. A positive association between FAw3 and education aligns with some of the wider literature, though results from randomised control trials remain conflicting or inconclusive (7,17–20). It has also been implicated in lower inflammation (21–23), increased HDL levels (24,25), and decreased triglyceride levels (25–28); all of which consistently associate with neurocognitive protective outcomes. A negative causal effect of education on eight additional lipid-related metabolites was also observed, indicating that higher education leads to reduced levels of these metabolites. These all belonged to the IDL or LDL family of metabolites which are consistently implicated in poor vascular outcomes (29–32), though their specific role in neurocognitive outcomes remains conflicting (33–37). Similarly, levels of eighteen LDL, IDL, and cholesterol-related metabolite sub-fractions also demonstrated a reduction when cognition levels increased. As with IDL.TG, however, there was no convincing evidence that effects translated through to AD. Extra caution is also warranted in the interpretation associations observed between cognition and these metabolites, as seventeen of the eighteen did not show consistency in effect direction when estimated using MR-Egger, and in many cases, significant instrument heterogeneity was evident, casting further doubt on the validity of these causal estimates. With the exception of FAw3, all associated metabolites heavily associate with APOE (2) and with only five instruments available for FAw3, attenuated signal remains a very real possibility. As with IDL.TG, larger replications, when available, will be an important future exercise. However, on the basis of results observed here, no metabolites were observed to underly the relationship between either education or cognition and AD.

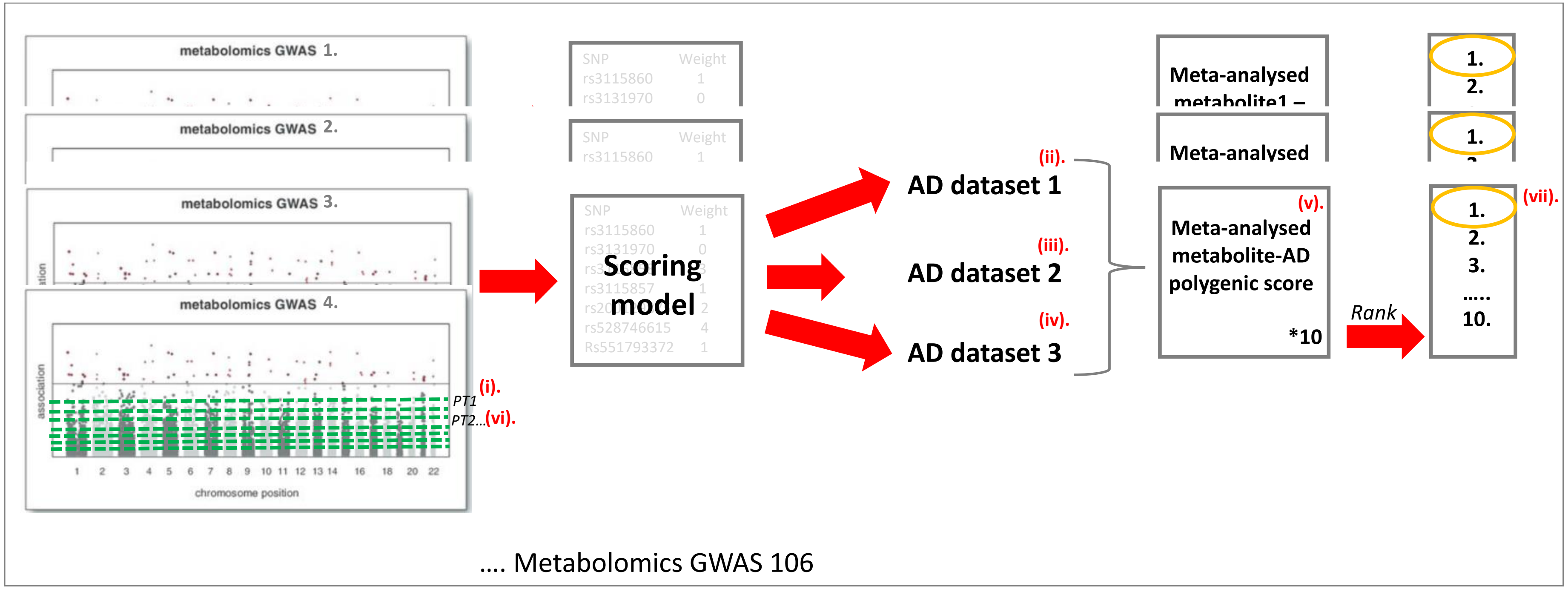


**Supplementary Figure S1. Illustrative overview of PRS analytical pipeline**. 1 metabolomic GWAS dataset at a time: (i) data is cut at the first of the ten pre-defined p-value thresholds. Any SNP with a p-value lower than the pre-defined threshold is included within the scoring model for metabolite[n] at threshold[k]. (ii) Genotype data from AD dataset 1 is then regressed against the scoring model for metabolite[n] at threshold[k]. (iii) AD dataset 2 is then regressed against the same scoring model, and then (iv) AD dataset 3. Coefficients and standard errors across the three regressed datasets for metabolite[n] at threshold[k] are (v) meta-analysed (and back-generated R2 computed) to obtain a single cross-trait meta-analysed polygenic estimate for metabolite[n] on AD, when the data is cut at p-value threshold[k]. This pipeline is then repeated for the same metabolite at threshold[k+1] to obtain a meta-analysed estimate at the next pre-defined pvalue threshold at which the metabolomic genomic data is cut. This is repeated across 10 cycles for metabolite[n] to obtain 10 meta-analysed cross-trait prs estimates, each representing a differing level of SNP inclusiveness. (vii) The 10 models are then ranked by the most significant threshold descending, and the highest ranked estimate is taken as the representative “best” cross-trait polygenic scoring model for metabolite[n] and AD. This pipeline is then repeated across all 106 metabolomic datasets to obtain 106 meta-analysed cross-trait PRS estimates.

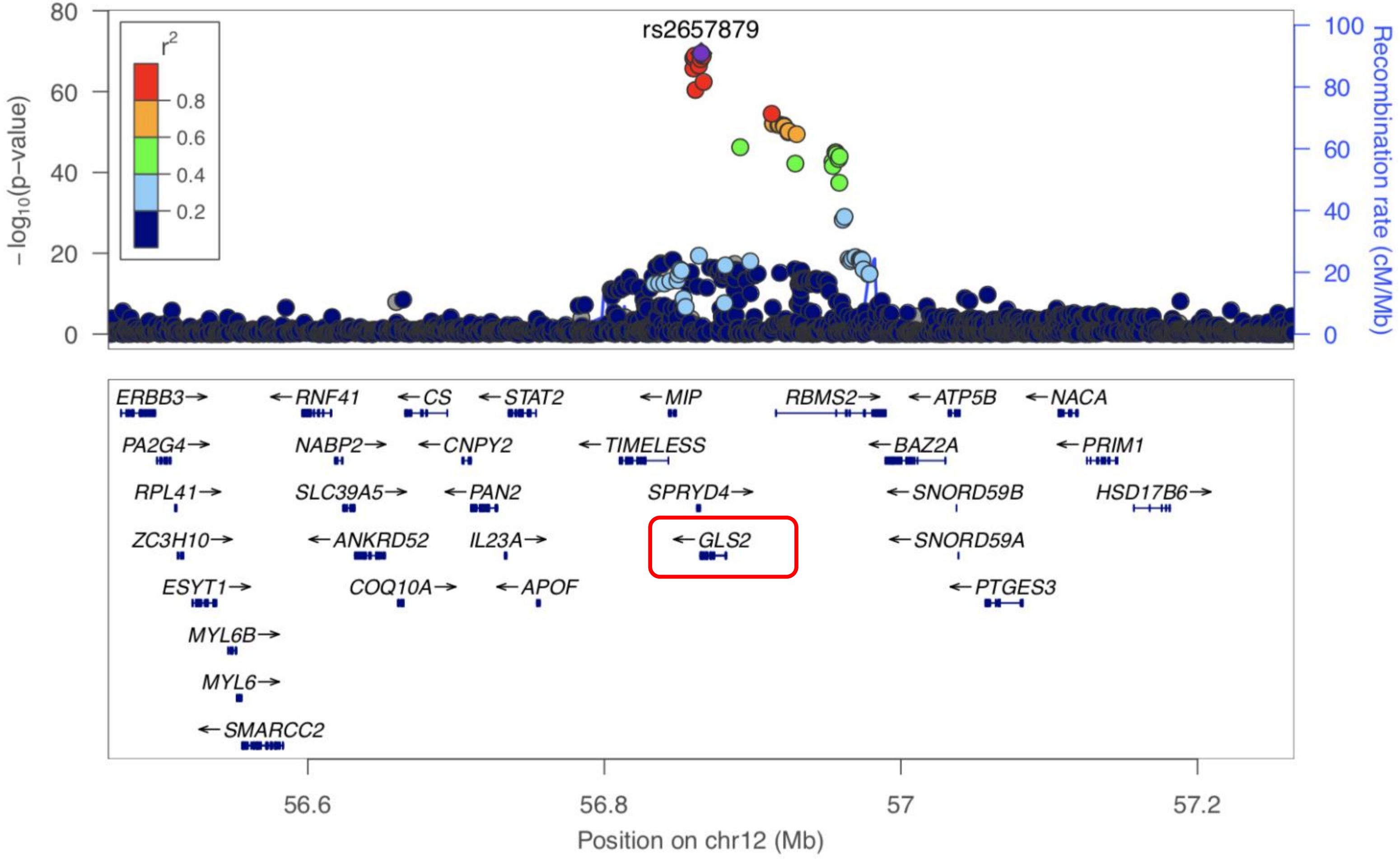


**Supplementary Figure S2.** Locus zoom plot for Glutamine influential point rs2657879. Plot demonstrates LD block which spans the GLS2 genomic region (bordered in red).

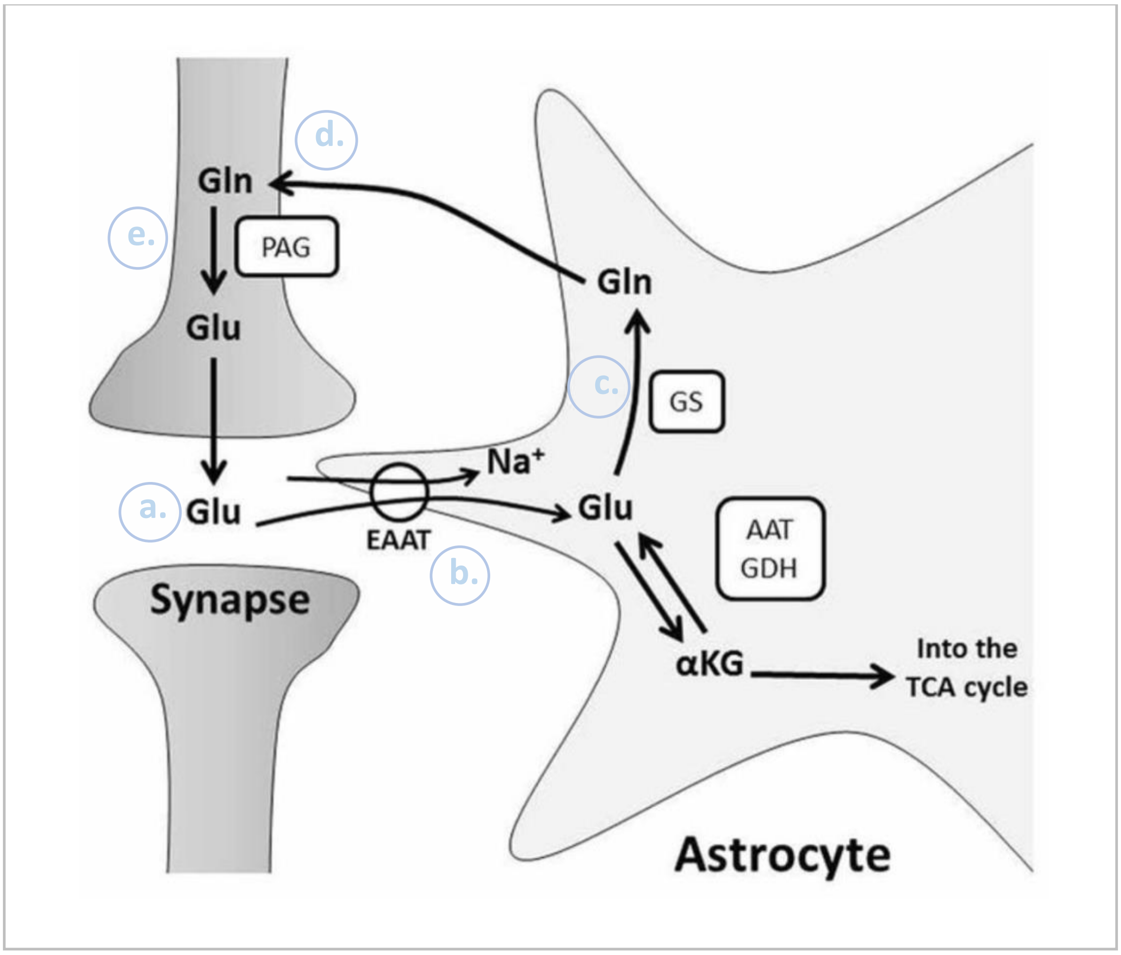


**Supplementary Figure S3 - Image obtained from Stobart & Anderson (38) and further annotated.** Diagrammatic illustration of the glutamine-glutamate cycle. During neurotransmission, glutamate (Glu) is released from the pre-synaptic cell into the synaptic cleft (a). Astrocytes re-uptake any Glue remaining within the cleft (b) and convert it to glutamine (Gln) via glutamine synthase(c). Gln is then released back into the extracellular space where it is re-uptaken by pre-synaptic cells (d) which then metabolize Gln back into Glu (e) and package into synaptic vesicles ready for the next round of neurotransmission. This cycle is critical for regulating appropriate neurotransmission within the central nervous system.

**S4a**

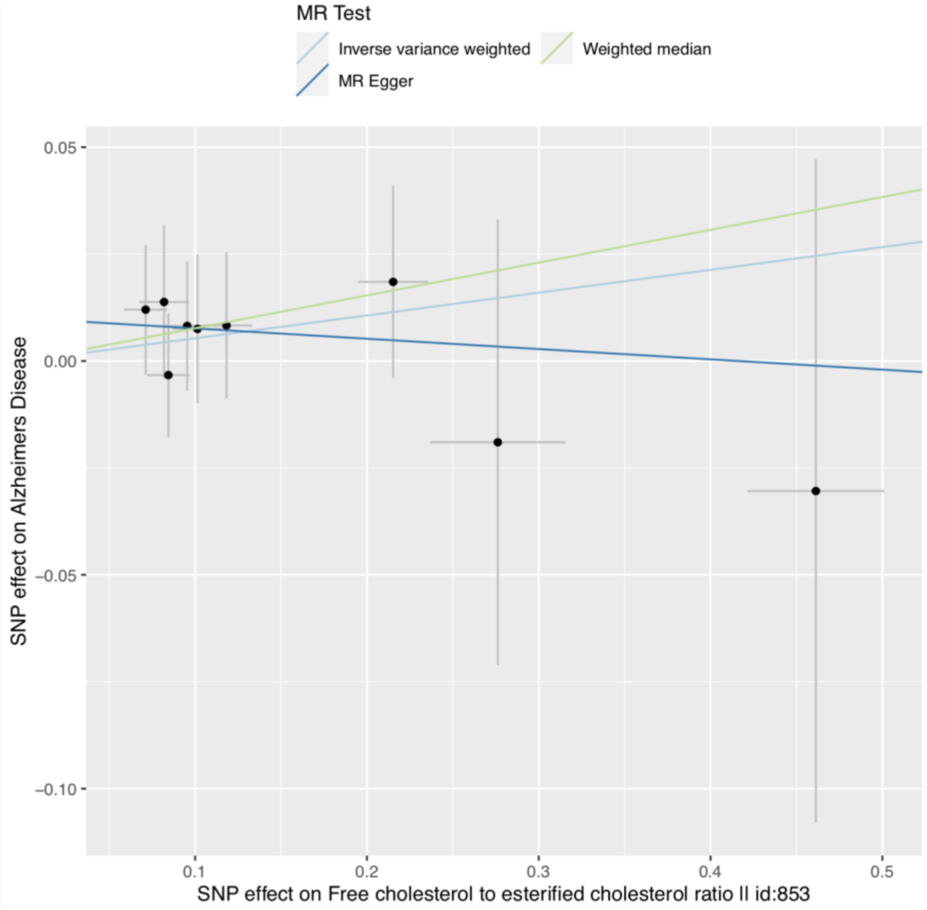
MR Test
Inverse variance weighted
Weighted median
MR Egger
0.05
0.00
−0.05
−0.10
SNP effect on Alzheimers Disease
0.1
0.2
0.3
0.4
0.5
SNP effect on Free cholesterol to esterified cholesterol ratio II id:853

**S4b**

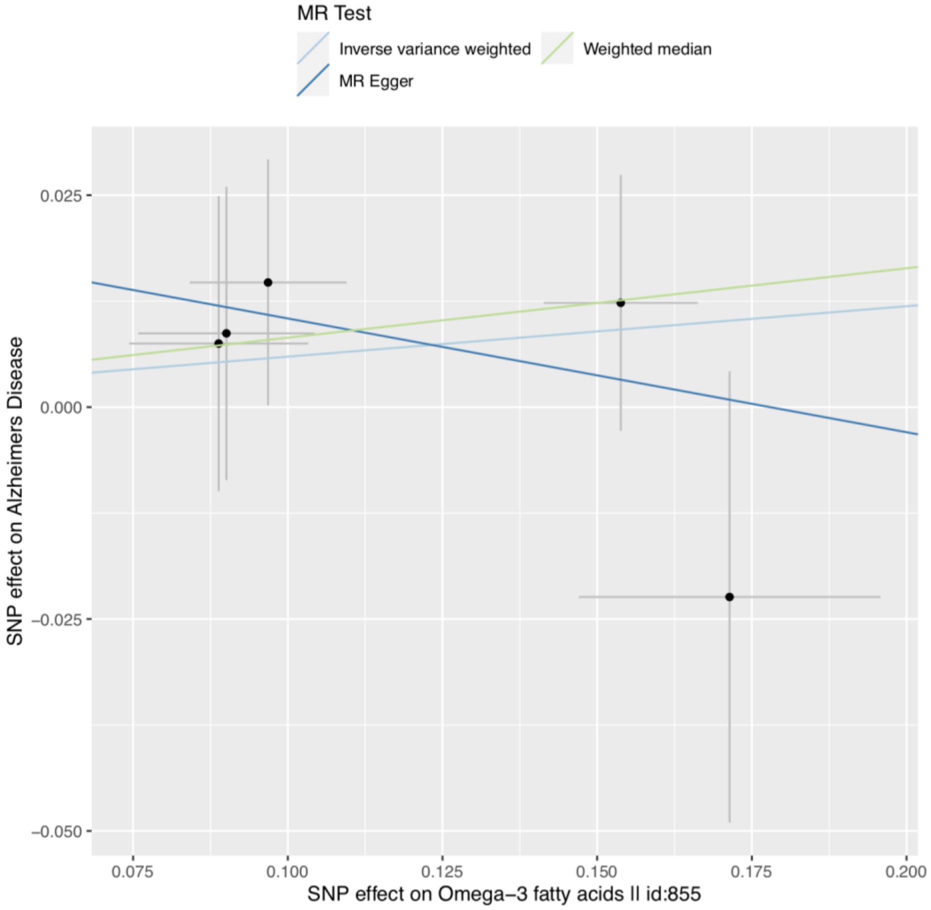
MR Test
Inverse variance weighted
Weighted median
MR Egger
0.025
0.000
−0.025
−0.050
SNP effect on Alzheimers Disease
0.075
0.100
0.125
0.150
0.175
0.200
SNP effect on Omega−3 fatty acids II id:855

**S4c**

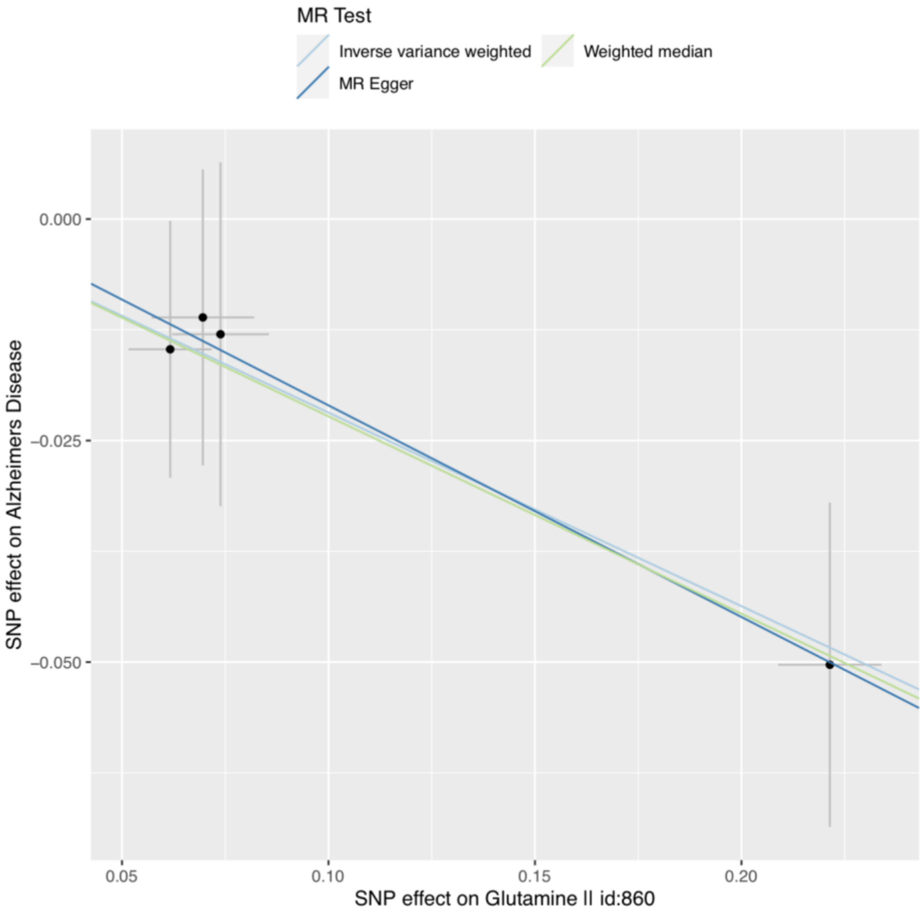
MR Test
Inverse variance weighted
Weighted median
MR Egger
0.000
−0.025
−0.050
SNP effect on Alzheimers Disease
0.05
0.10
0.15
0.20
SNP effect on Glutamine II id:860

**S4d**

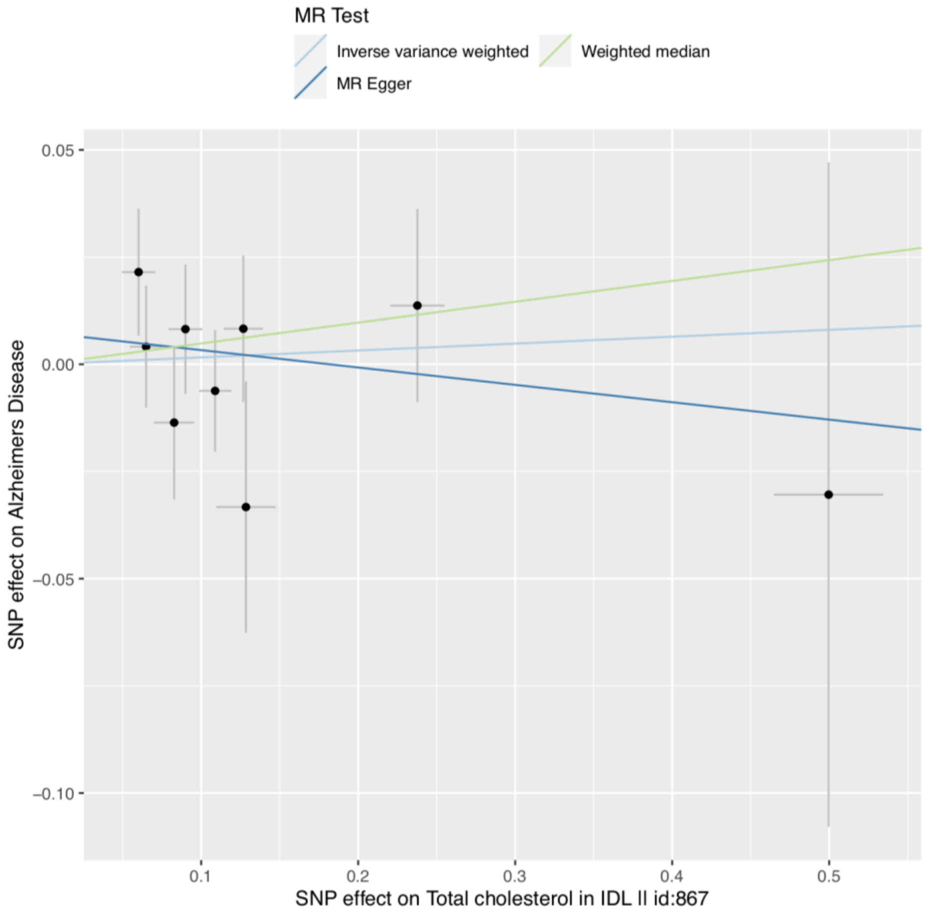
MR Test
Inverse variance weighted
Weighted median
MR Egger
0.05
0.00
−0.05
−0.10
SNP effect on Alzheimers Disease
0.1
0.2
0.3
0.4
0.5
SNP effect on Total cholesterol in IDL II id:867

**S4e**

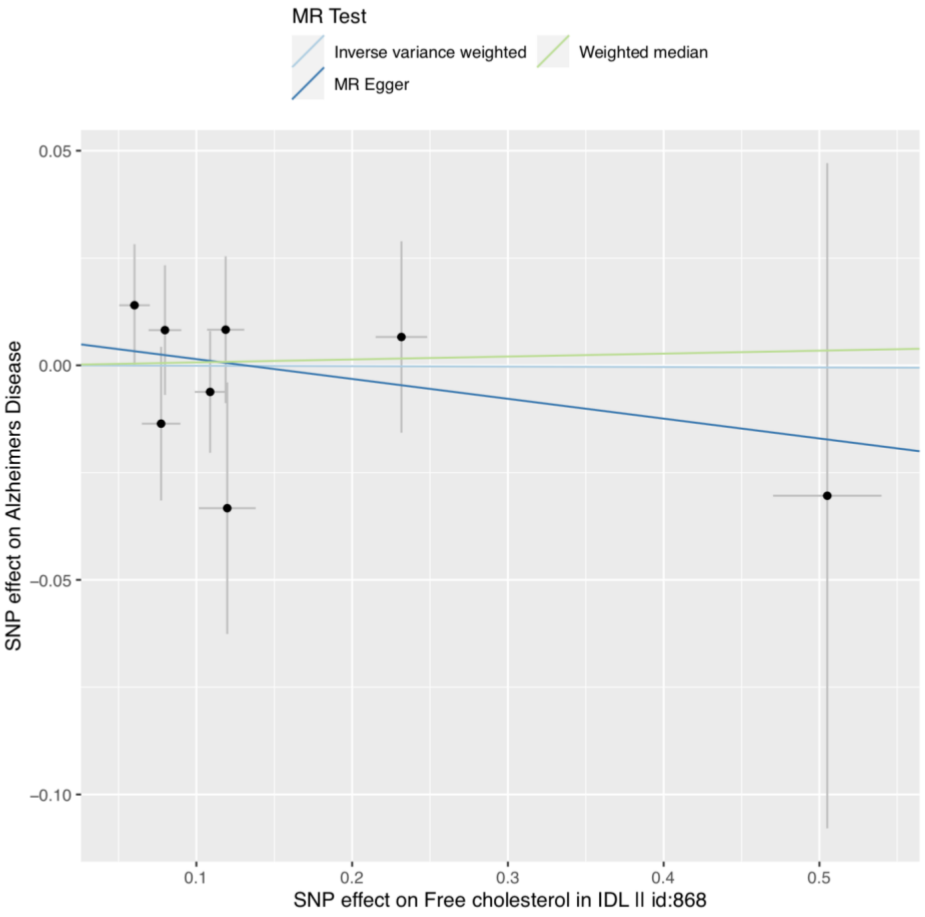
MR Test
Inverse variance weighted
Weighted median
MR Egger
0.05
0.00
−0.05
−0.10
SNP effect on Alzheimers Disease
0.1
0.2
0.3
0.4
0.5
SNP effect on Free cholesterol in IDL II id:868

**S4f**

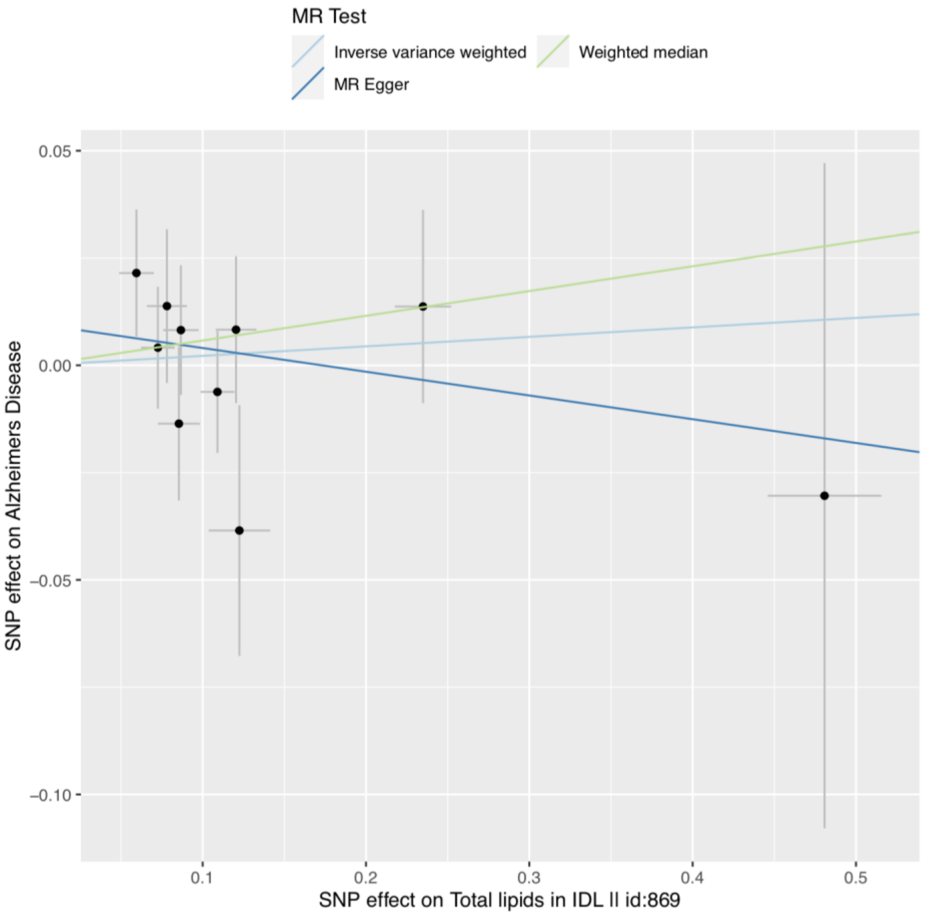
MR Test
Inverse variance weighted
Weighted median
MR Egger
0.05
0.00
−0.05
−0.10
SNP effect on Alzheimers Disease
0.1
0.2
0.3
0.4
0.5
SNP effect on Total lipids in IDL II id:869

**S4g**

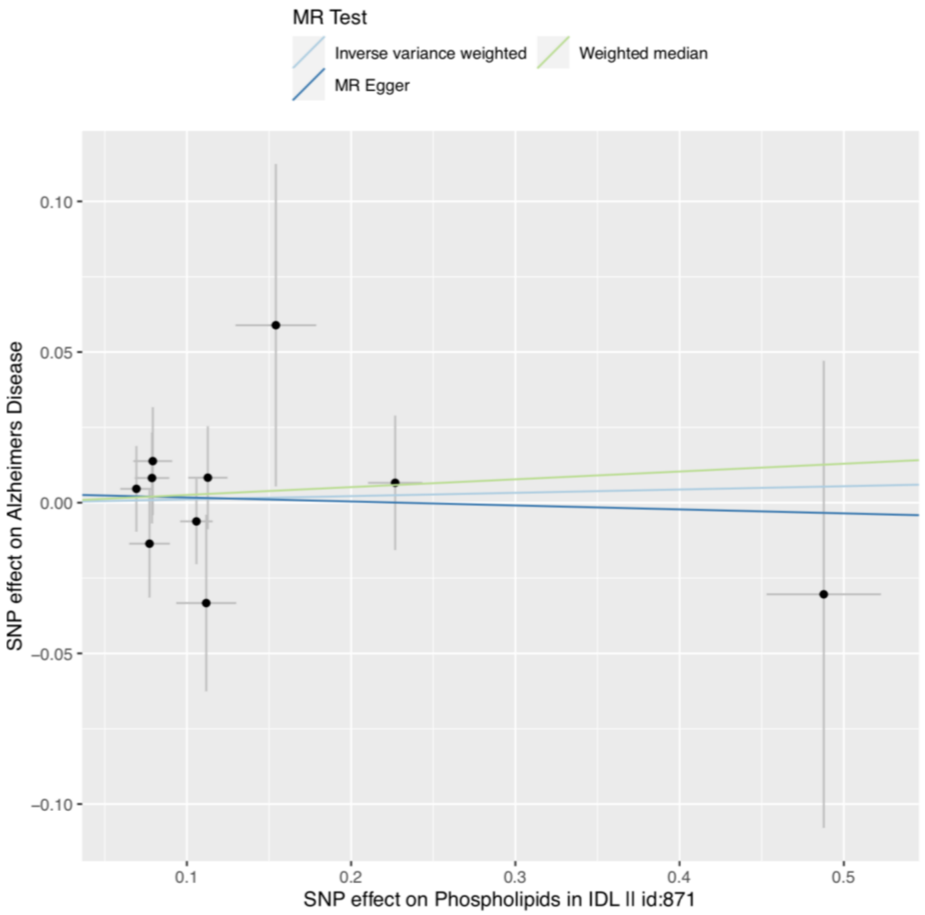
MR Test
Inverse variance weighted
Weighted median
MR Egger
0.10
0.05
0.00
−0.05
−0.10
SNP effect on Alzheimers Disease
0.1
0.2
0.3
0.4
0.5
SNP effect on Phospholipids in IDL II id:871

**S4h**

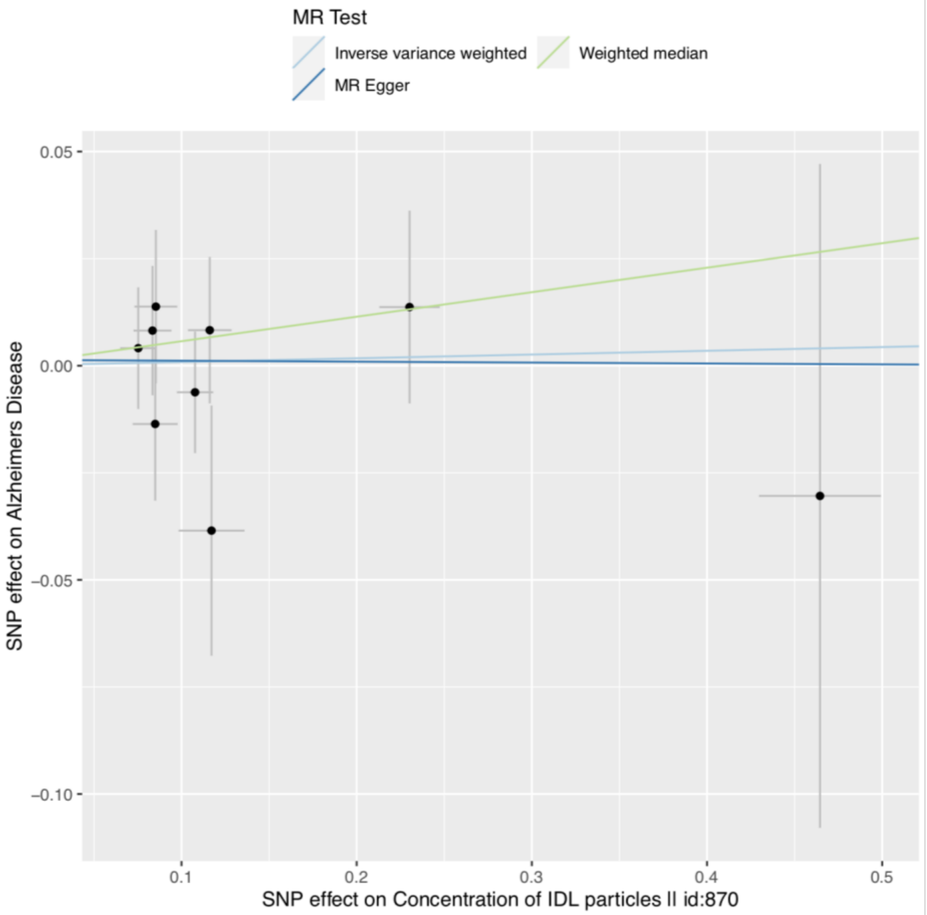
MR Test
Inverse variance weighted
Weighted median
MR Egger
0.05
0.00
−0.05
−0.10
SNP effect on Alzheimers Disease
0.1
0.2
0.3
0.4
0.5
SNP effect on Concentration of IDL particles II id:870

**S4i**

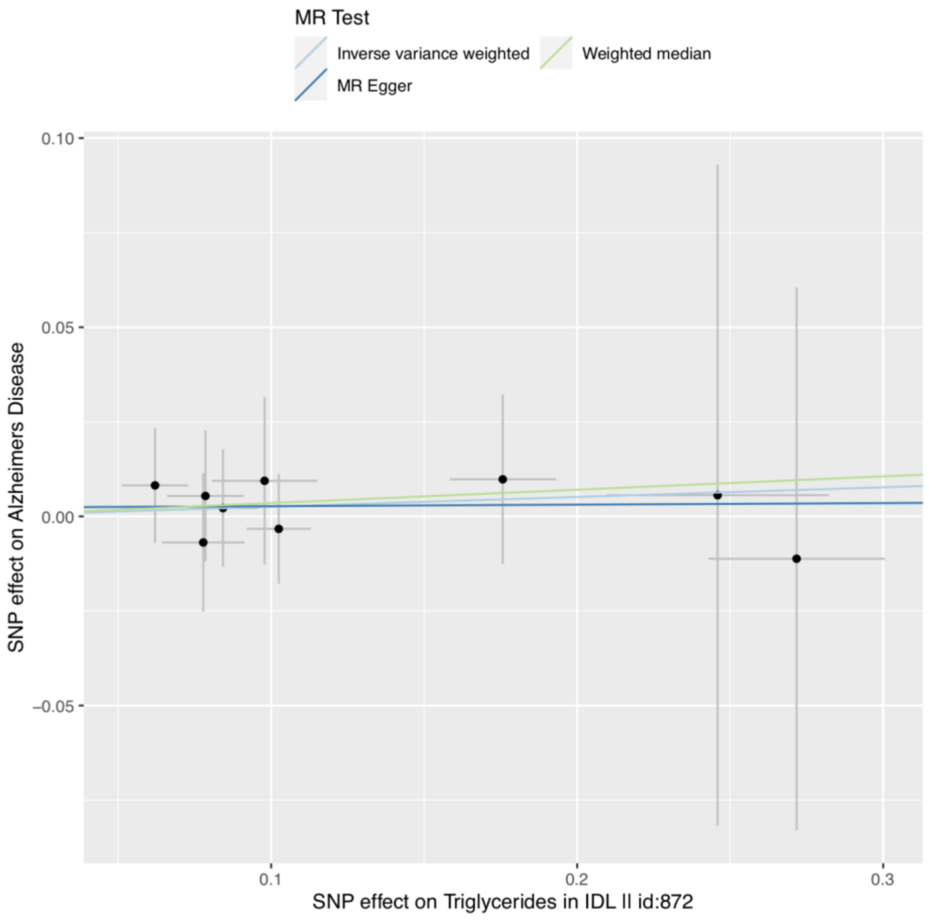


**S4j**

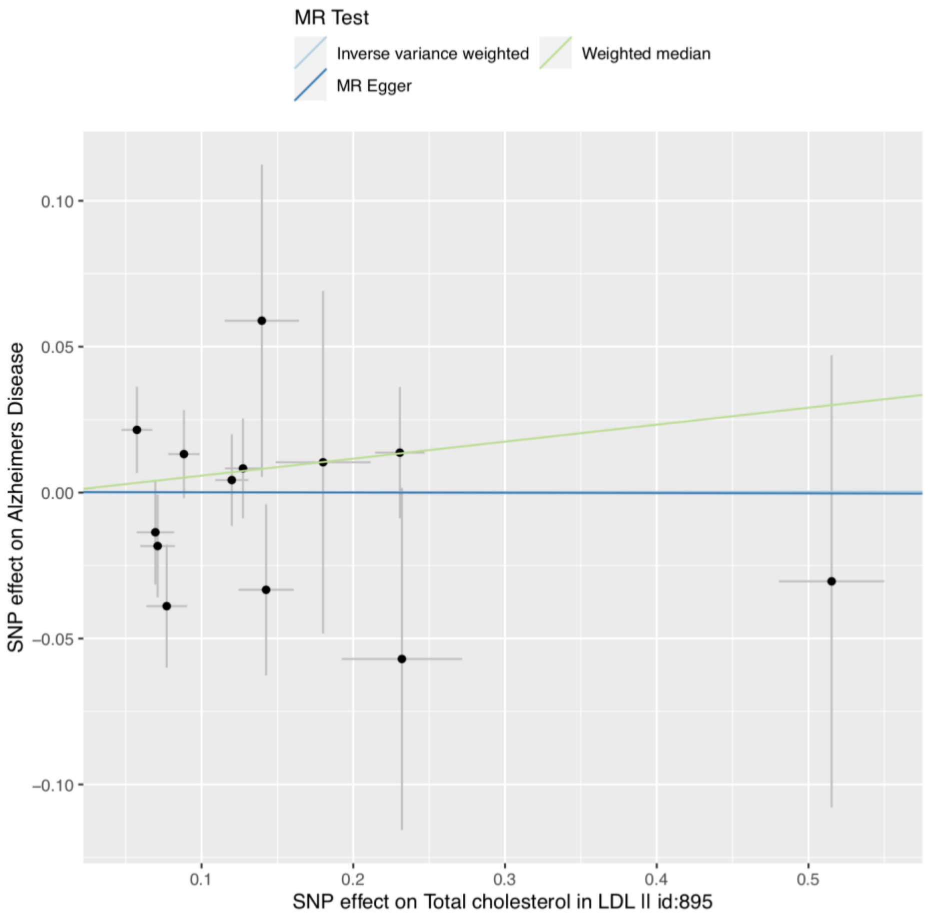


**S4k**

**S4l**

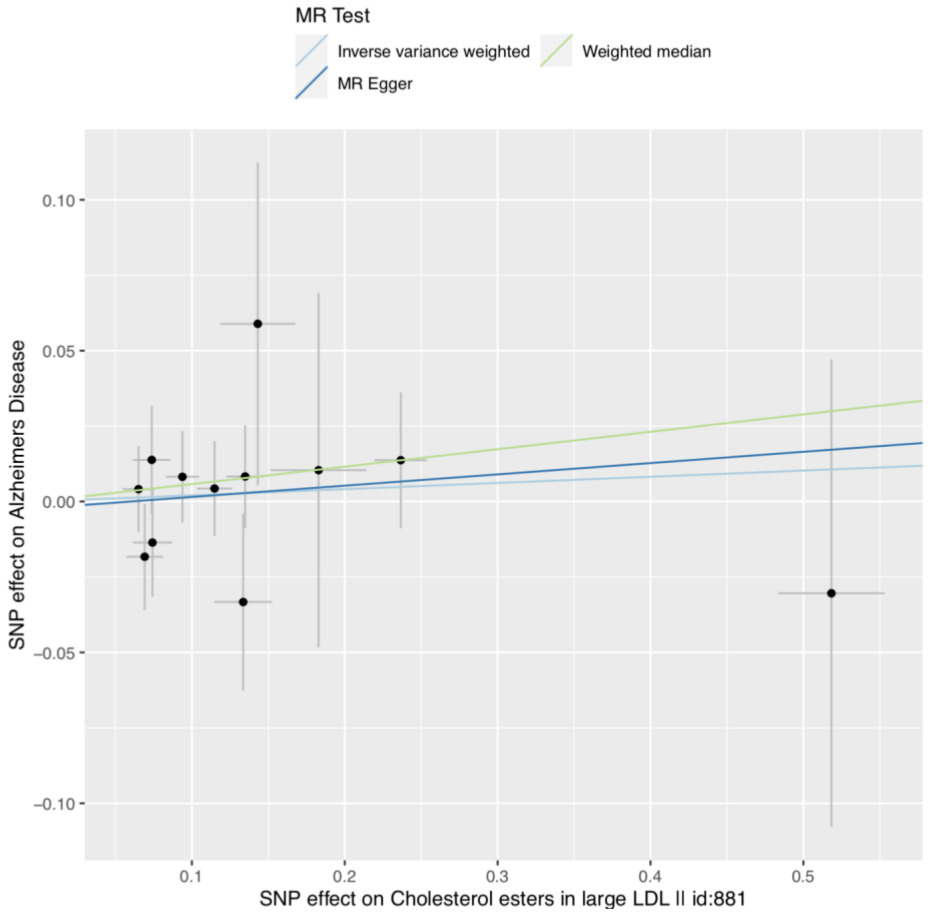
MR Test
Inverse variance weighted
Weighted median
MR Egger
0.10
0.05
0.00
−0.05
−0.10
SNP effect on Alzheimers Disease
0.1
0.2
0.3
0.4
0.5
SNP effect on Cholesterol esters in large LDL || id:881

**S4m**

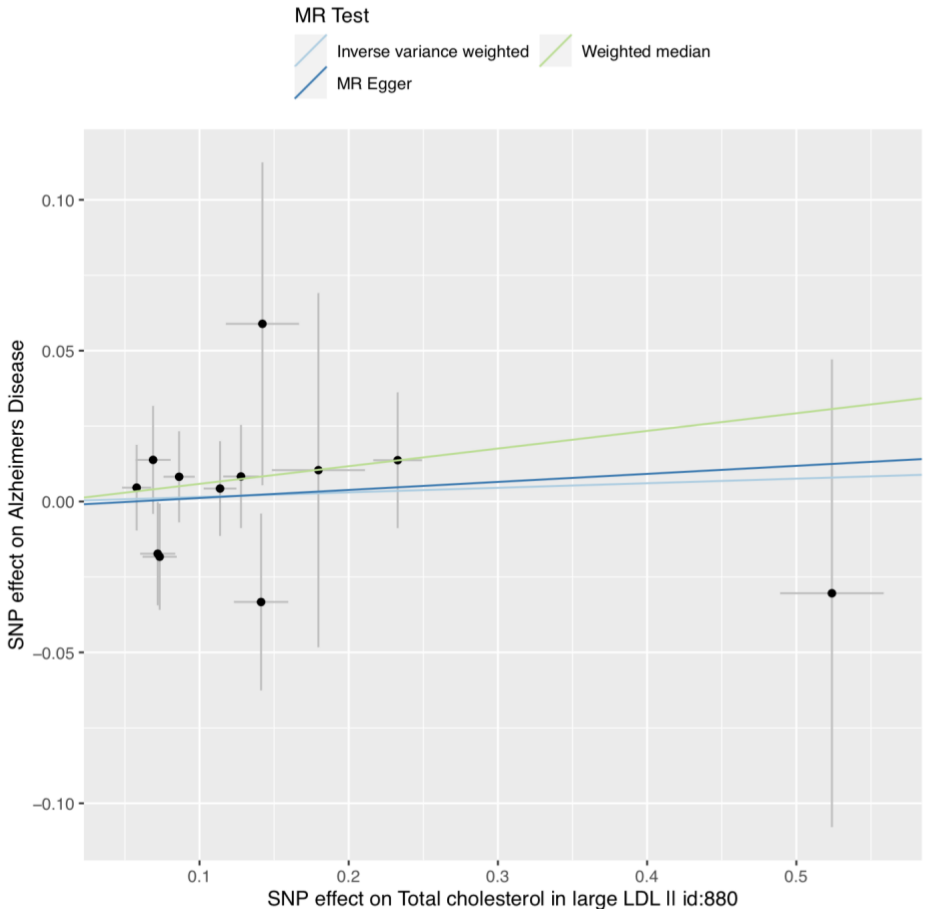
MR Test
Inverse variance weighted
Weighted median
MR Egger
0.10
0.05
0.00
−0.05
−0.10
SNP effect on Alzheimers Disease
0.1
0.2
0.3
0.4
0.5
SNP effect on Total cholesterol in large LDL || id:880

**S4n**

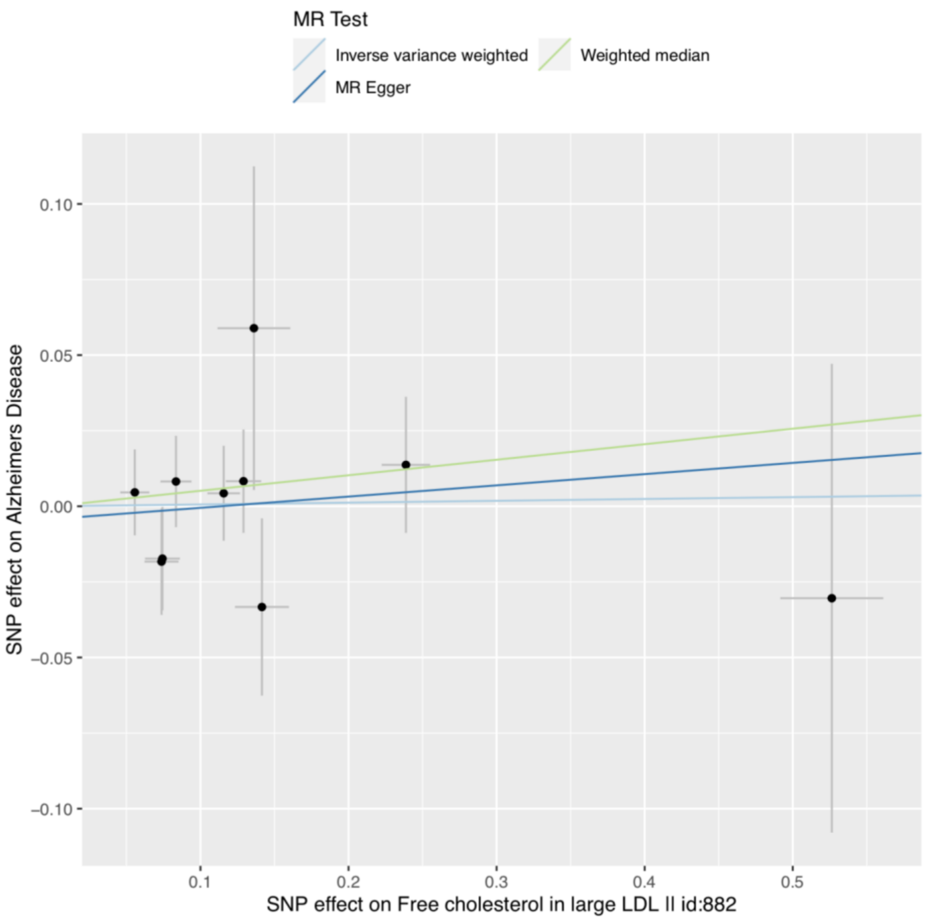
MR Test
Inverse variance weighted
Weighted median
MR Egger
0.10
0.05
0.00
−0.05
−0.10
SNP effect on Alzheimers Disease
0.1
0.2
0.3
0.4
0.5
SNP effect on Free cholesterol in large LDL ll id:882

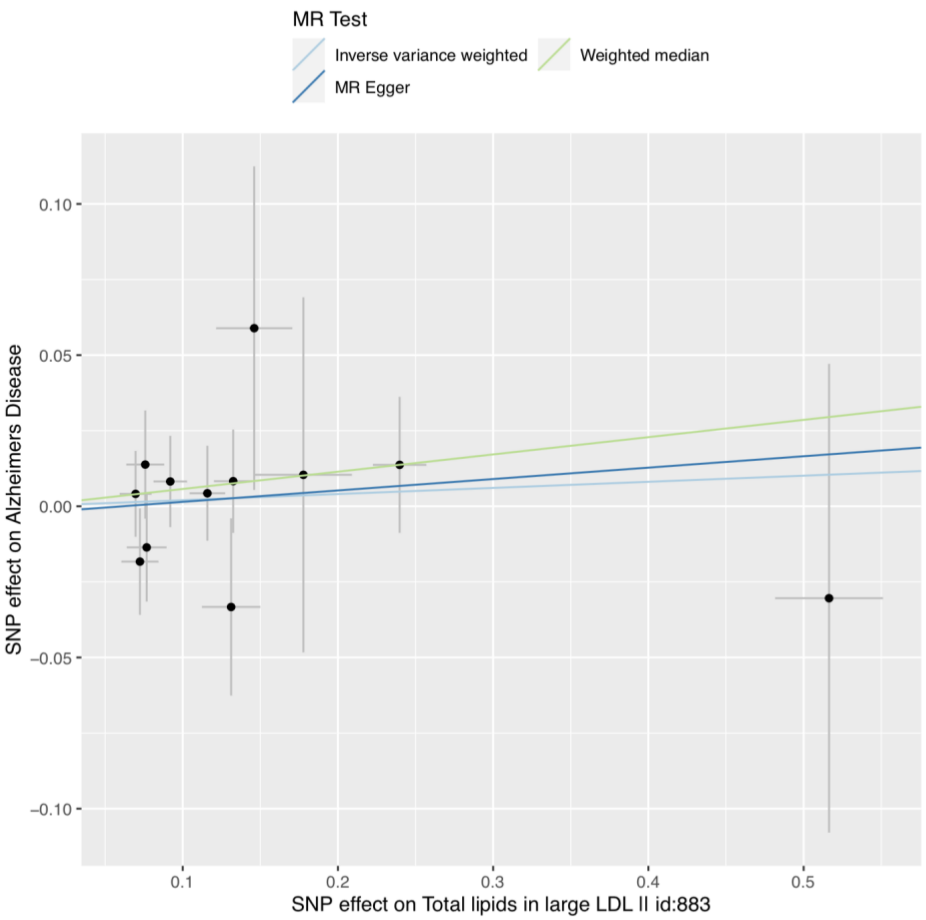
MR Test
Inverse variance weighted
Weighted median
MR Egger
0.10
0.05
0.00
−0.05
−0.10
SNP effect on Alzheimers Disease
0.1
0.2
0.3
0.4
0.5
SNP effect on Total lipids in large LDL ll id:883

**S4o**

**S4p**

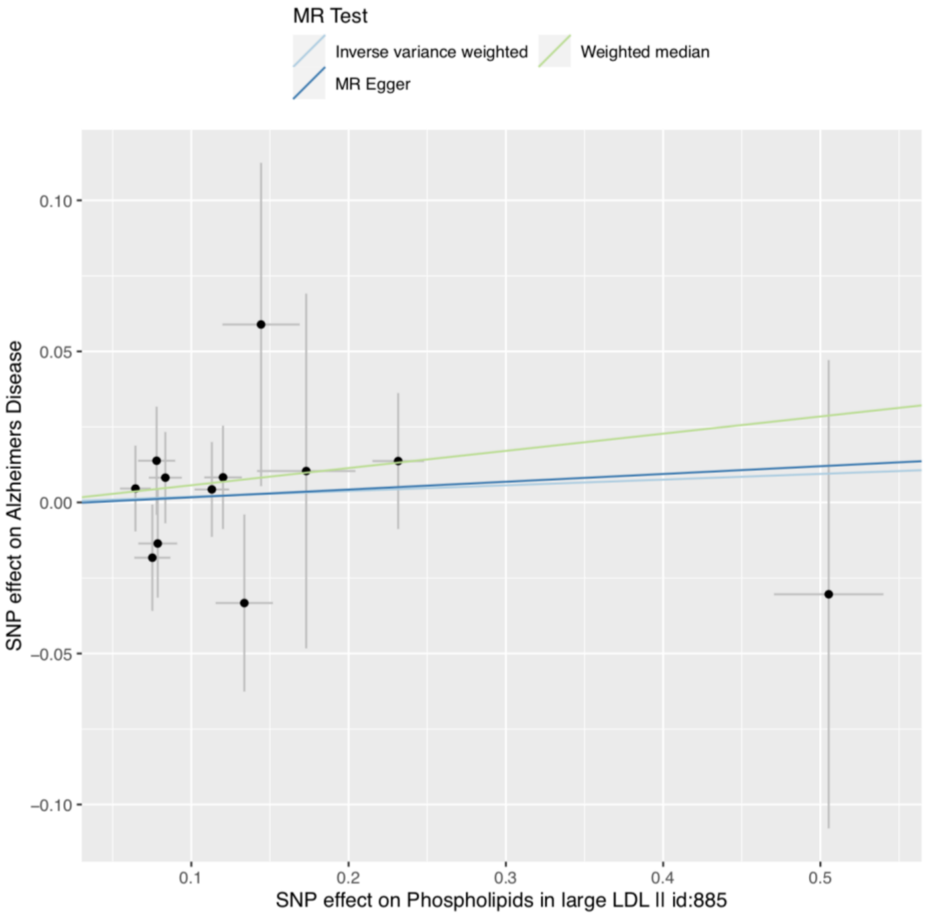
MR Test
Inverse variance weighted
Weighted median
MR Egger
SNP effect on Alzheimers Disease
SNP effect on Phospholipids in large LDL II id:885
0.10
0.05
0.00
−0.05
−0.10
0.1
0.2
0.3
0.4
0.5

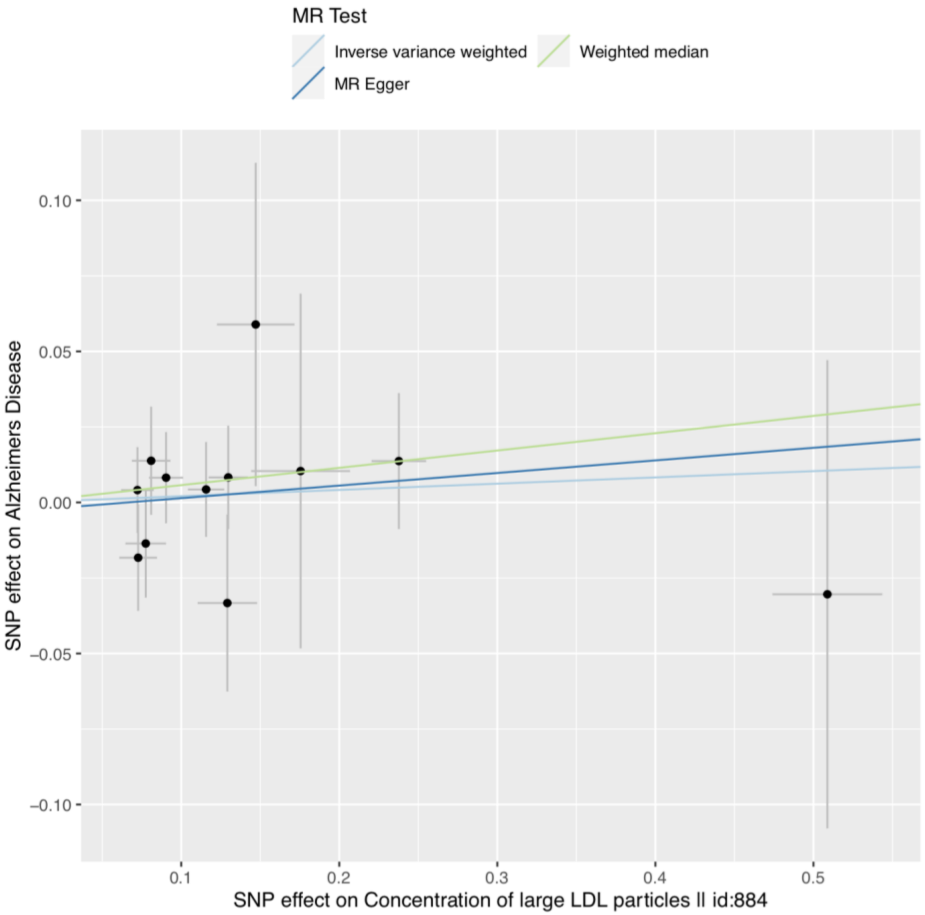
MR Test
Inverse variance weighted
Weighted median
MR Egger
SNP effect on Alzheimers Disease
SNP effect on Concentration of large LDL particles II id:884
0.10
0.05
0.00
−0.05
−0.10
0.1
0.2
0.3
0.4
0.5

**S4q**

**S4r**

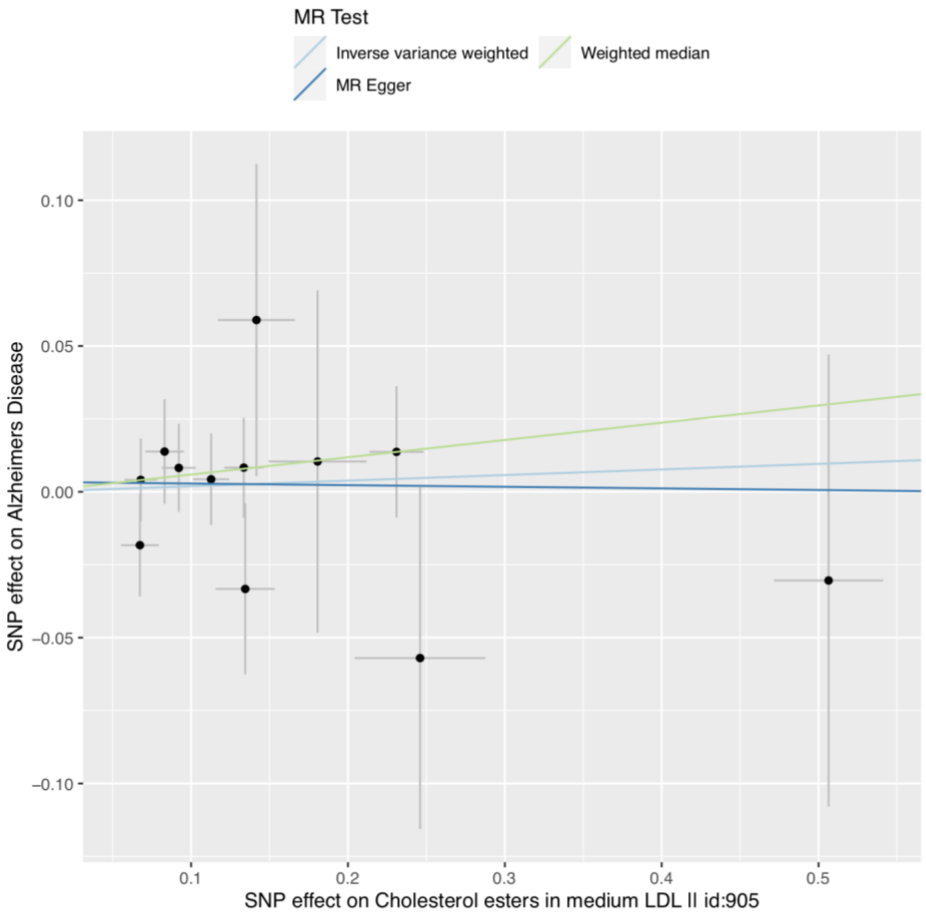
MR Test
Inverse variance weighted
Weighted median
MR Egger
0.10
0.05
0.00
−0.05
−0.10
SNP effect on Alzheimers Disease
0.1
0.2
0.3
0.4
0.5
SNP effect on Cholesterol esters in medium LDL II id:905

S4s

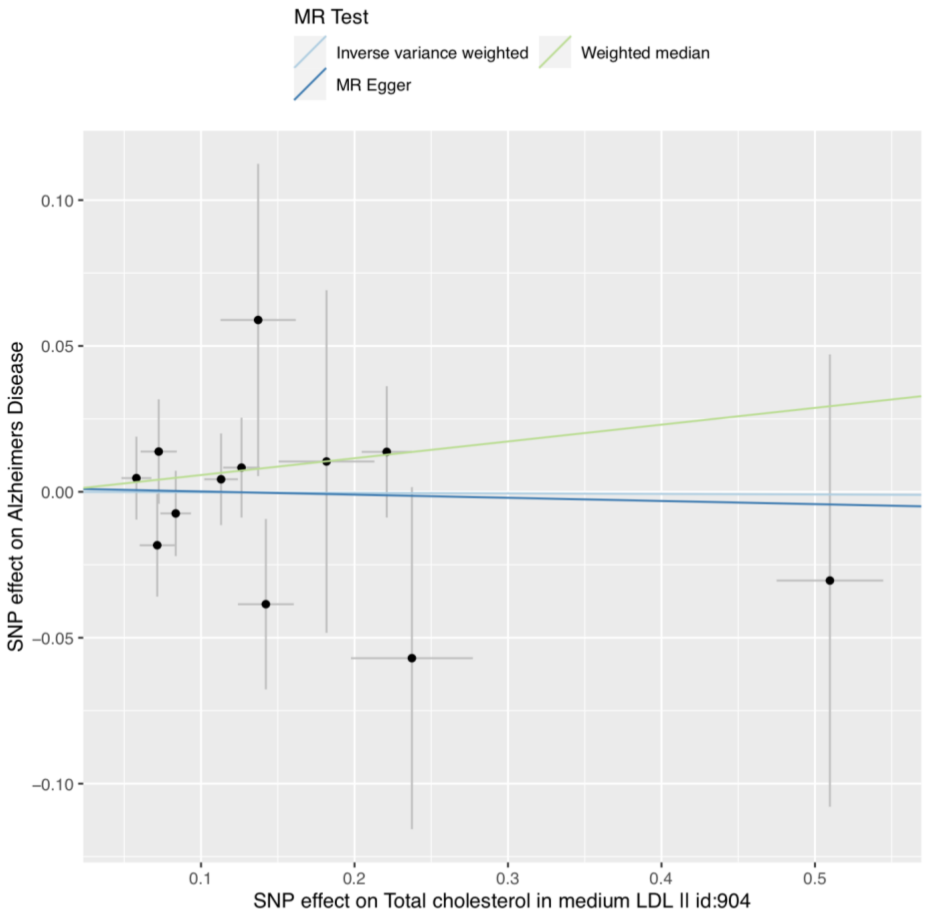
MR Test
Inverse variance weighted
Weighted median
MR Egger
0.10
0.05
0.00
−0.05
−0.10
SNP effect on Alzheimers Disease
0.1
0.2
0.3
0.4
0.5
SNP effect on Total cholesterol in medium LDL II id:904

S4t

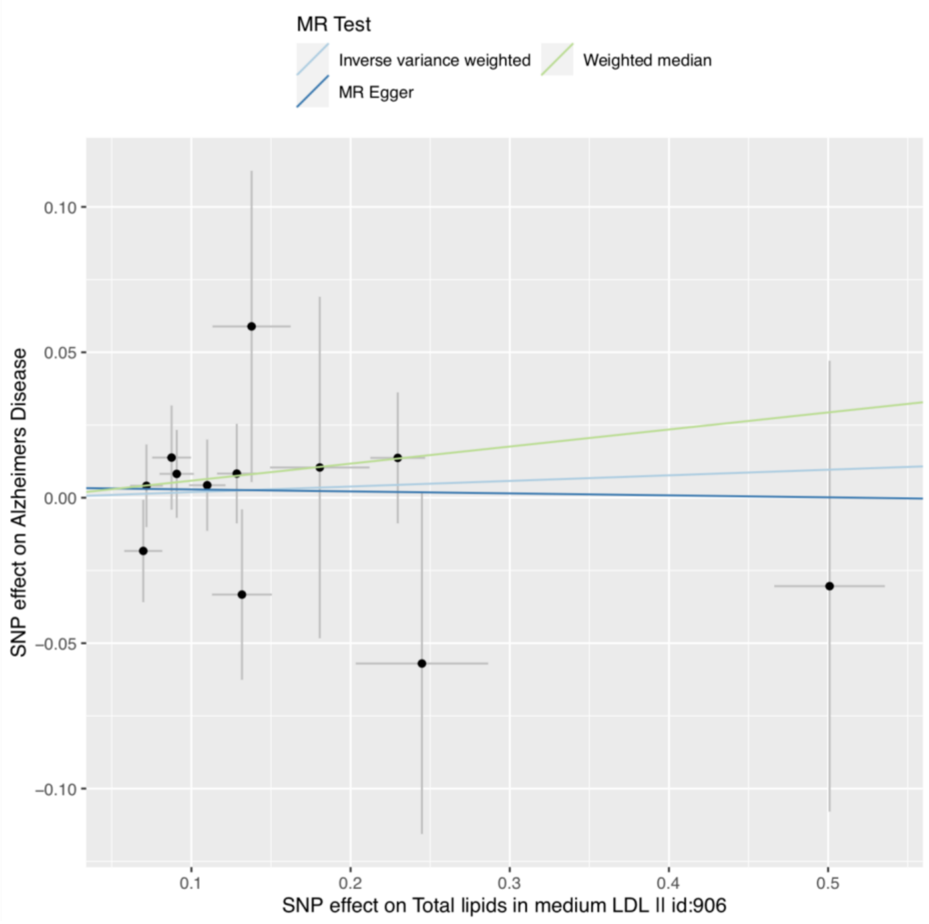
MR Test
Inverse variance weighted
Weighted median
MR Egger
0.10
0.05
0.00
−0.05
−0.10
SNP effect on Alzheimers Disease
0.1
0.2
0.3
0.4
0.5
SNP effect on Total lipids in medium LDL || id:906

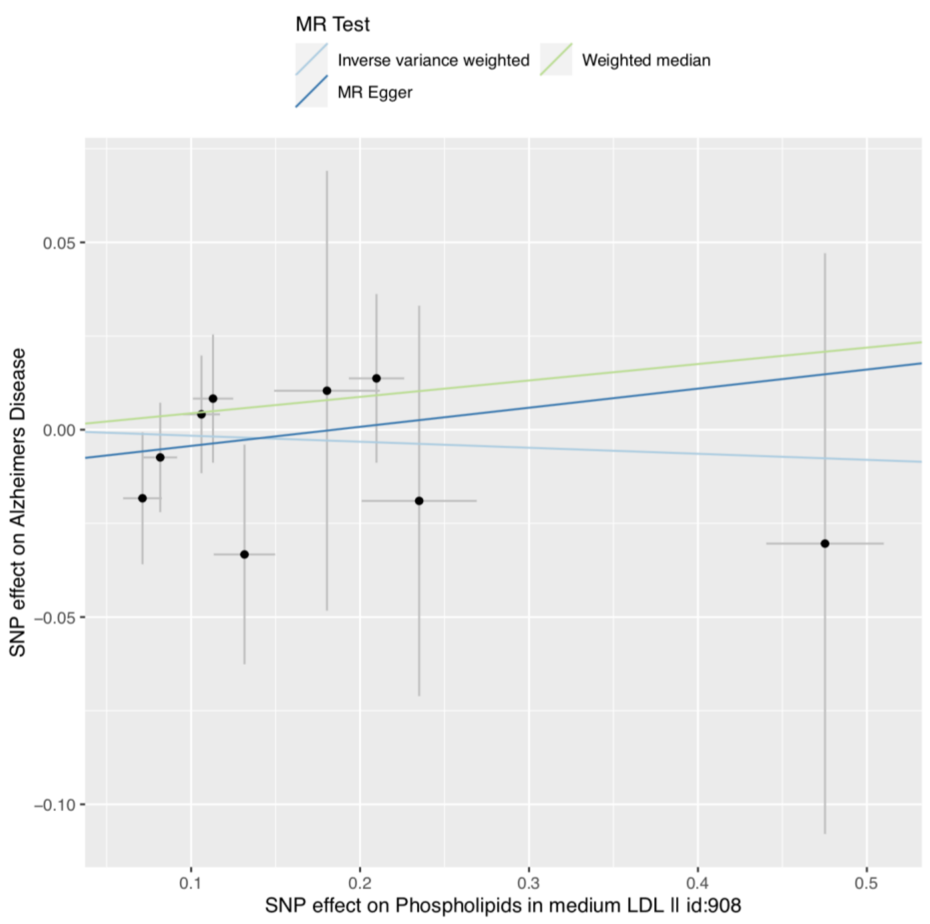
MR Test
Inverse variance weighted
Weighted median
MR Egger
0.05
0.00
−0.05
−0.10
SNP effect on Alzheimers Disease
0.1
0.2
0.3
0.4
0.5
SNP effect on Phospholipids in medium LDL || id:908

**S4u**

**S4v**

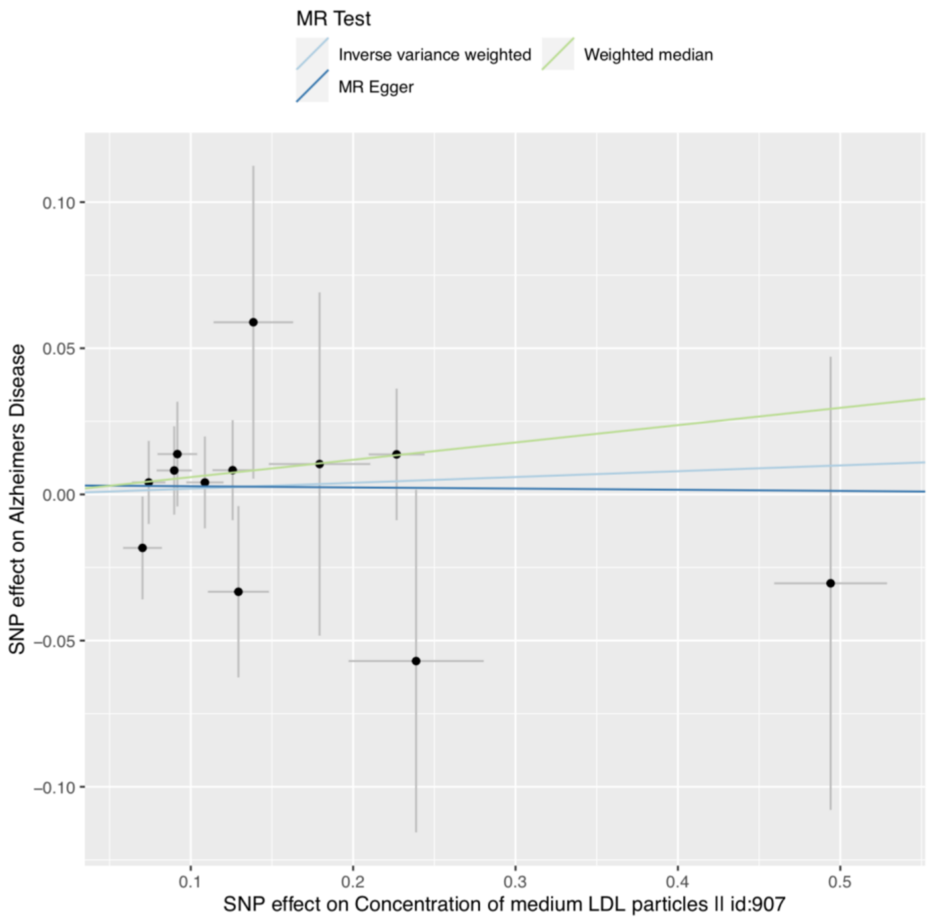
MR Test
Inverse variance weighted
Weighted median
MR Egger
SNP effect on Alzheimers Disease
SNP effect on Concentration of medium LDL particles II id:907
0.10
0.05
0.00
−0.05
−0.10
0.1
0.2
0.3
0.4
0.5

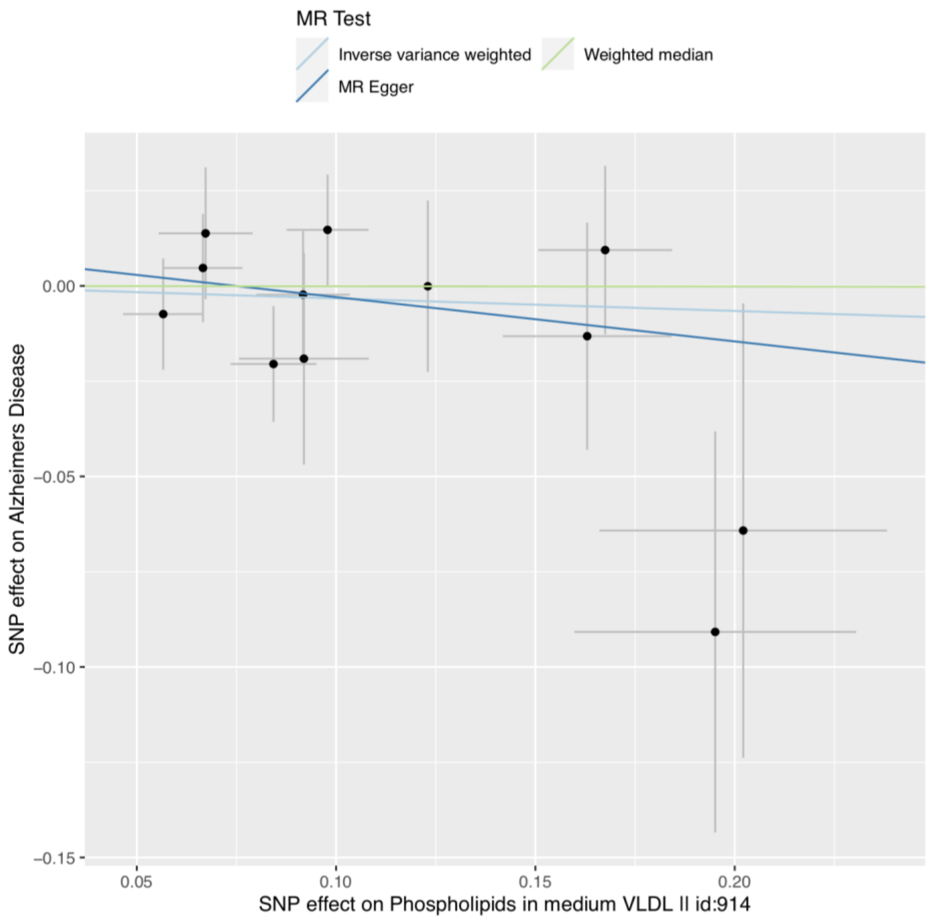
MR Test
Inverse variance weighted
Weighted median
MR Egger
SNP effect on Alzheimers Disease
SNP effect on Phospholipids in medium VLDL II id:914
0.00
−0.05
−0.10
−0.15
0.05
0.10
0.15
0.20

**S4w**

**S4x**

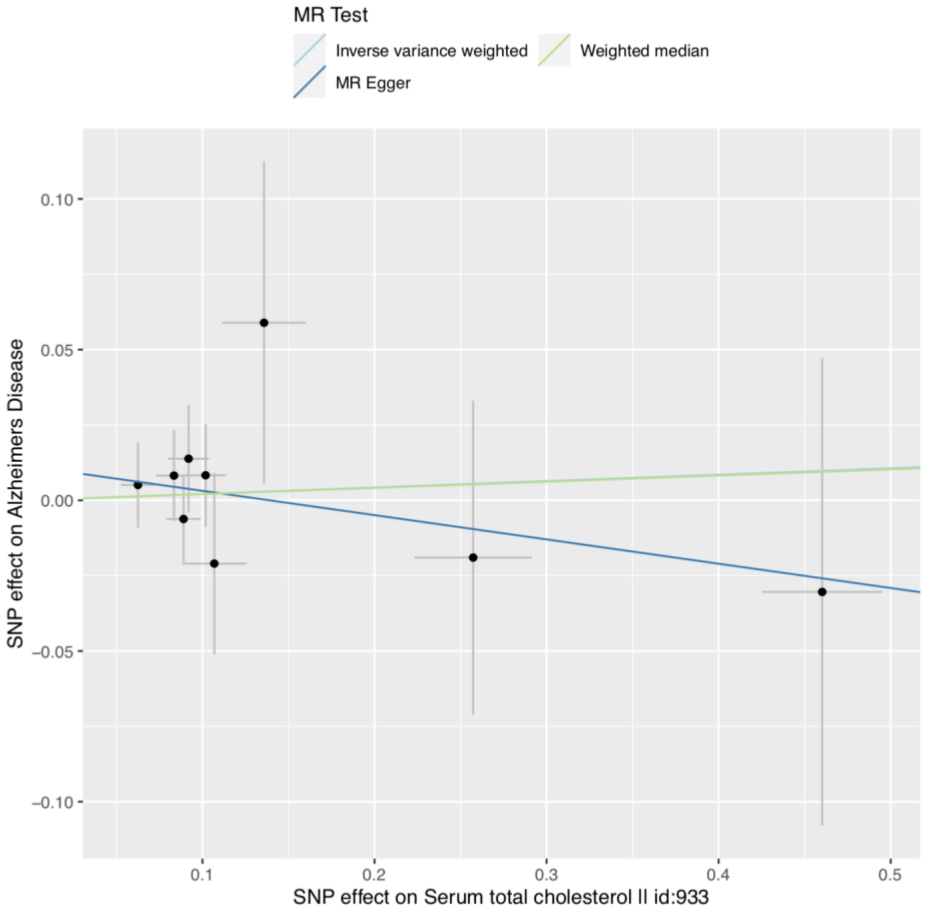
MR Test
Inverse variance weighted
Weighted median
MR Egger
0.10
0.05
0.00
−0.05
−0.10
SNP effect on Alzheimers Disease
0.1
0.2
0.3
0.4
0.5
SNP effect on Serum total cholesterol II id:933

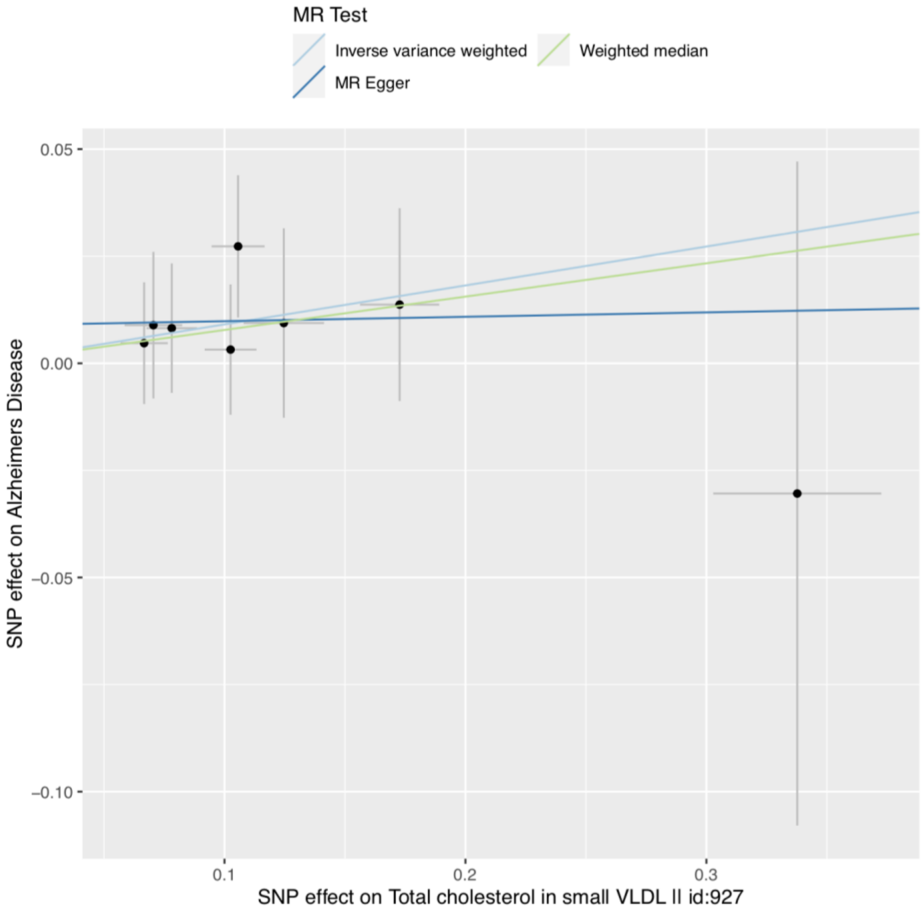
MR Test
Inverse variance weighted
Weighted median
MR Egger
0.05
0.00
−0.05
−0.10
SNP effect on Alzheimers Disease
0.1
0.2
0.3
SNP effect on Total cholesterol in small VLDL II id:927

**S4y**

**S4z**

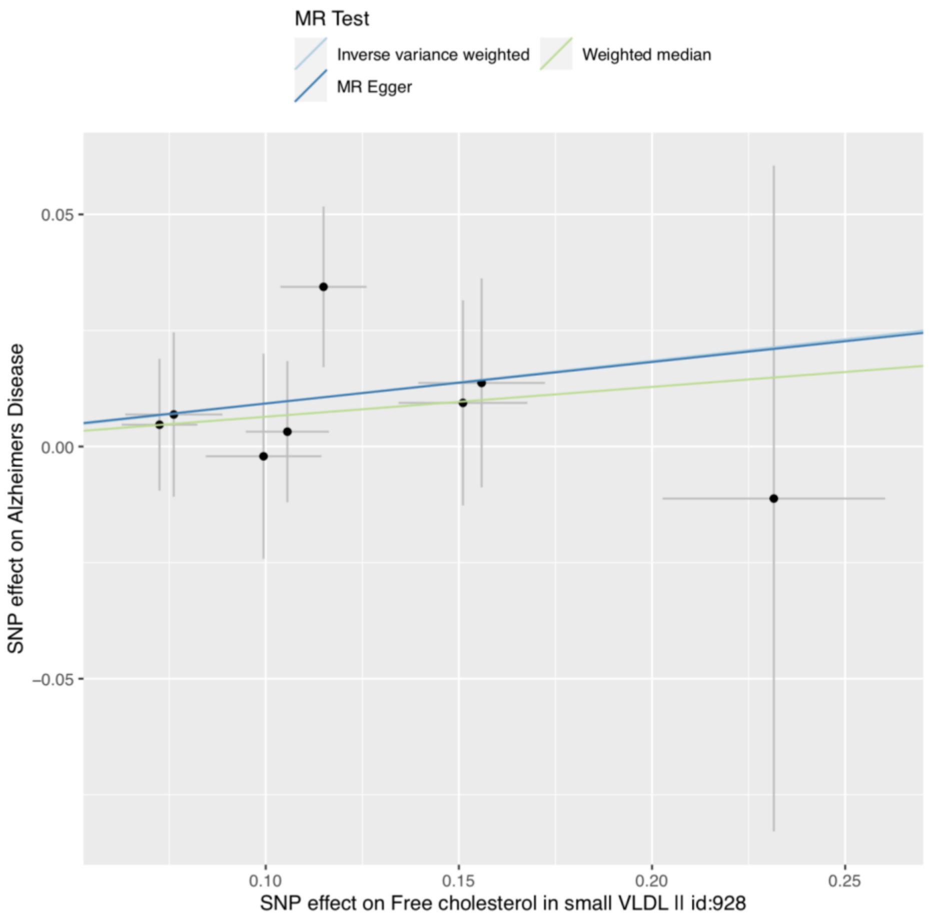
MR Test
Inverse variance weighted
Weighted median
MR Egger
0.05
0.00
−0.05
0.10
0.15
0.20
0.25
SNP effect on Alzheimers Disease
SNP effect on Free cholesterol in small VLDL II id:928

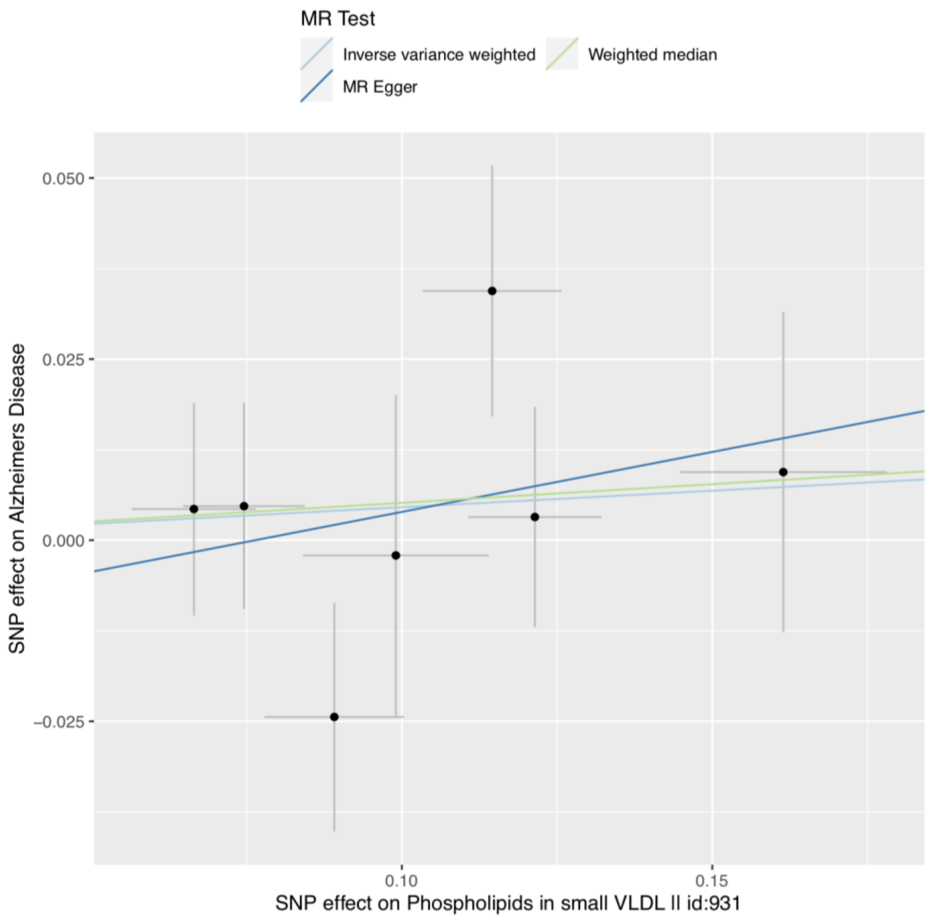
MR Test
Inverse variance weighted
Weighted median
MR Egger
0.050
0.025
0.000
−0.025
0.10
0.15
SNP effect on Alzheimers Disease
SNP effect on Phospholipids in small VLDL II id:931

**S4aa**

**S4ab**

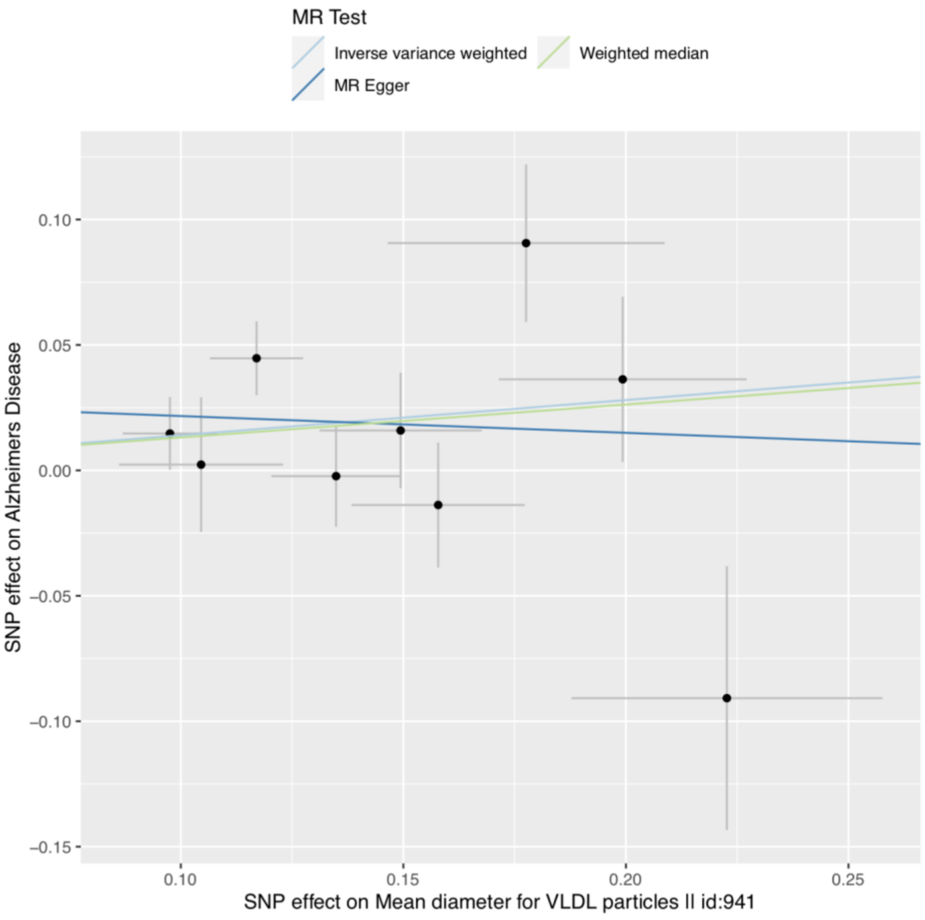
MR Test
Inverse variance weighted
Weighted median
MR Egger
0.10
0.05
0.00
−0.05
−0.10
−0.15
SNP effect on Alzheimers Disease
0.10
0.15
0.20
0.25
SNP effect on Mean diameter for VLDL particles II id:941

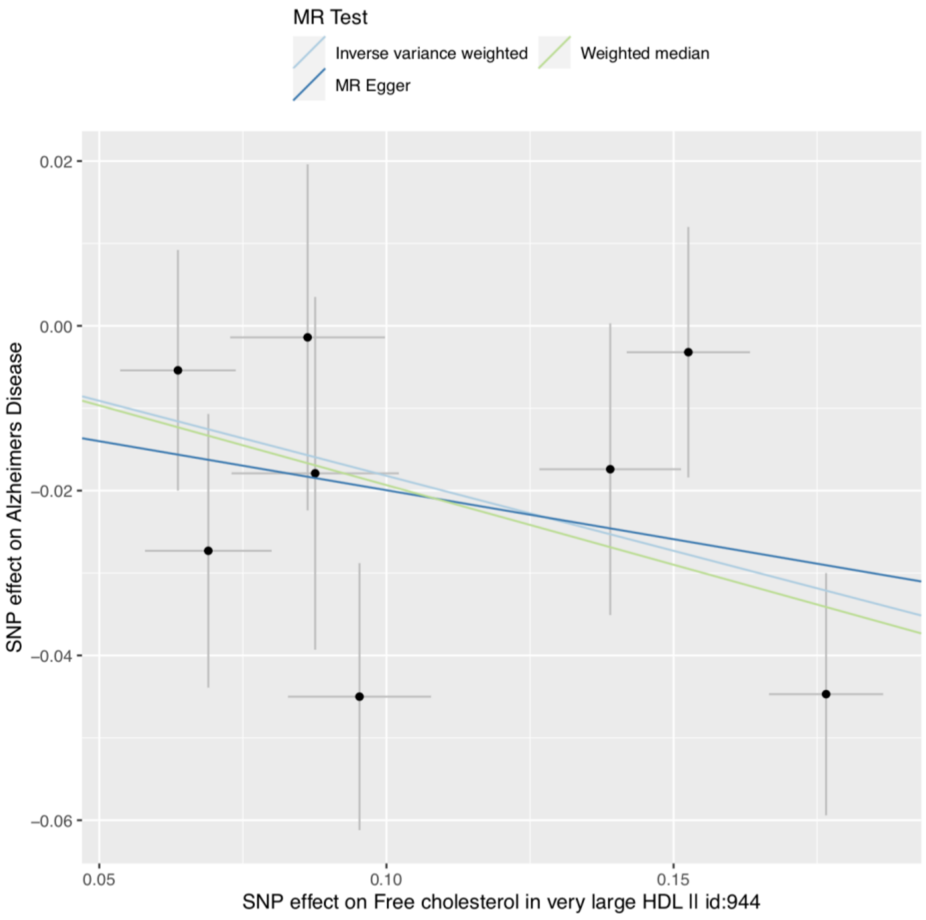
MR Test
Inverse variance weighted
Weighted median
MR Egger
0.02
0.00
−0.02
−0.04
−0.06
SNP effect on Alzheimers Disease
0.05
0.10
0.15
SNP effect on Free cholesterol in very large HDL II id:944

**S4ac**

**S4ad**

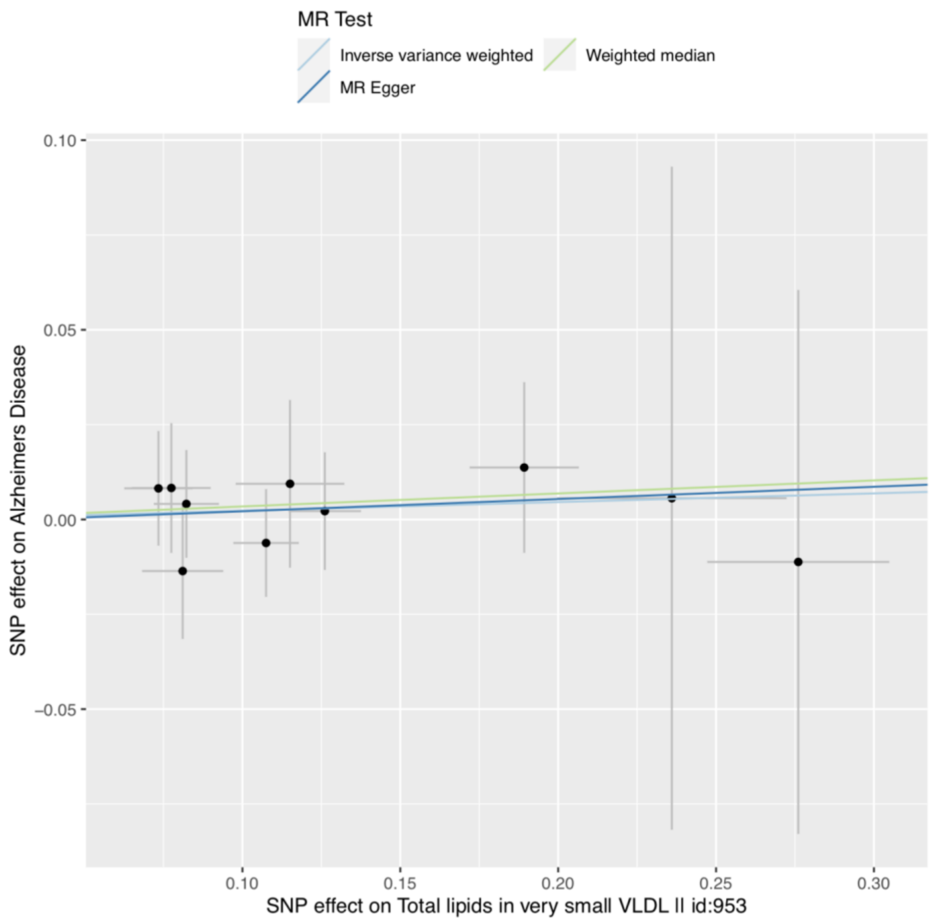
MR Test
Inverse variance weighted
Weighted median
MR Egger
SNP effect on Alzheimers Disease
SNP effect on Total lipids in very small VLDL II id:953
0.10
0.05
0.00
−0.05
0.10
0.15
0.20
0.25
0.30

**S4ae**

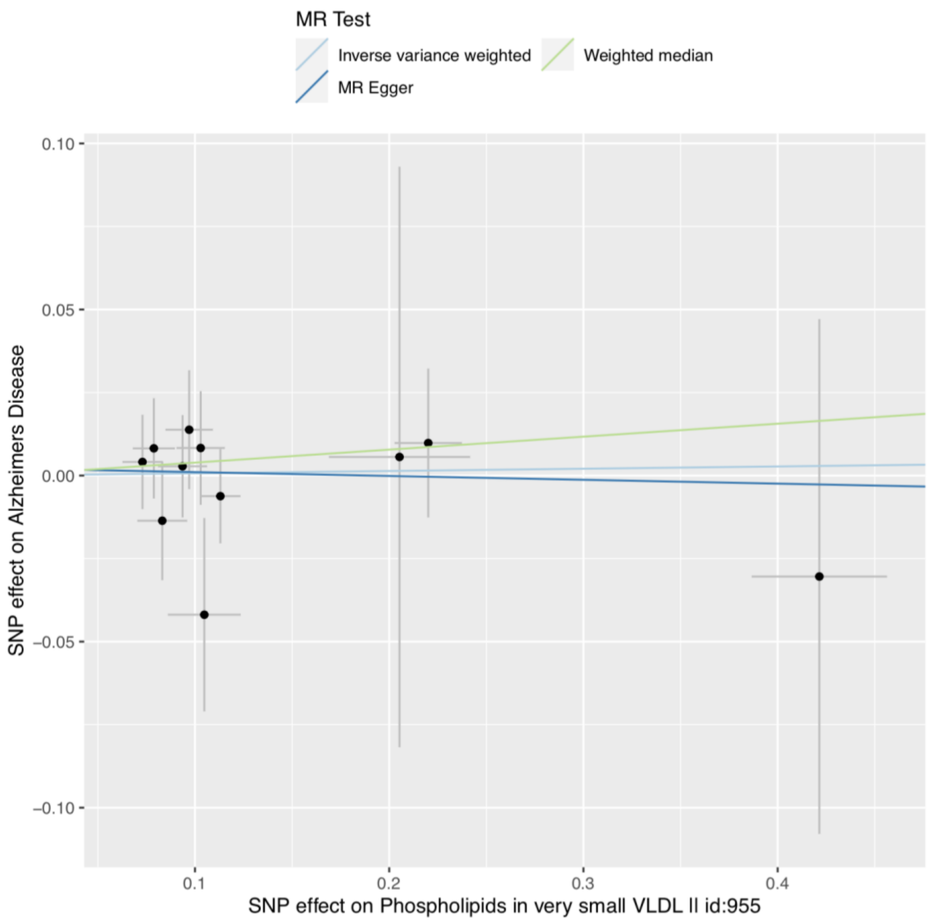
MR Test
Inverse variance weighted
Weighted median
MR Egger
SNP effect on Alzheimers Disease
SNP effect on Phospholipids in very small VLDL II id:955
0.10
0.05
0.00
−0.05
−0.10
0.1
0.2
0.3
0.4

**S4af**

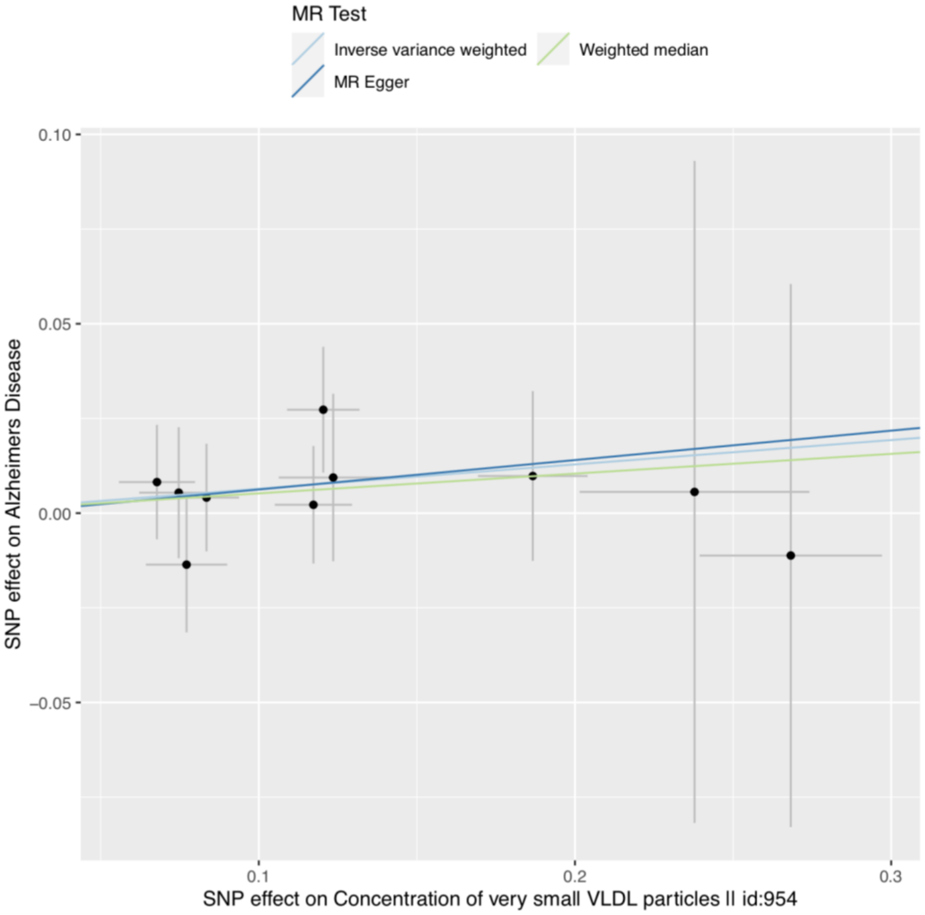
MR Test
Inverse variance weighted
Weighted median
MR Egger
0.10
0.05
0.00
−0.05
0.1
0.2
0.3
SNP effect on Alzheimers Disease
SNP effect on Concentration of very small VLDL particles II id:954

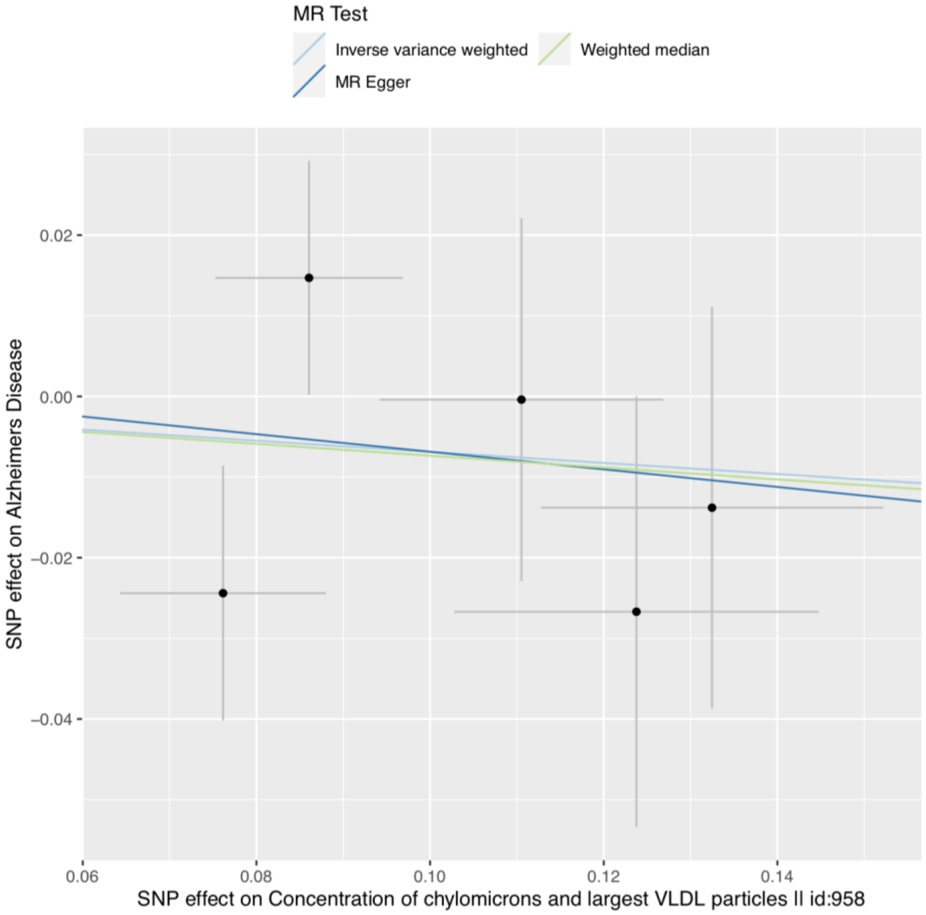
MR Test
Inverse variance weighted
Weighted median
MR Egger
0.02
0.00
−0.02
−0.04
0.06
0.08
0.10
0.12
0.14
SNP effect on Alzheimers Disease
SNP effect on Concentration of chylomicrons and largest VLDL particles II id:958

**S4ag**

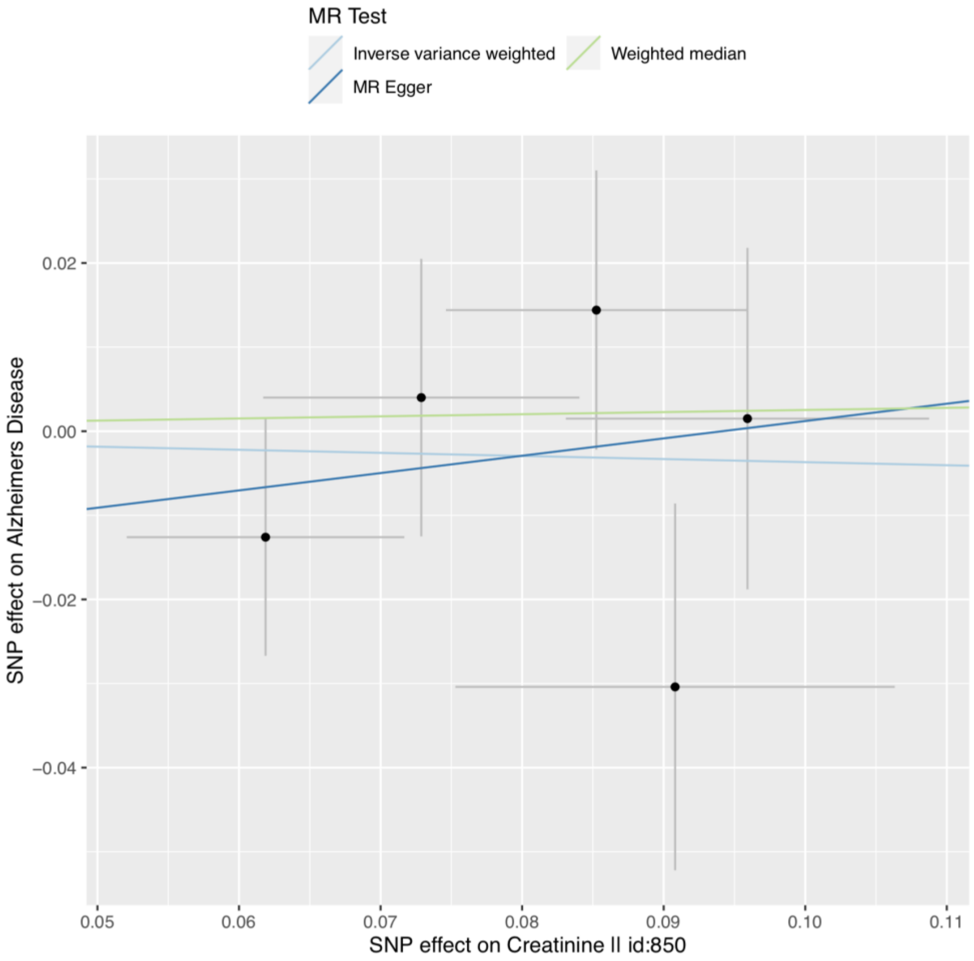


**Supplementary Figure S4(a-ag).** Scatter plot illustrating primary (IVW) and secondary (Egger and weighted median) causal slope estimates when each metabolite is set as exposure and AD set as outcome.

**S5a**

**S5b**

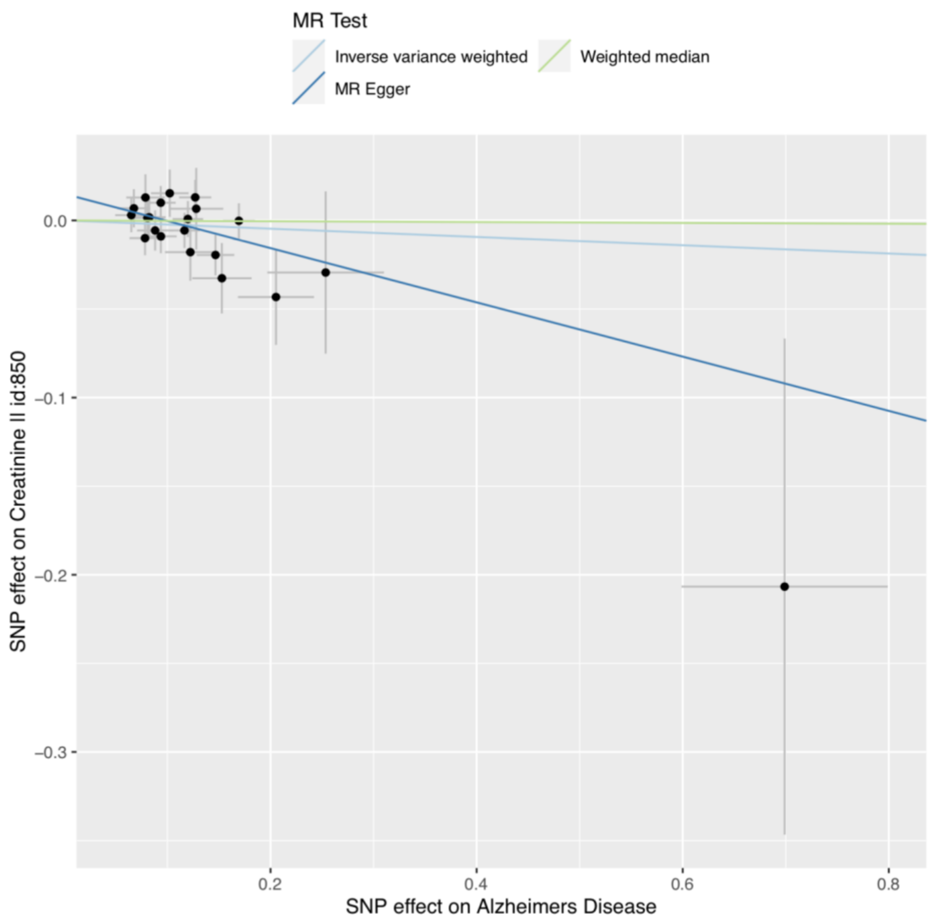
MR Test
Inverse variance weighted
Weighted median
MR Egger
SNP effect on Creatinine II id:850
SNP effect on Alzheimers Disease
0.0
−0.1
−0.2
−0.3
0.2
0.4
0.6
0.8

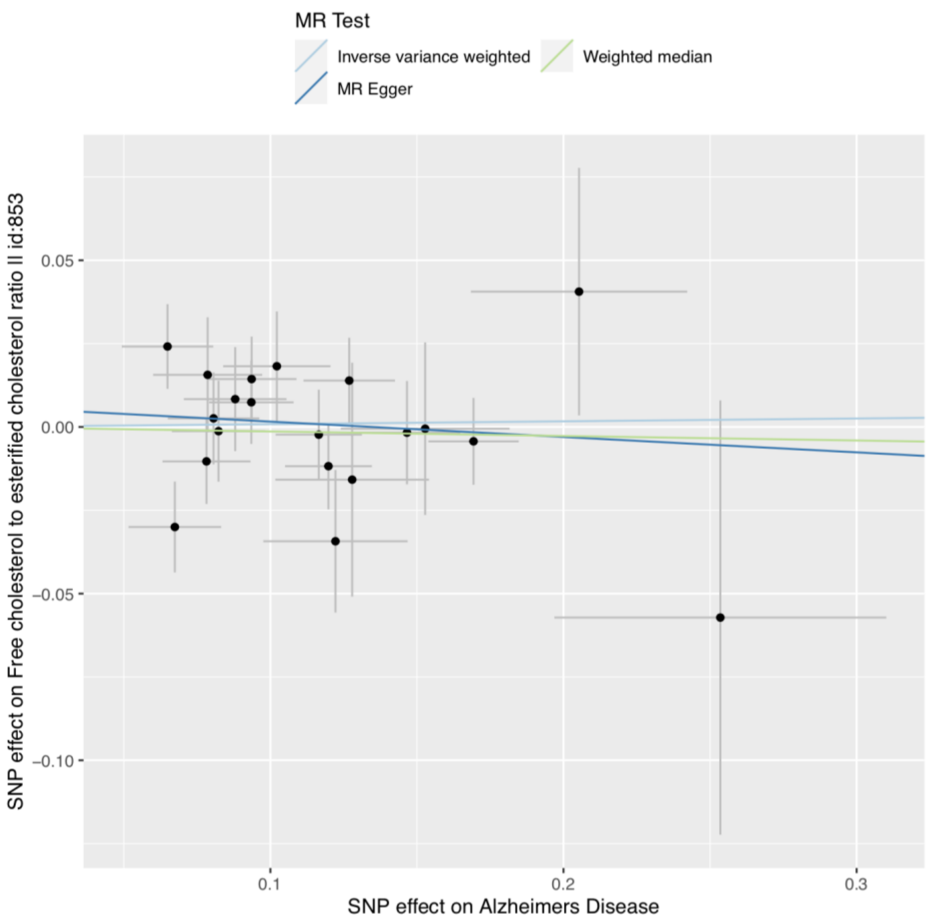
MR Test
Inverse variance weighted
Weighted median
MR Egger
SNP effect on Free cholesterol to esterified cholesterol ratio II id:853
SNP effect on Alzheimers Disease
0.05
0.00
−0.05
−0.10
0.1
0.2
0.3

**S5c**

**S5d**

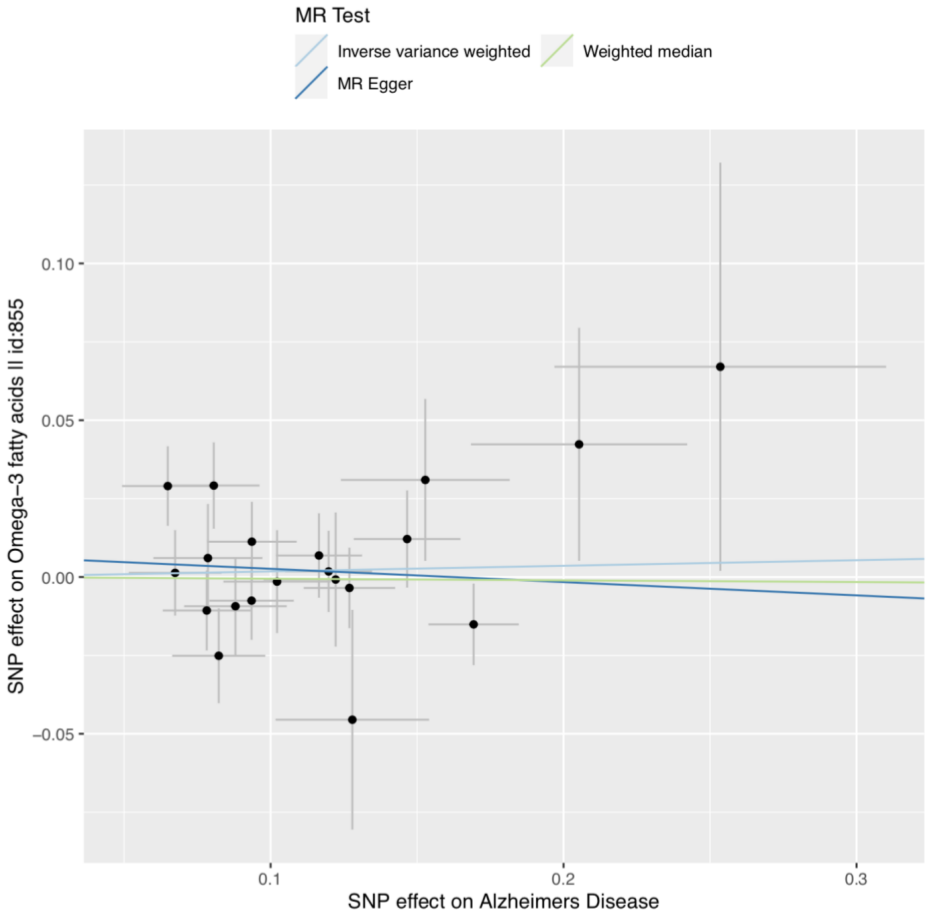
MR Test
Inverse variance weighted
Weighted median
MR Egger
0.10
0.05
0.00
−0.05
SNP effect on Omega−3 fatty acids II id:855
0.1
0.2
0.3
SNP effect on Alzheimers Disease

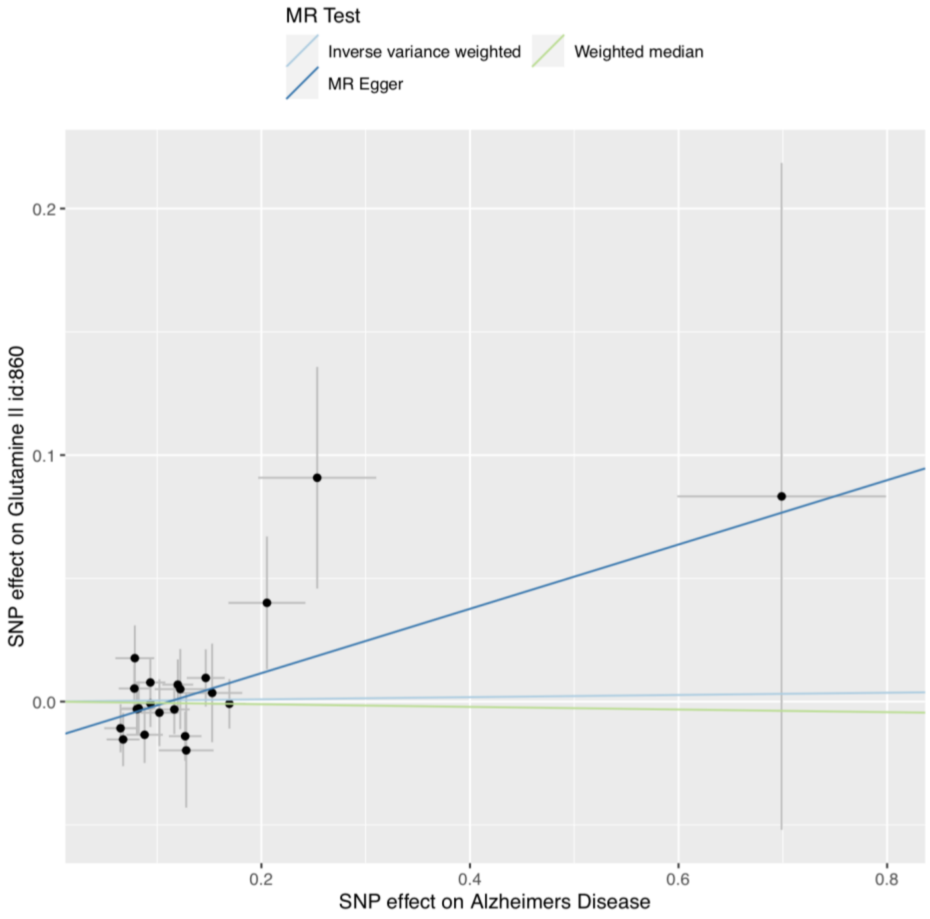
MR Test
Inverse variance weighted
Weighted median
MR Egger
0.2
0.1
0.0
SNP effect on Glutamine II id:860
0.2
0.4
0.6
0.8
SNP effect on Alzheimers Disease

**S5e**

**S5f**

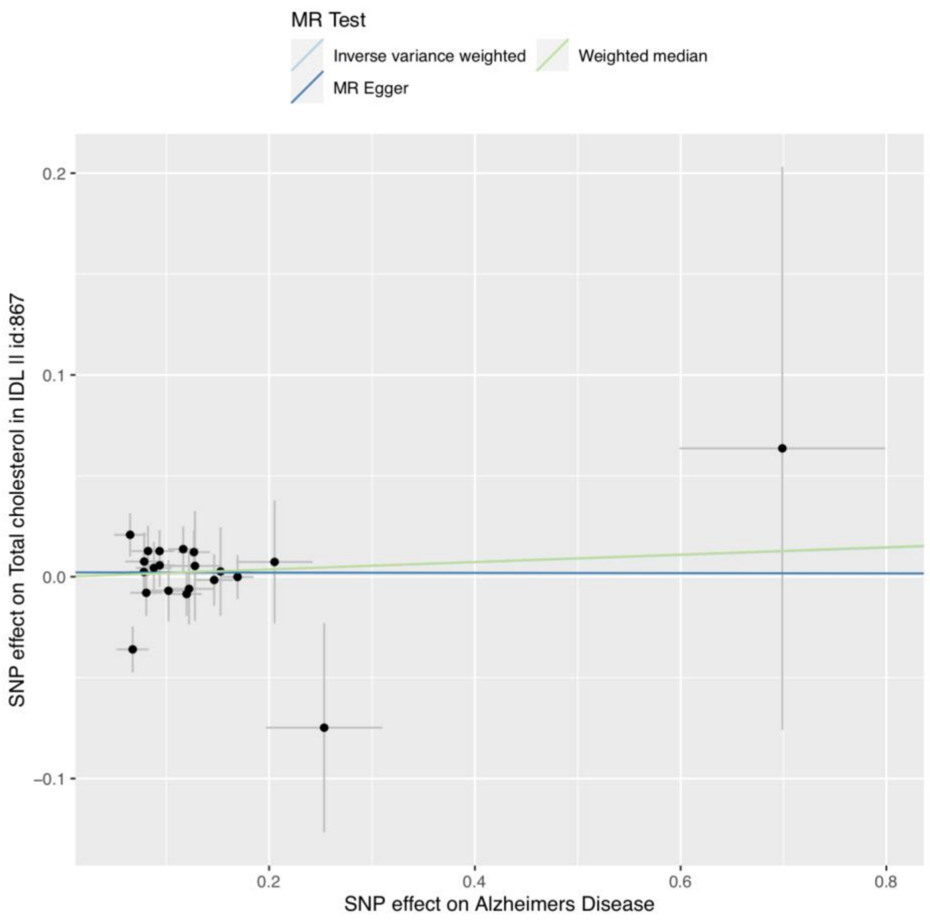
MR Test
Inverse variance weighted
Weighted median
MR Egger
SNP effect on Total cholesterol in IDL II id:867
SNP effect on Alzheimers Disease
0.2
0.1
0.0
−0.1
0.2
0.4
0.6
0.8

**S5g**

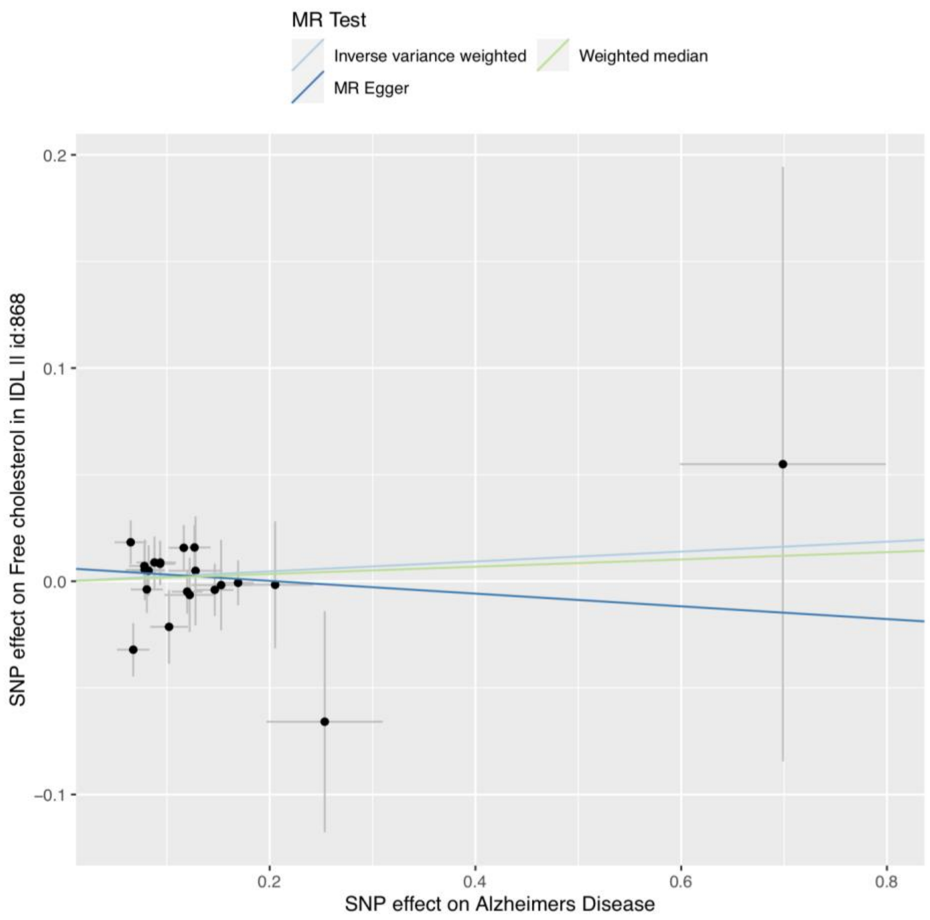
MR Test
Inverse variance weighted
Weighted median
MR Egger
SNP effect on Free cholesterol in IDL II id:868
SNP effect on Alzheimers Disease
0.2
0.1
0.0
−0.1
0.2
0.4
0.6
0.8

**S5h**

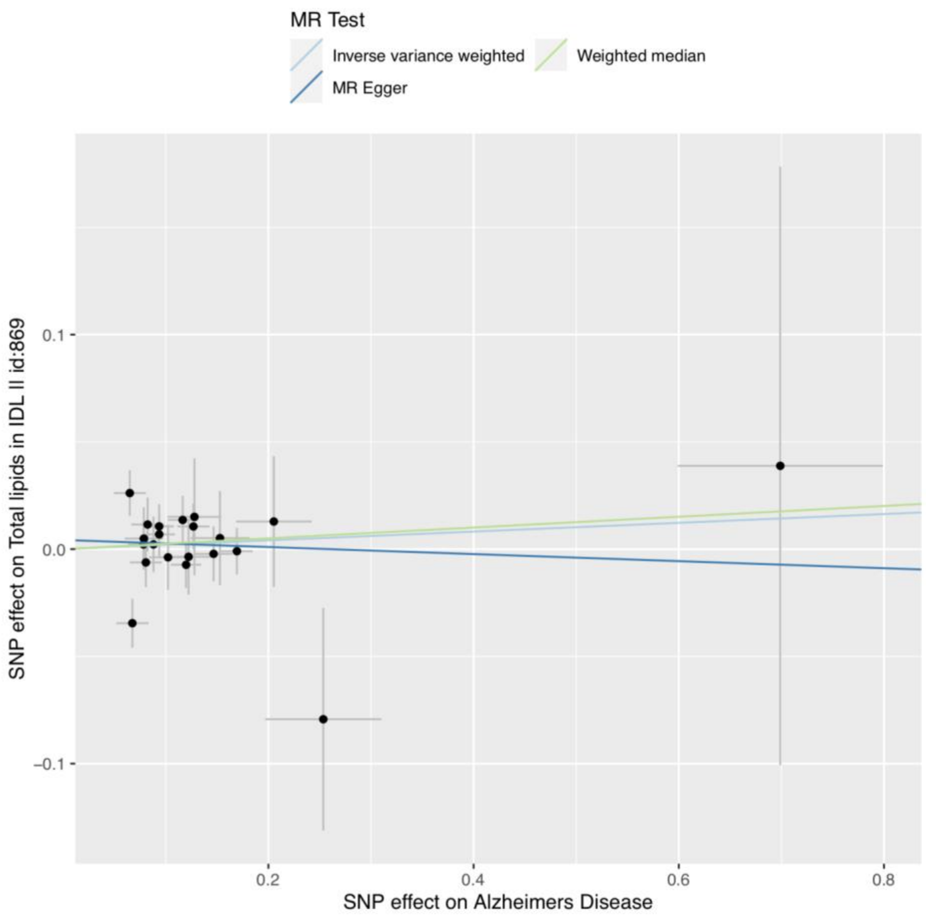
MR Test
Inverse variance weighted
Weighted median
MR Egger
SNP effect on Total lipids in IDL II id:869
0.1
0.0
−0.1
0.2
0.4
0.6
0.8
SNP effect on Alzheimers Disease

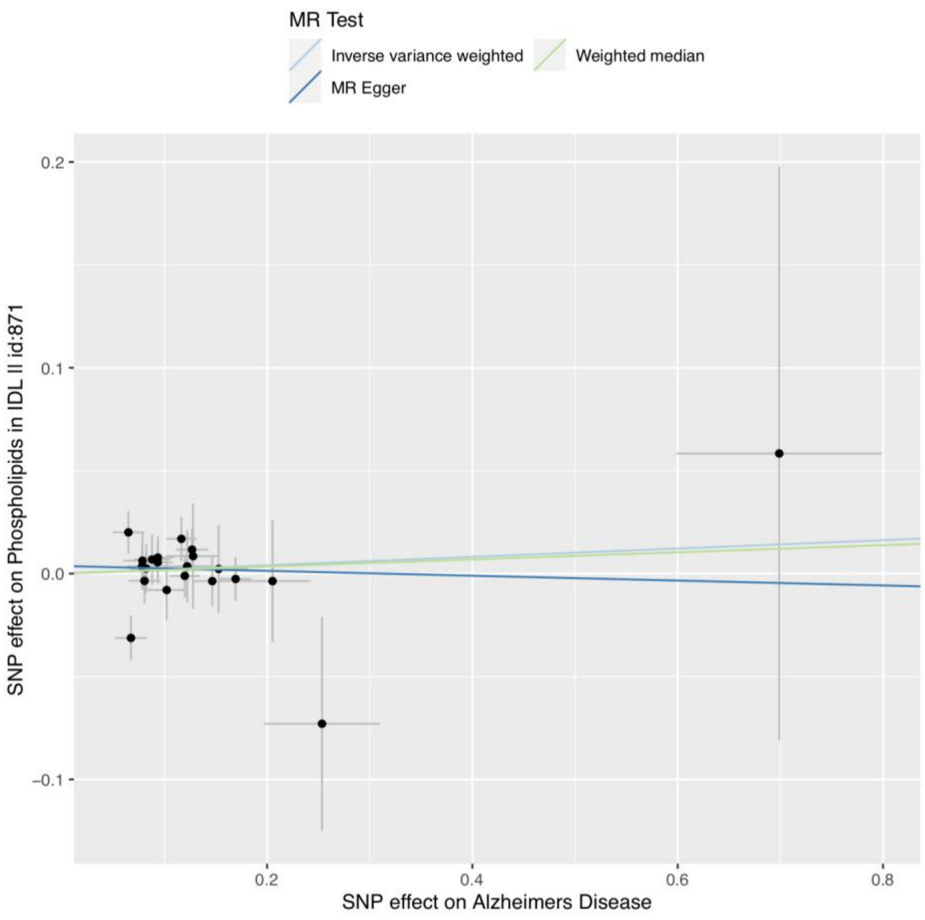
MR Test
Inverse variance weighted
Weighted median
MR Egger
SNP effect on Phospholipids in IDL II id:871
0.2
0.1
0.0
−0.1
0.2
0.4
0.6
0.8
SNP effect on Alzheimers Disease

**S5i**

**S5j**

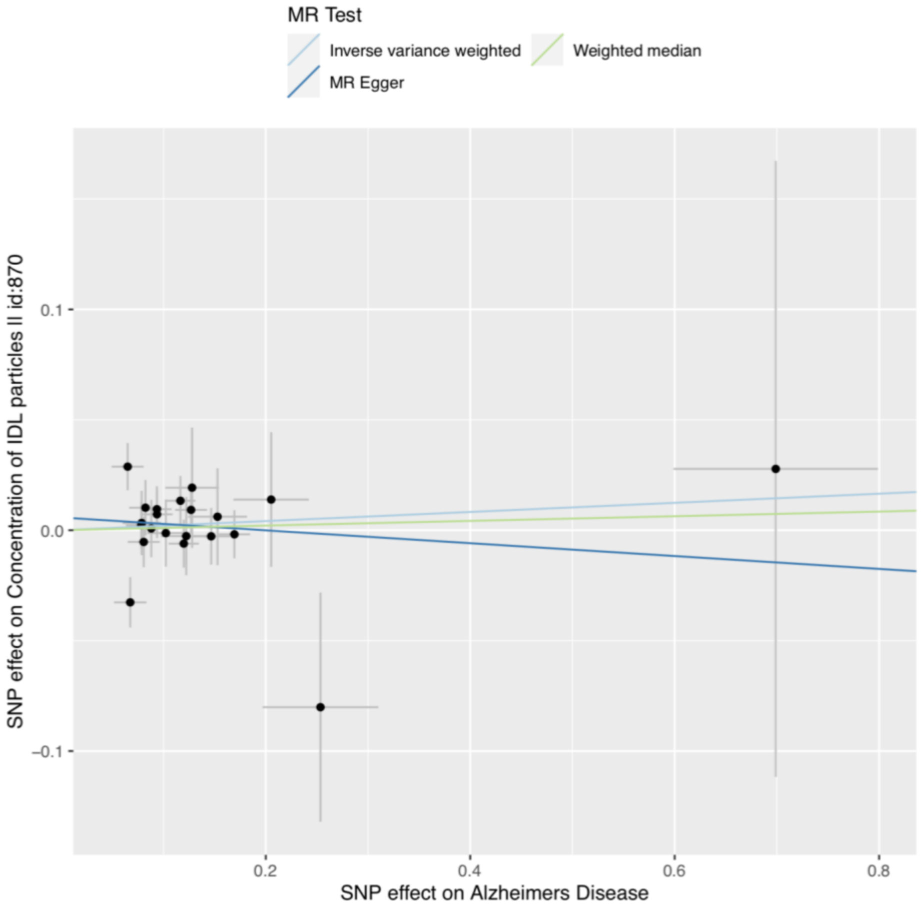
MR Test
Inverse variance weighted
Weighted median
MR Egger
SNP effect on Concentration of IDL particles II id:870
0.1
0.0
−0.1
0.2
0.4
0.6
0.8
SNP effect on Alzheimers Disease

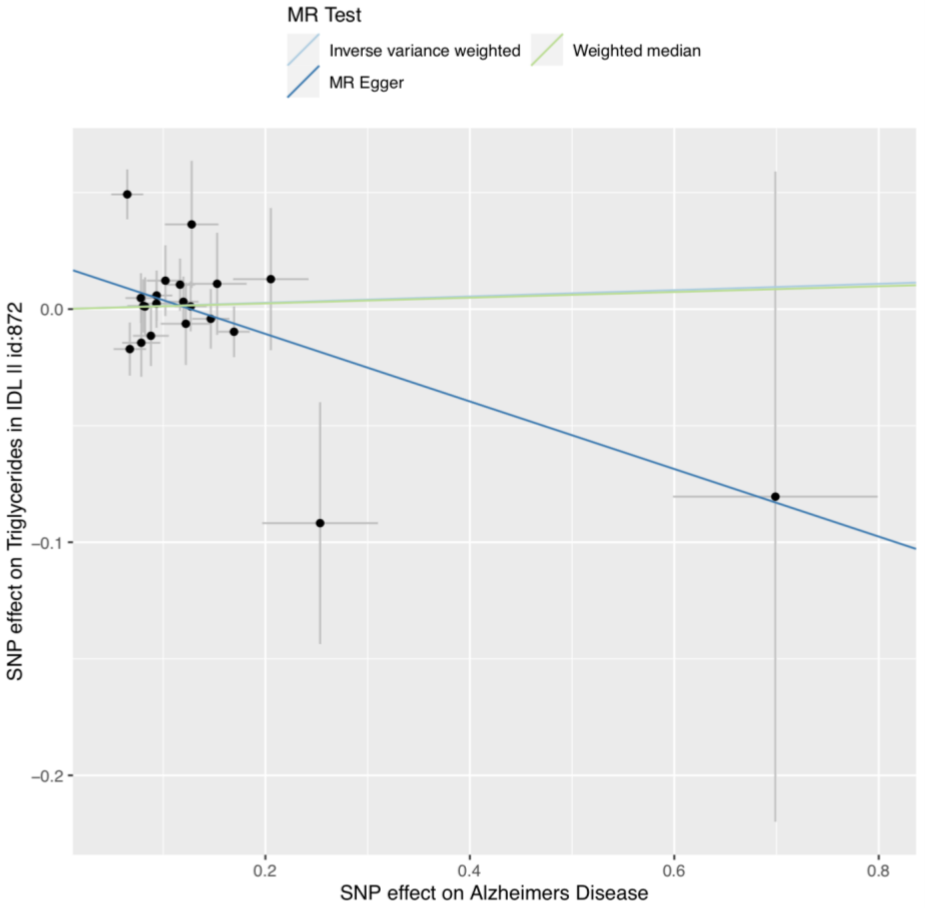
MR Test
Inverse variance weighted
Weighted median
MR Egger
SNP effect on Triglycerides in IDL II id:872
0.0
−0.1
−0.2
0.2
0.4
0.6
0.8
SNP effect on Alzheimers Disease

**S5k**

**S5l**

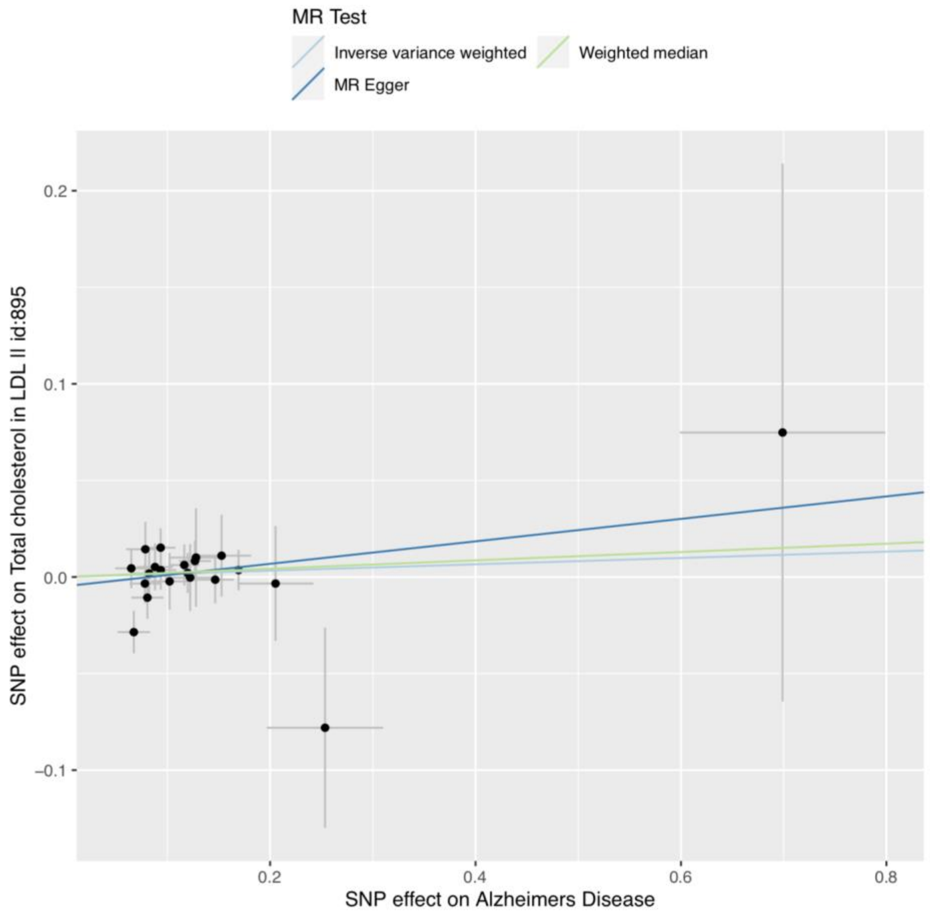
MR Test
Inverse variance weighted
Weighted median
MR Egger
SNP effect on Total cholesterol in LDL II id:895
SNP effect on Alzheimers Disease
0.2
0.1
0.0
−0.1
0.2
0.4
0.6
0.8

**S5m**

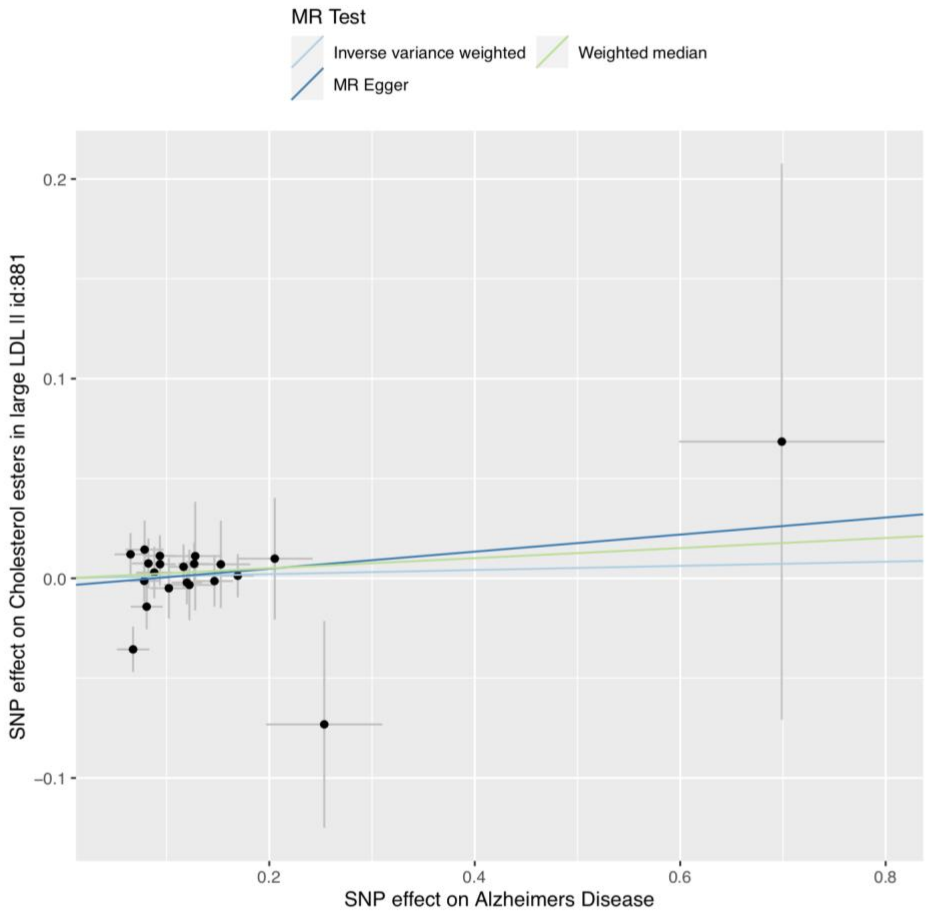
MR Test
Inverse variance weighted
Weighted median
MR Egger
SNP effect on Cholesterol esters in large LDL II id:881
SNP effect on Alzheimers Disease
0.2
0.1
0.0
−0.1
0.2
0.4
0.6
0.8

**S5n**

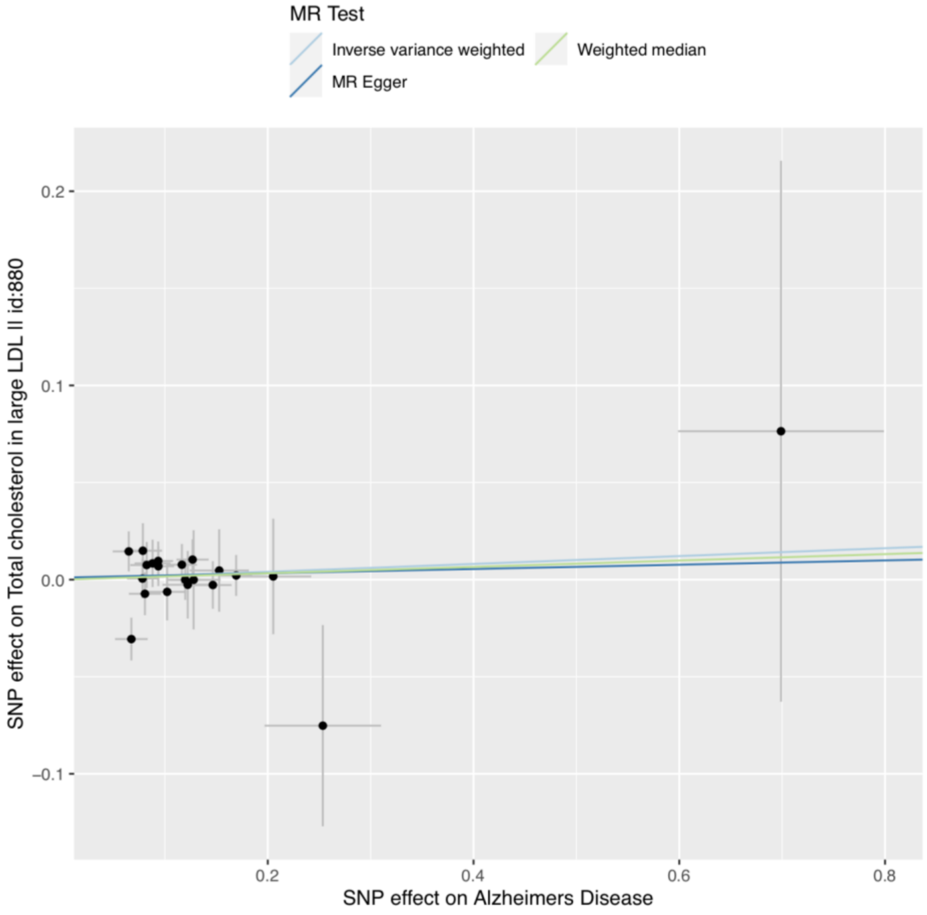
MR Test
Inverse variance weighted
Weighted median
MR Egger
SNP effect on Total cholesterol in large LDL II id:880
SNP effect on Alzheimers Disease
0.2
0.1
0.0
−0.1
0.2
0.4
0.6
0.8

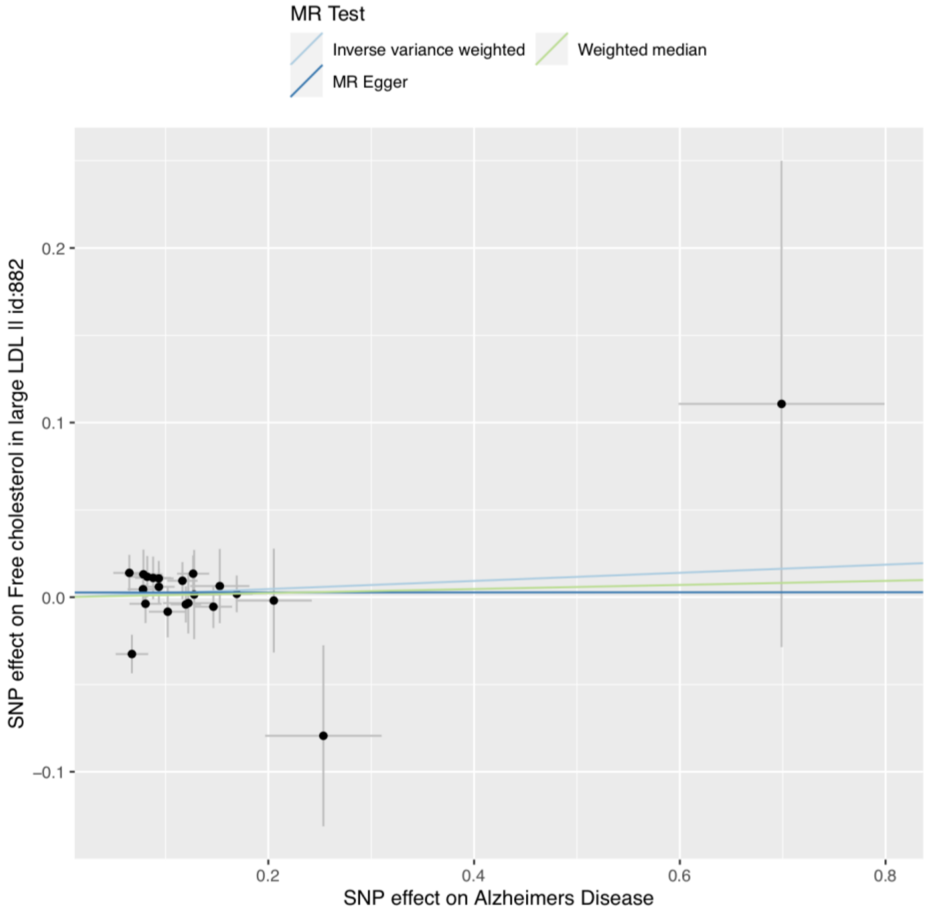
MR Test
Inverse variance weighted
Weighted median
MR Egger
SNP effect on Free cholesterol in large LDL II id:882
SNP effect on Alzheimers Disease
0.2
0.1
0.0
−0.1
0.2
0.4
0.6
0.8

**S5o**

**S5p**

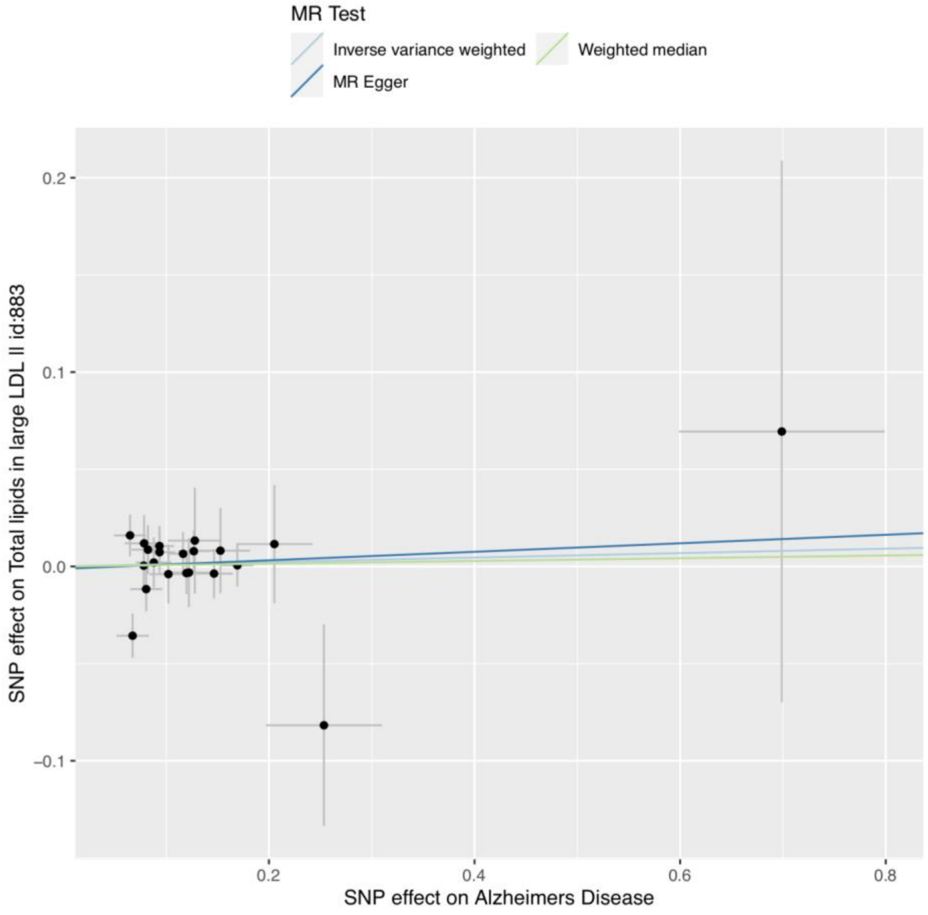
MR Test
Inverse variance weighted
Weighted median
MR Egger
SNP effect on Total lipids in large LDL II id:883
SNP effect on Alzheimers Disease
0.2
0.1
0.0
−0.1
0.2
0.4
0.6
0.8

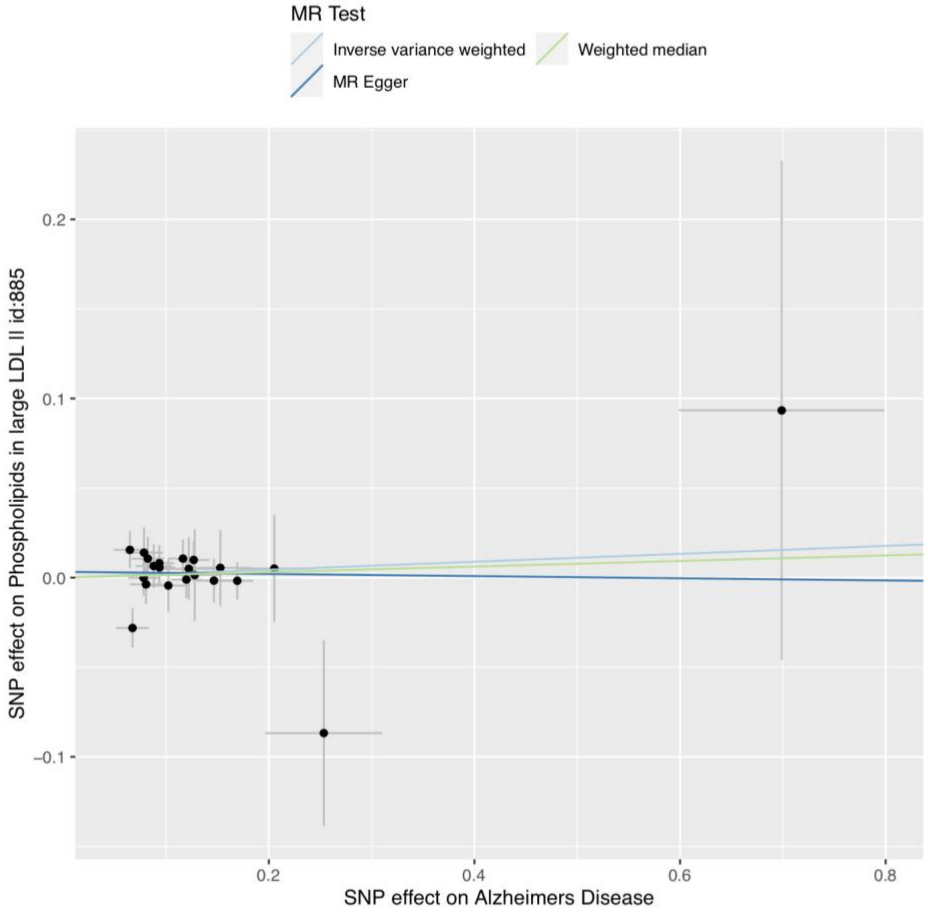
MR Test
Inverse variance weighted
Weighted median
MR Egger
SNP effect on Phospholipids in large LDL II id:885
SNP effect on Alzheimers Disease
0.2
0.1
0.0
−0.1
0.2
0.4
0.6
0.8

**S5q**

**S5r**

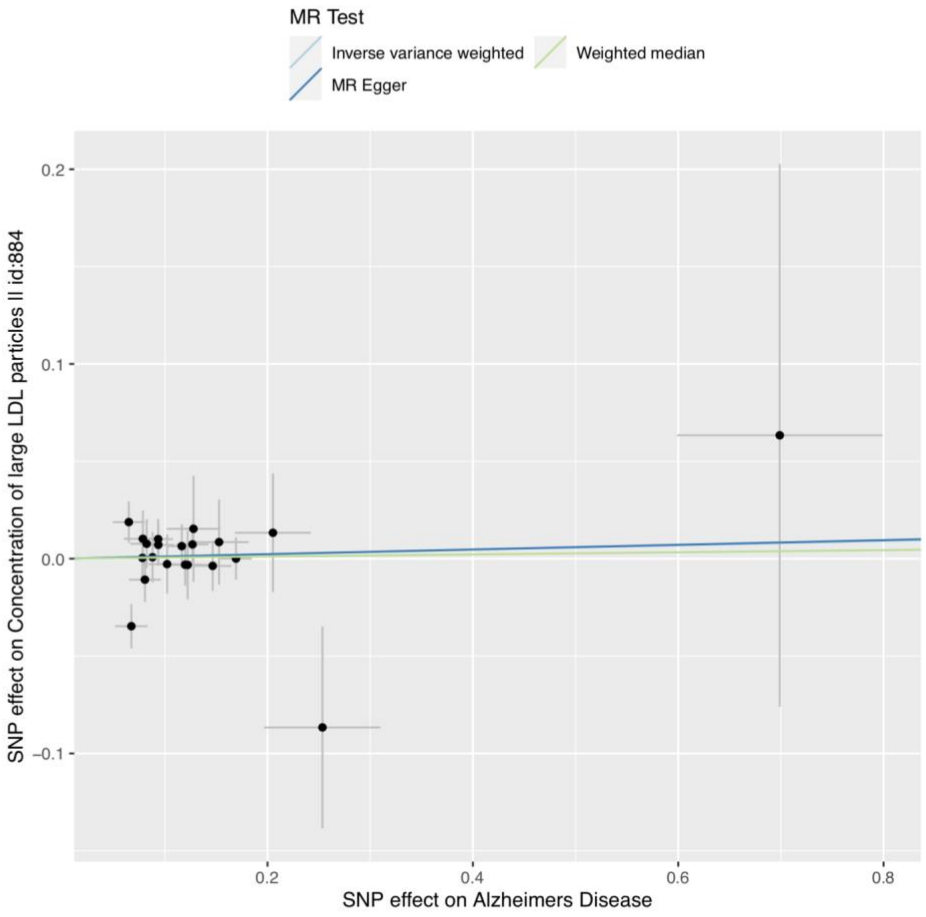
MR Test
Inverse variance weighted
Weighted median
MR Egger
0.2
0.1
0.0
−0.1
SNP effect on Concentration of large LDL particles II id:884
0.2
0.4
0.6
0.8
SNP effect on Alzheimers Disease

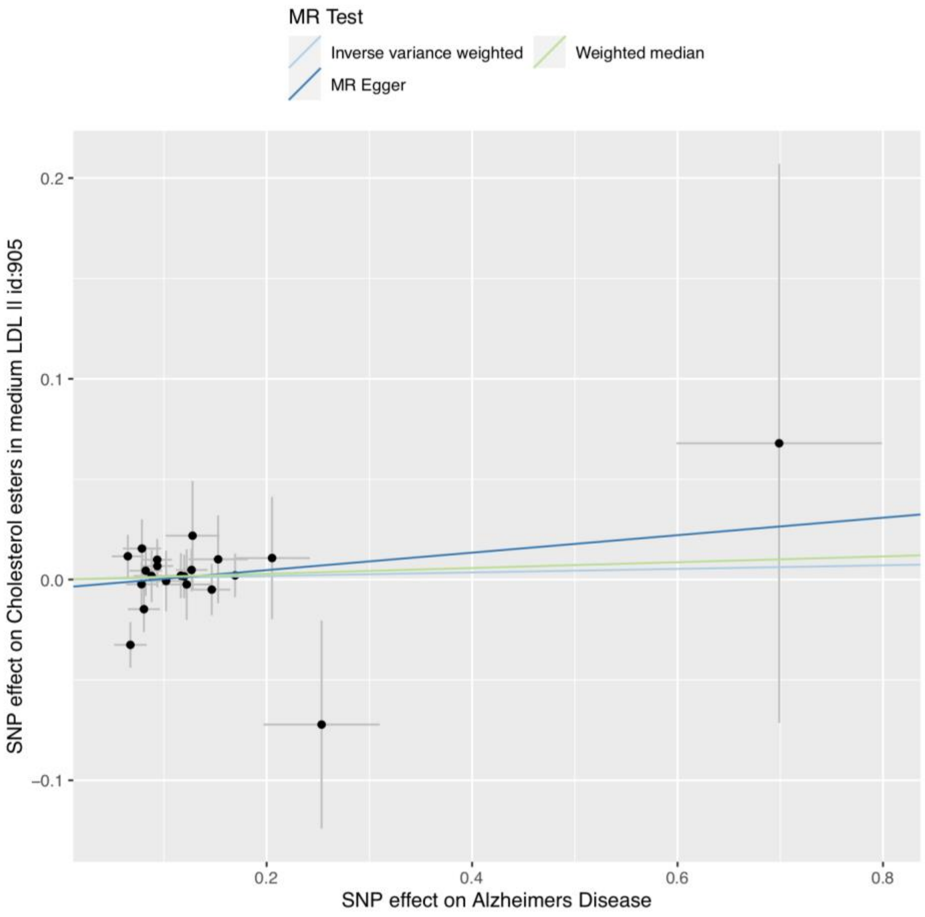
MR Test
Inverse variance weighted
Weighted median
MR Egger
0.2
0.1
0.0
−0.1
SNP effect on Cholesterol esters in medium LDL II id:905
0.2
0.4
0.6
0.8
SNP effect on Alzheimers Disease

**S5s**

**S5t**

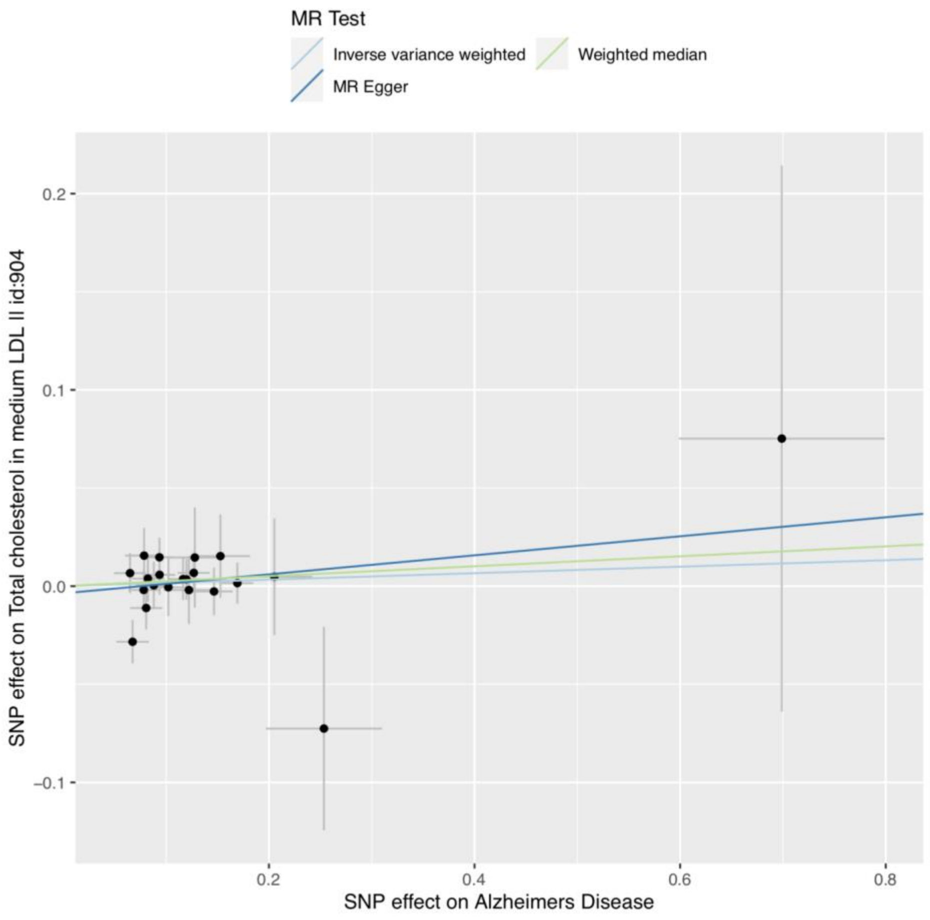
MR Test
Inverse variance weighted
Weighted median
MR Egger
SNP effect on Total cholesterol in medium LDL II id:904
SNP effect on Alzheimers Disease
0.2
0.1
0.0
−0.1
0.2
0.4
0.6
0.8

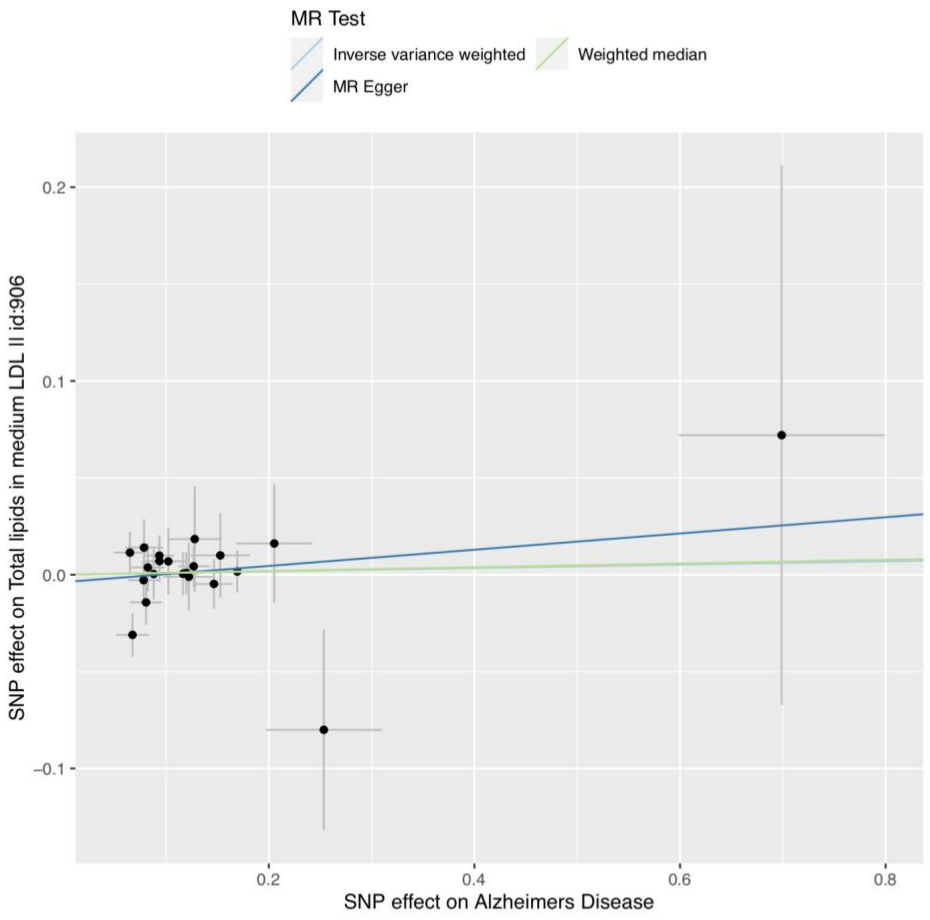
MR Test
Inverse variance weighted
Weighted median
MR Egger
SNP effect on Total lipids in medium LDL II id:906
SNP effect on Alzheimers Disease
0.2
0.1
0.0
−0.1
0.2
0.4
0.6
0.8

**S5u**

**S5v**

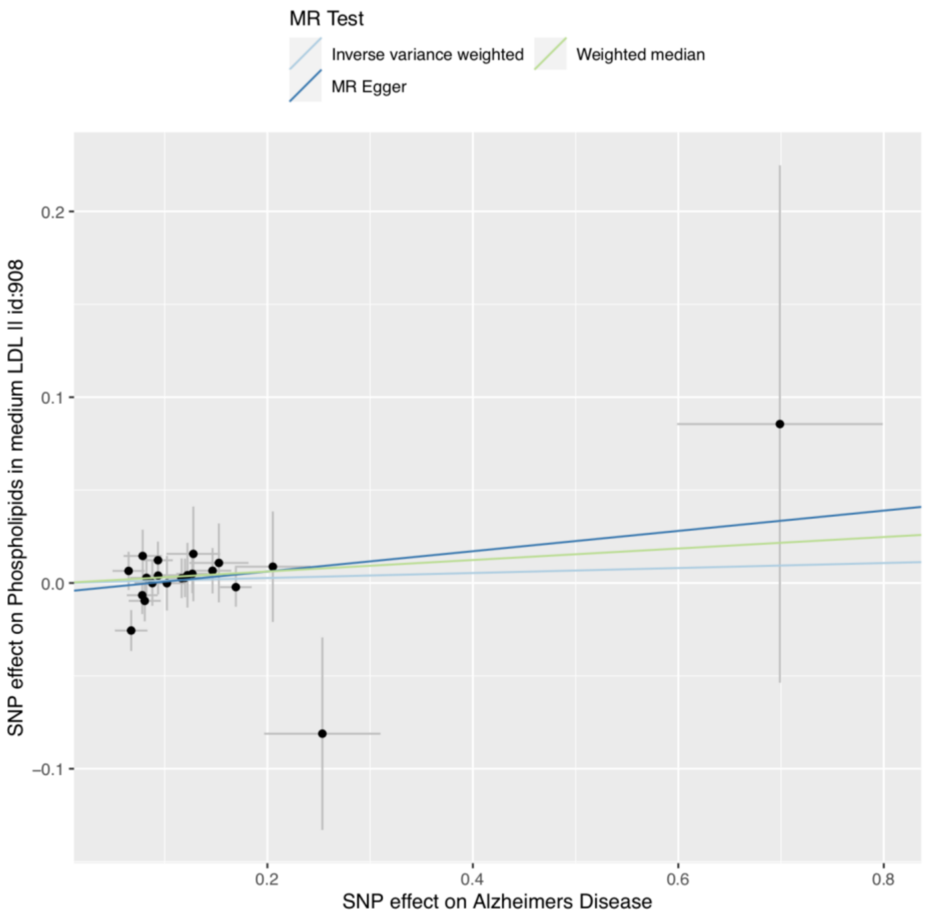
MR Test
Inverse variance weighted
Weighted median
MR Egger
SNP effect on Phospholipids in medium LDL II id:908
SNP effect on Alzheimers Disease
0.2
0.1
0.0
−0.1
0.2
0.4
0.6
0.8

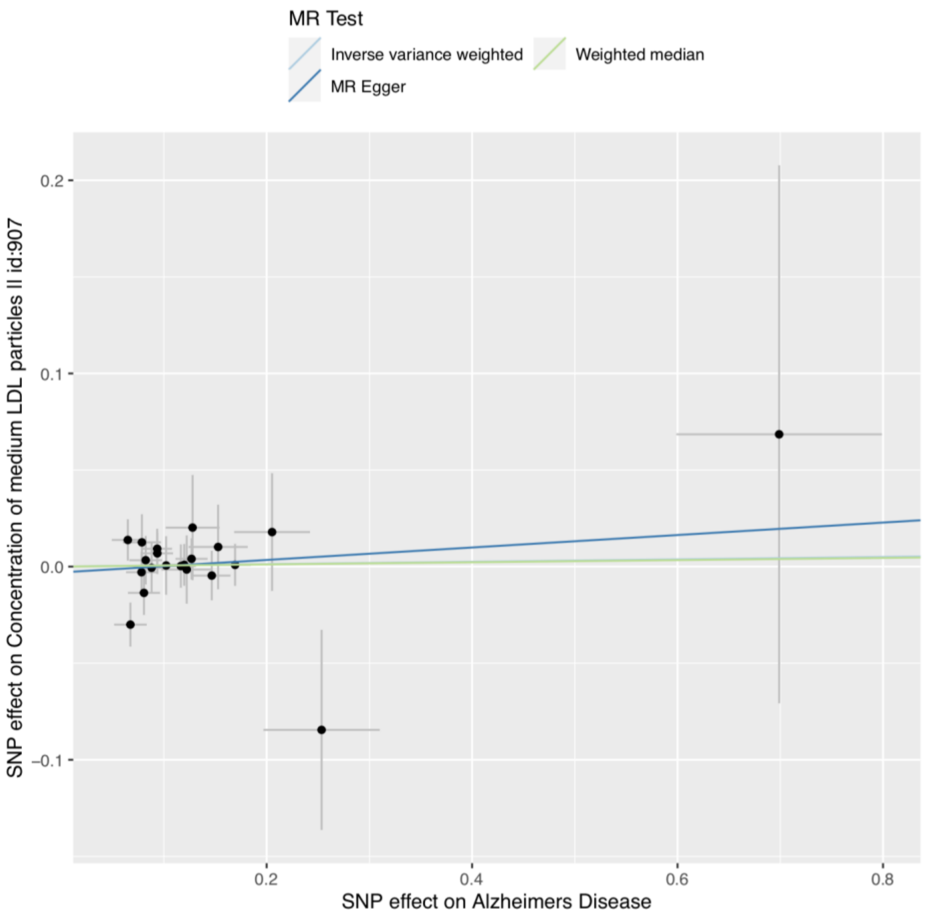
MR Test
Inverse variance weighted
Weighted median
MR Egger
SNP effect on Concentration of medium LDL particles II id:907
SNP effect on Alzheimers Disease
0.2
0.1
0.0
−0.1
0.2
0.4
0.6
0.8

**S5w**

**S5x**

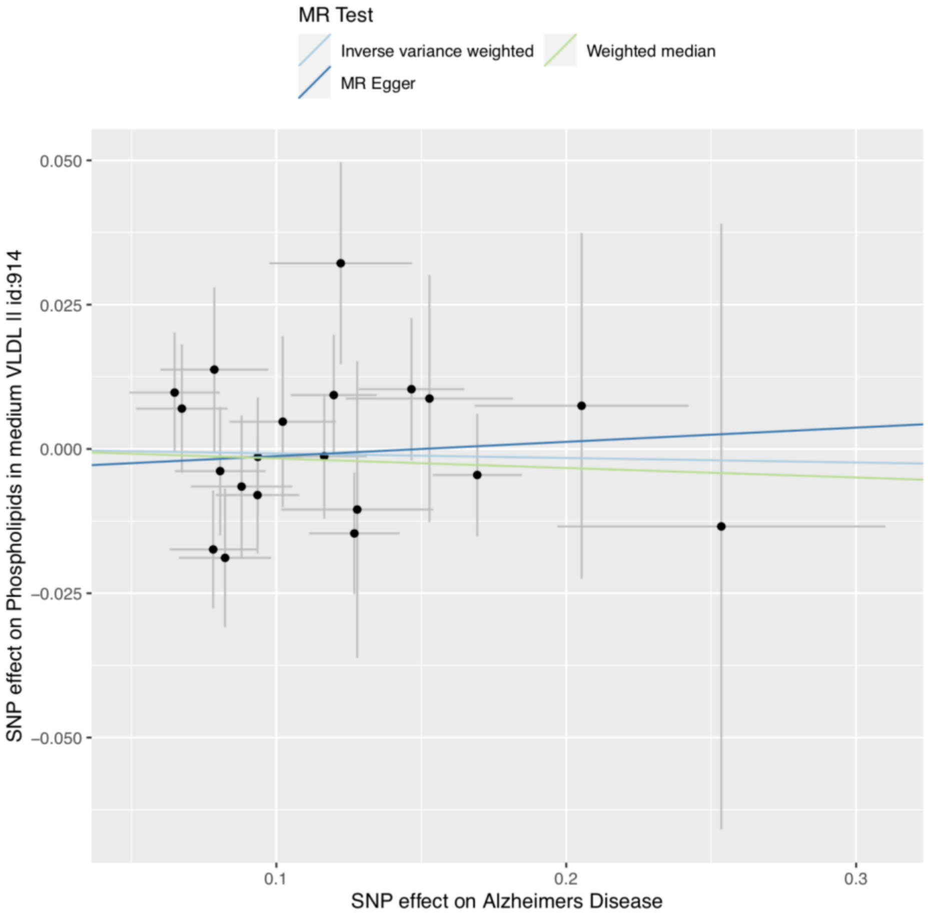

MR Test
Inverse variance weighted
Weighted median
MR Egger
0.050
0.025
0.000
−0.025
−0.050
SNP effect on Phospholipids in medium VLDL II id:914
0.1
0.2
0.3
SNP effect on Alzheimers Disease


**S5y**

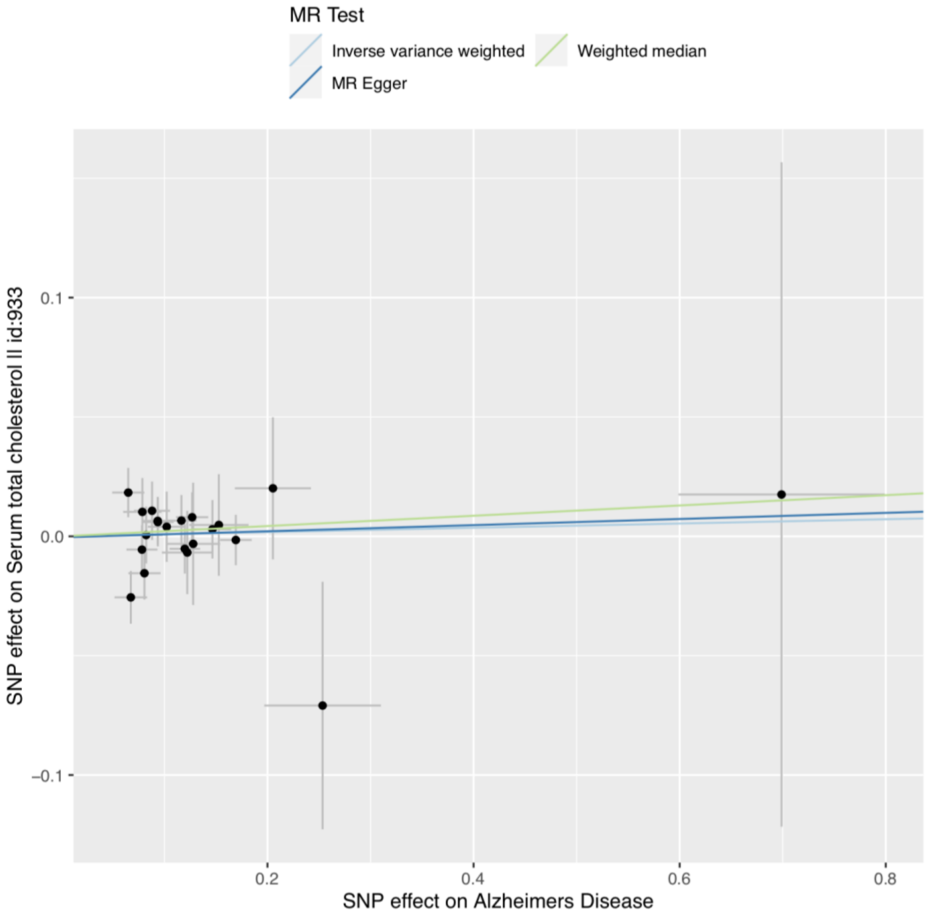

MR Test
Inverse variance weighted
Weighted median
MR Egger
0.1
0.0
−0.1
SNP effect on Serum total cholesterol II id:933
0.2
0.4
0.6
0.8
SNP effect on Alzheimers Disease


**S5z**

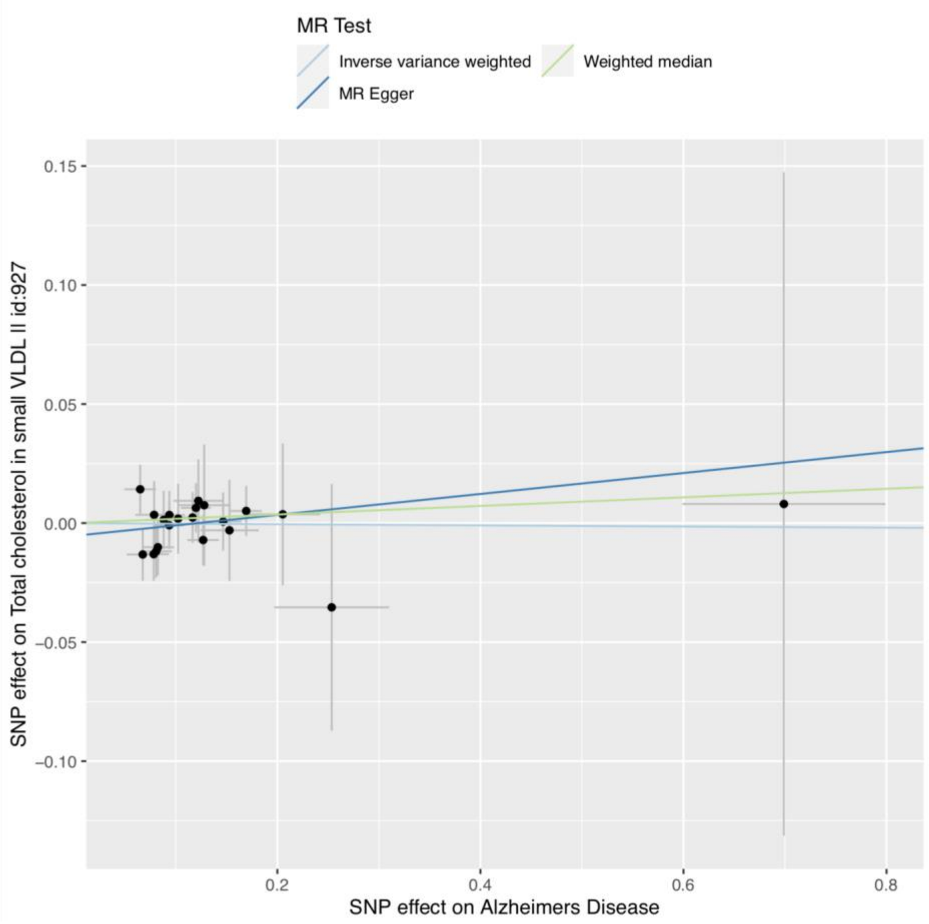
MR Test
Inverse variance weighted
Weighted median
MR Egger
0.15
0.10
0.05
0.00
−0.05
−0.10
SNP effect on Total cholesterol in small VLDL II id:927
0.2
0.4
0.6
0.8
SNP effect on Alzheimers Disease

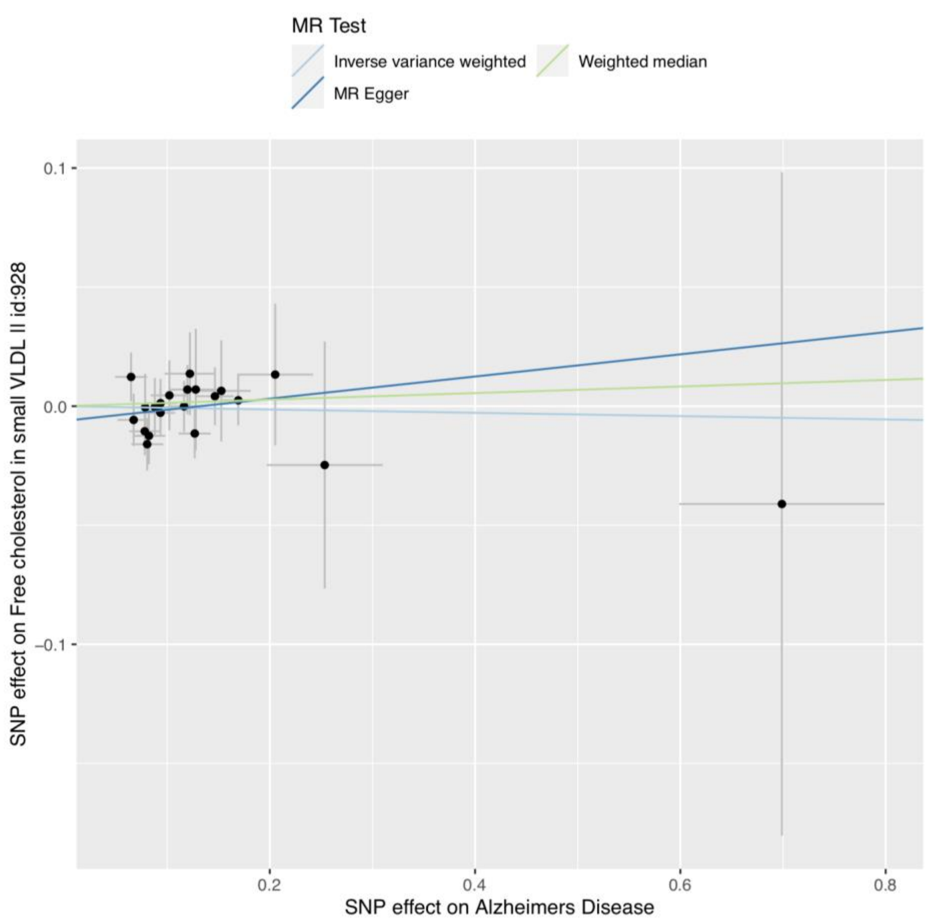
MR Test
Inverse variance weighted
Weighted median
MR Egger
0.1
0.0
−0.1
SNP effect on Free cholesterol in small VLDL II id:928
0.2
0.4
0.6
0.8
SNP effect on Alzheimers Disease

**S5aa** **S5ab**

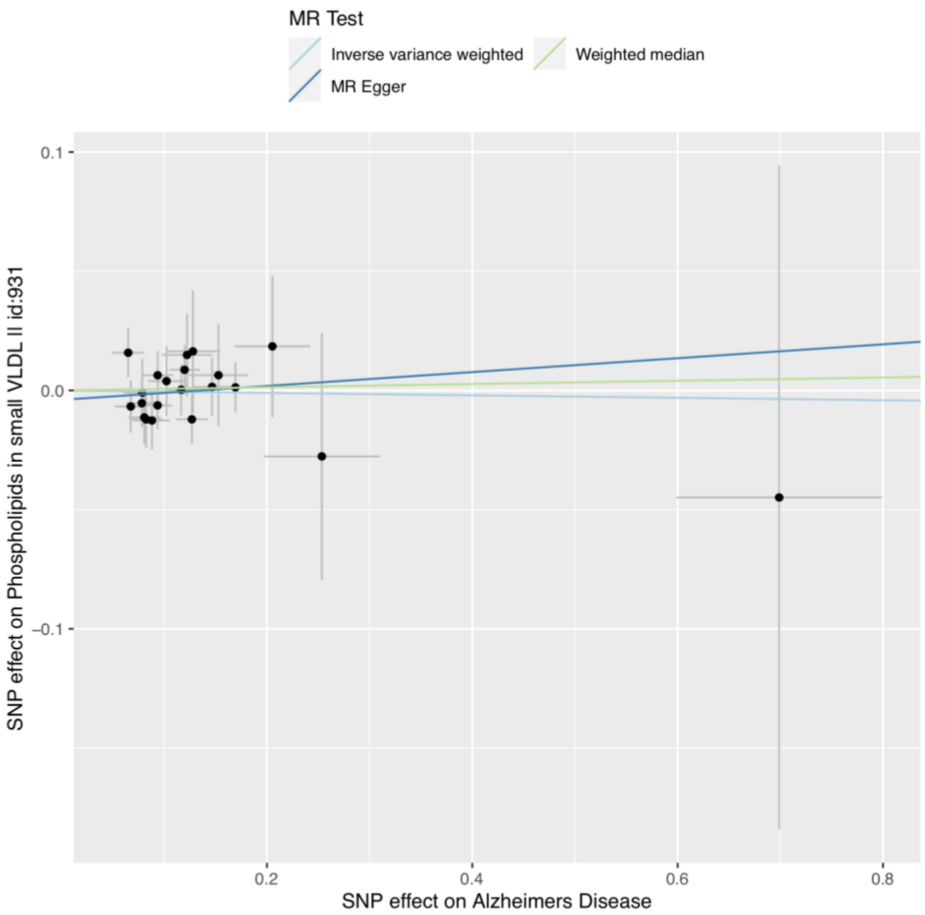
MR Test
Inverse variance weighted
Weighted median
MR Egger
0.1
0.0
−0.1
0.2
0.4
0.6
0.8
SNP effect on Phospholipids in small VLDL II id:931
SNP effect on Alzheimers Disease

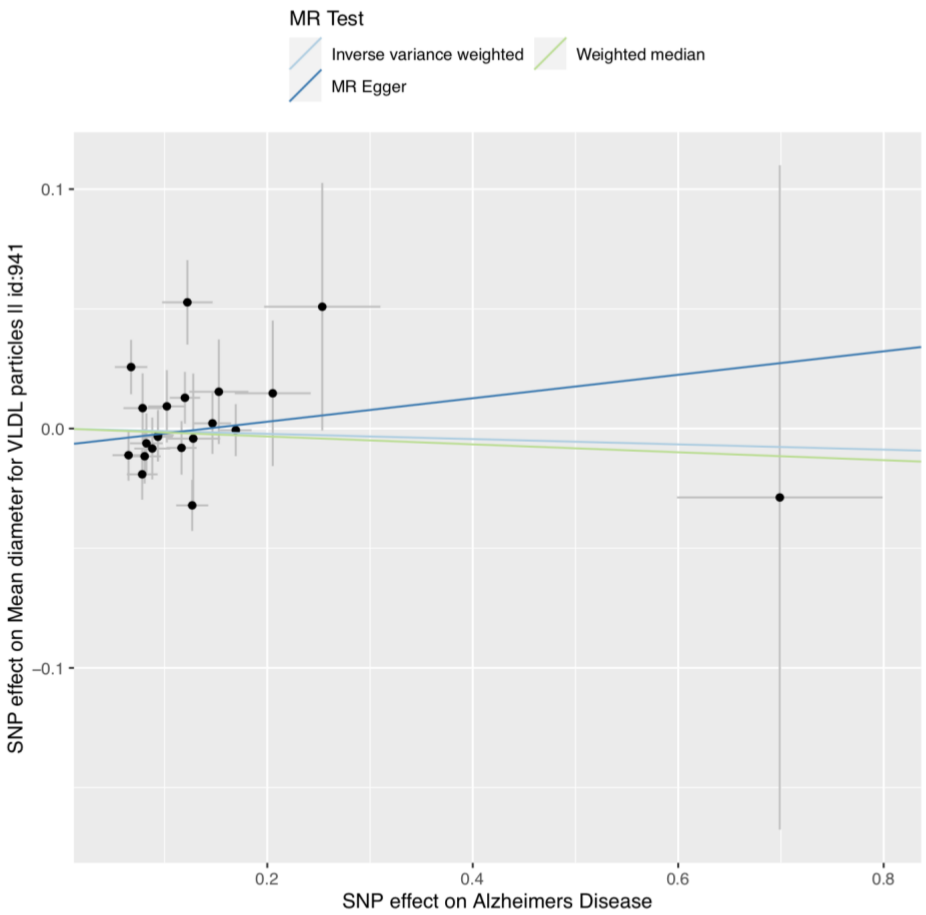
MR Test
Inverse variance weighted
Weighted median
MR Egger
0.1
0.0
−0.1
0.2
0.4
0.6
0.8
SNP effect on Mean diameter for VLDL particles II id:941
SNP effect on Alzheimers Disease

**S5ac**

**S5ad**

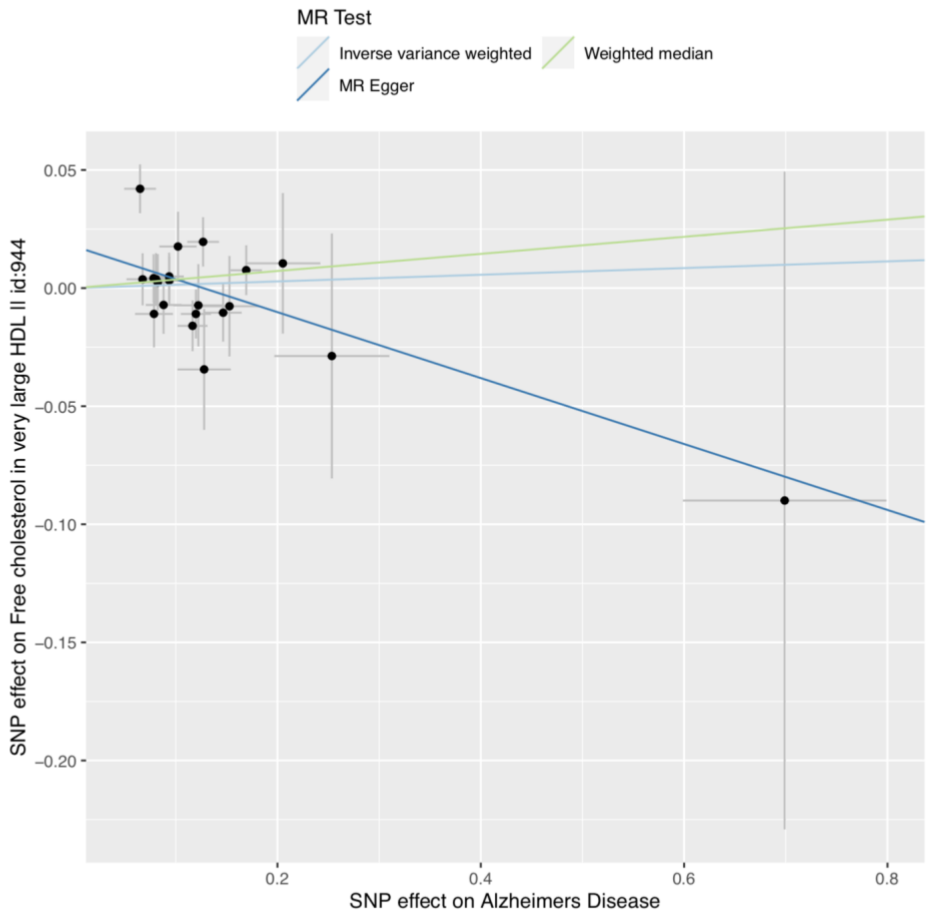
MR Test
Inverse variance weighted
Weighted median
MR Egger
SNP effect on Free cholesterol in very large HDL II id:944
0.05
0.00
−0.05
−0.10
−0.15
−0.20
0.2
0.4
0.6
0.8
SNP effect on Alzheimers Disease

**S5ae**

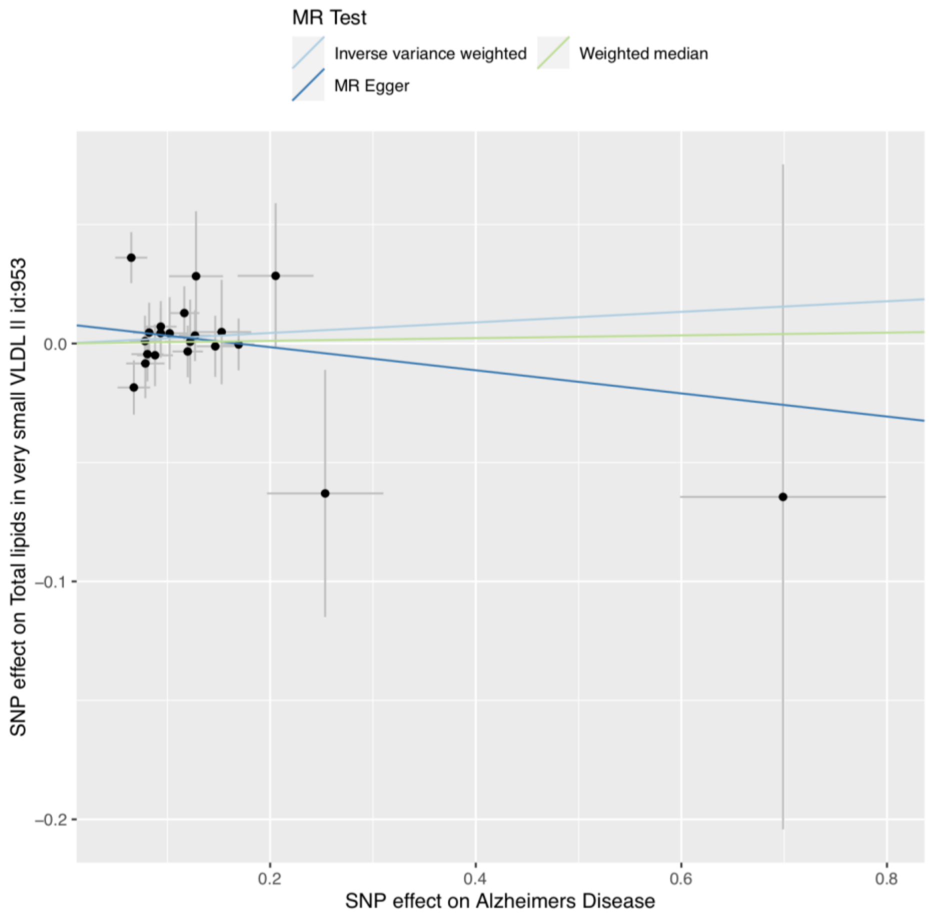
MR Test
Inverse variance weighted
Weighted median
MR Egger
SNP effect on Total lipids in very small VLDL II id:953
0.0
−0.1
−0.2
0.2
0.4
0.6
0.8
SNP effect on Alzheimers Disease

**S5af**

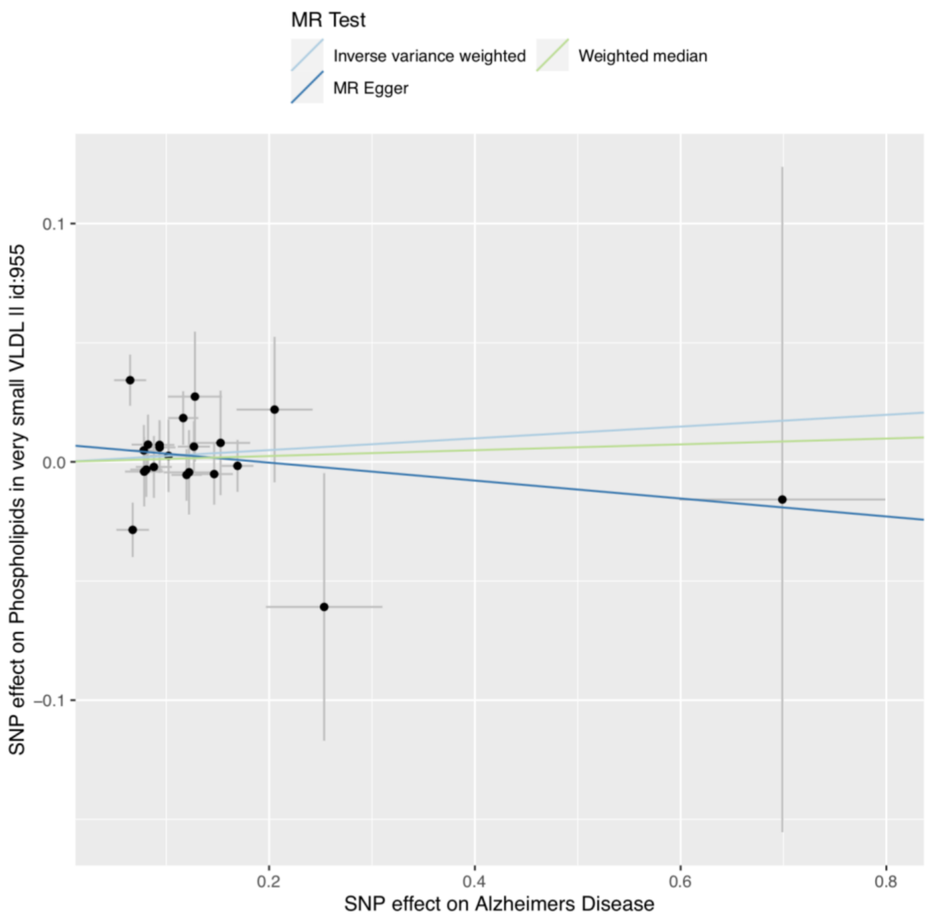
MR Test
Inverse variance weighted
Weighted median
MR Egger
SNP effect on Phospholipids in very small VLDL II id:955
0.1
0.0
−0.1
0.2
0.4
0.6
0.8
SNP effect on Alzheimers Disease

**S5ag**

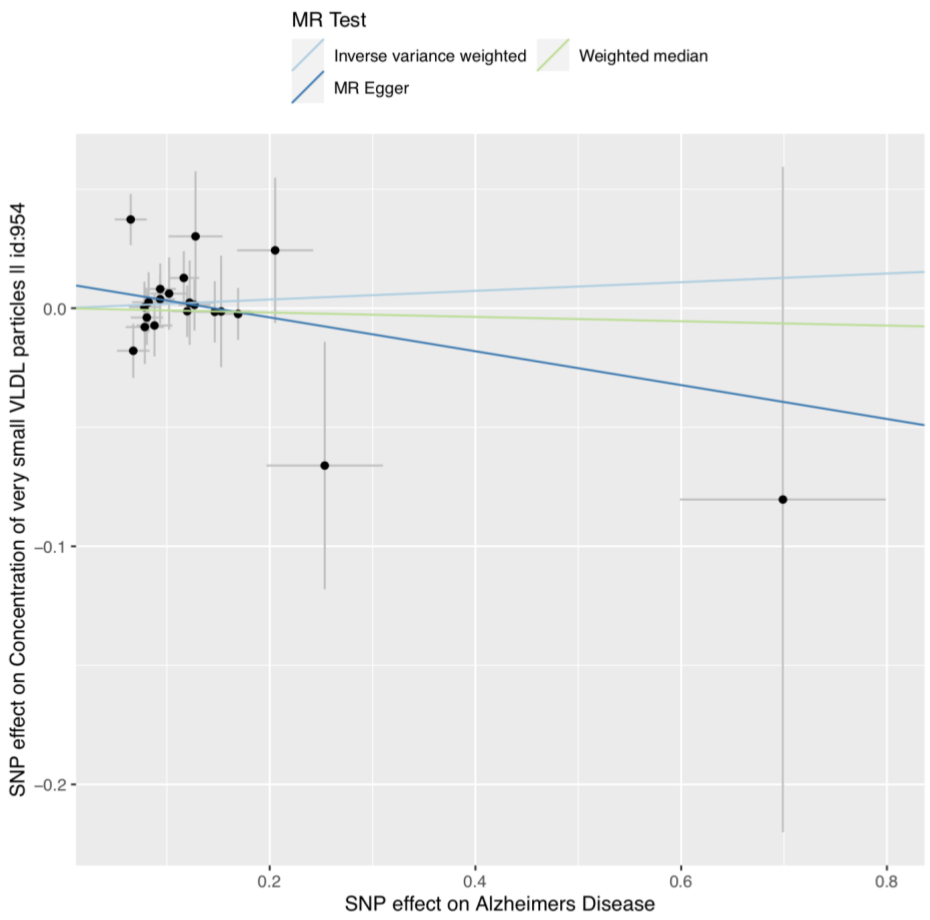
MR Test
Inverse variance weighted
Weighted median
MR Egger
SNP effect on Concentration of very small VLDL particles II id:954
0.0
−0.1
−0.2
0.2
0.4
0.6
0.8
SNP effect on Alzheimers Disease

**S5ah**

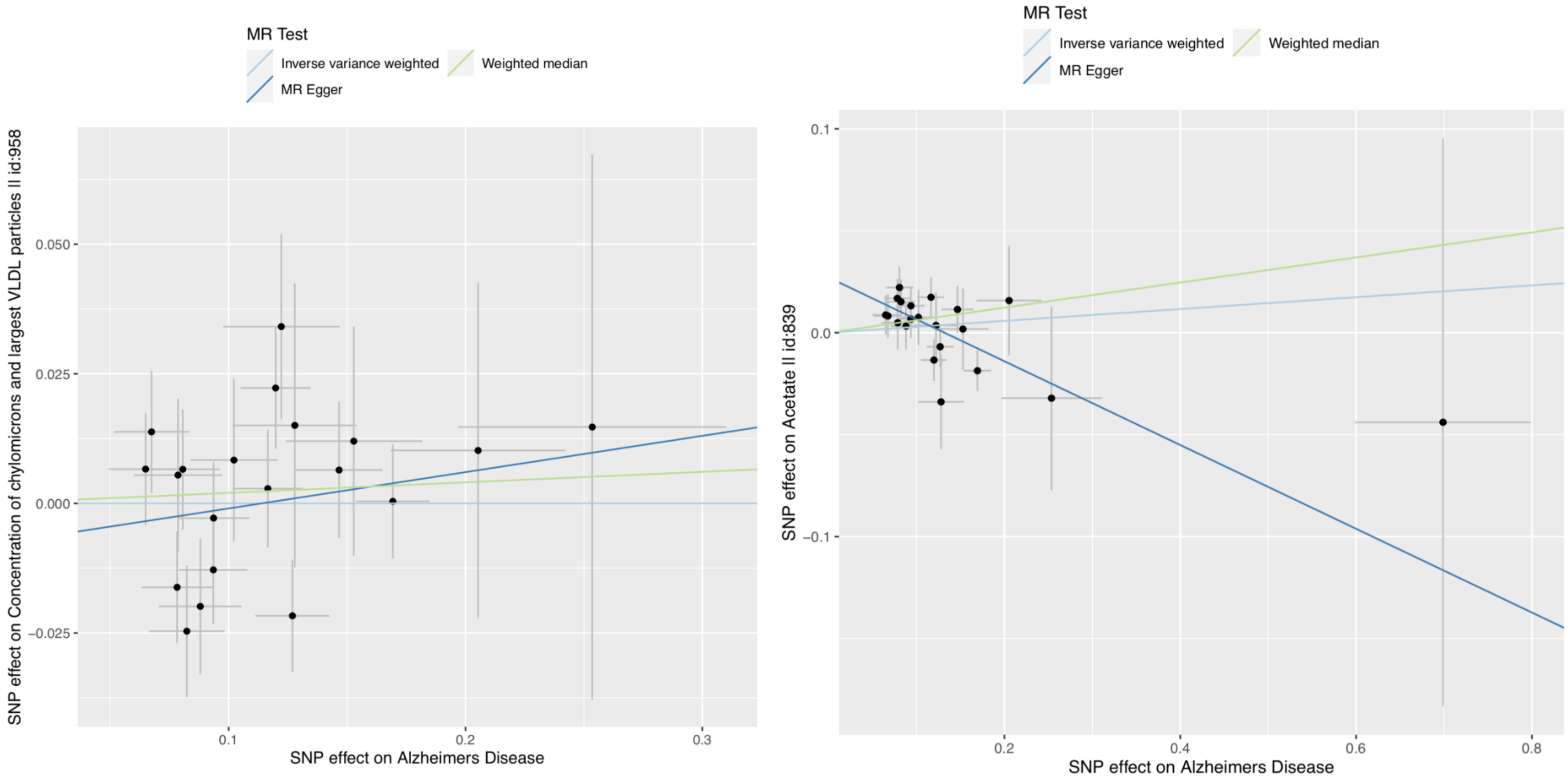


**Supplementary Figure S5(a-ah).** Scatter plot illustrating primary (IVW) and secondary (Egger and weighted median) causal slope estimates when AD is set as exposure and each metabolite set as outcome.

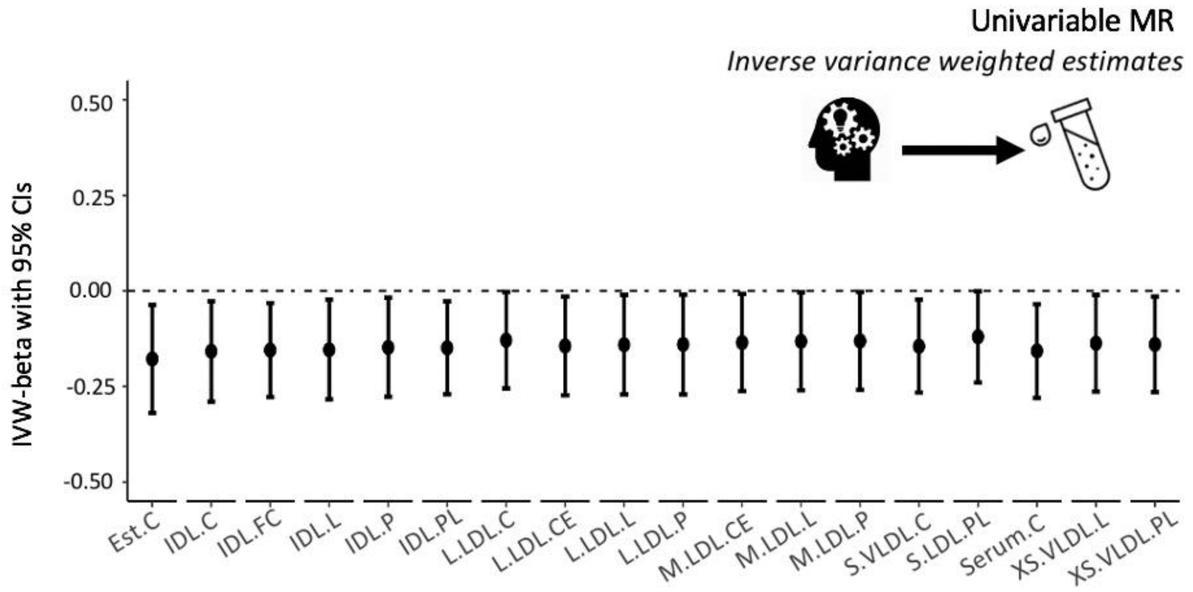


**Supplementary Figure S6.** Univariable effects of cognition on 18 lipid-related metabolites, significant at *p*<0.05.

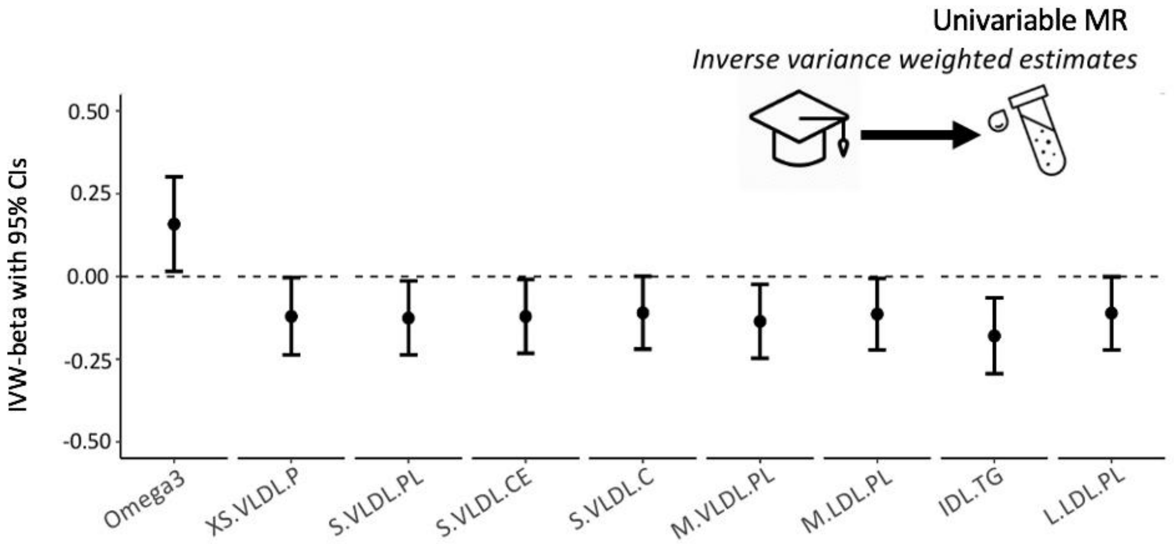


**Supplementary Figure S7.** Univariable effects of education nine metabolites, significant at *p*<0.05.

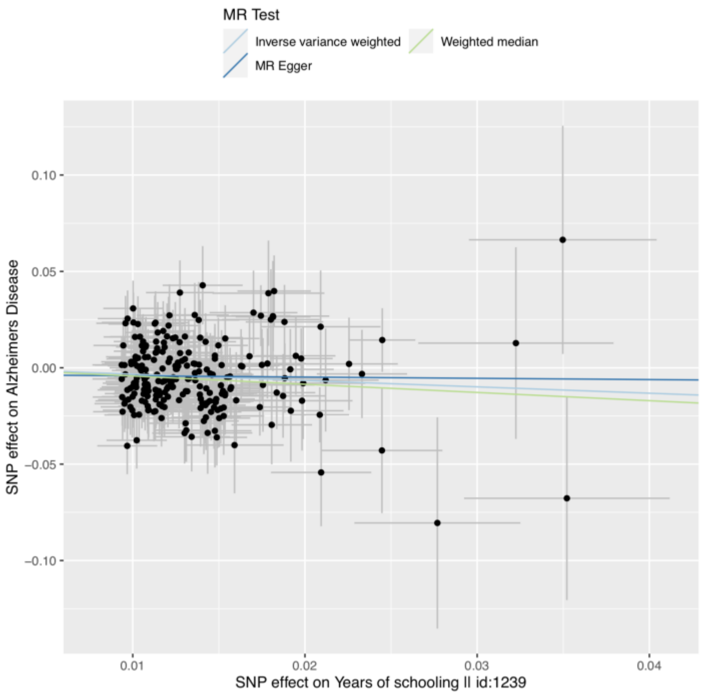


**Supplementary Figure S8.** Scatter plot illustrating primary (IVW) and secondary (Egger and weighted median) causal slope estimates when education set as exposure and AD set as outcome.

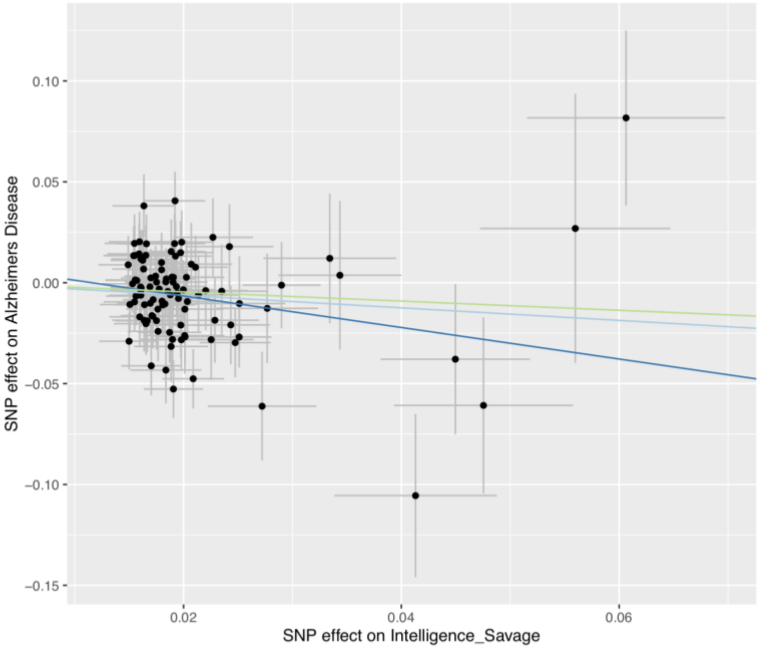


**Supplementary Figure S9.** Scatter plot illustrating primary (IVW) and secondary (Egger and weighted median) causal slope estimates when cognition set as exposure and AD set as outcome.

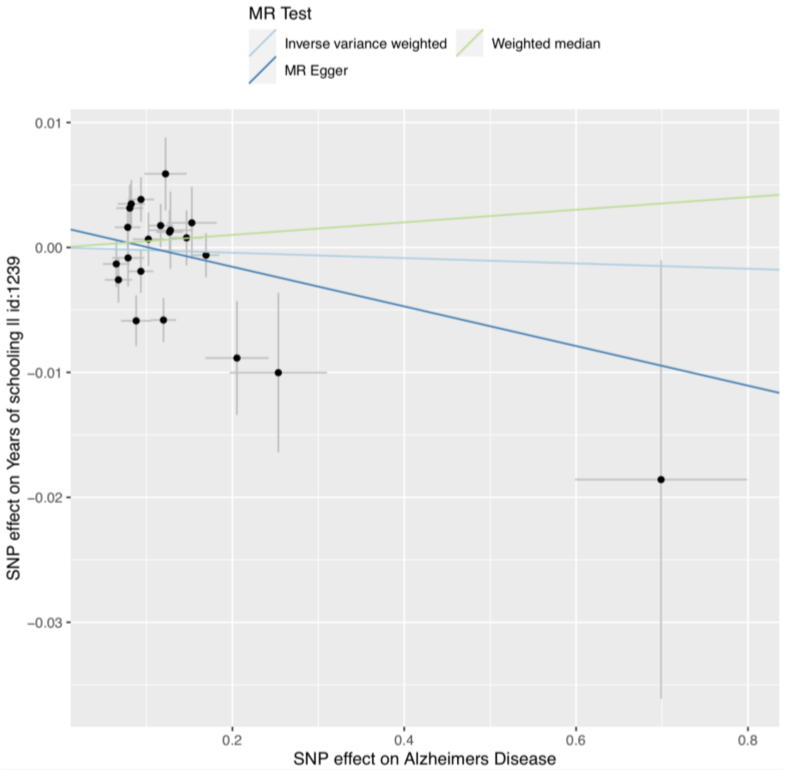


**Supplementary Figure S10.** Scatter plot illustrating primary (IVW) and secondary (Egger and weighted median) causal slope estimates when AD set as exposure and education as outcome.

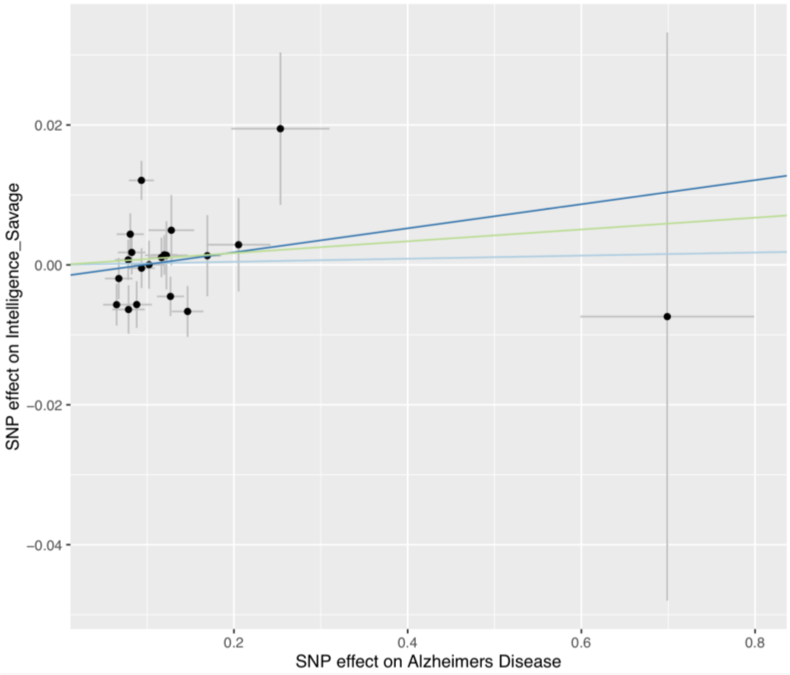


**Supplementary Figure S11.** Scatter plot illustrating primary (IVW) and secondary (Egger and weighted median) causal slope estimates when AD set as exposure and cognition as outcome.

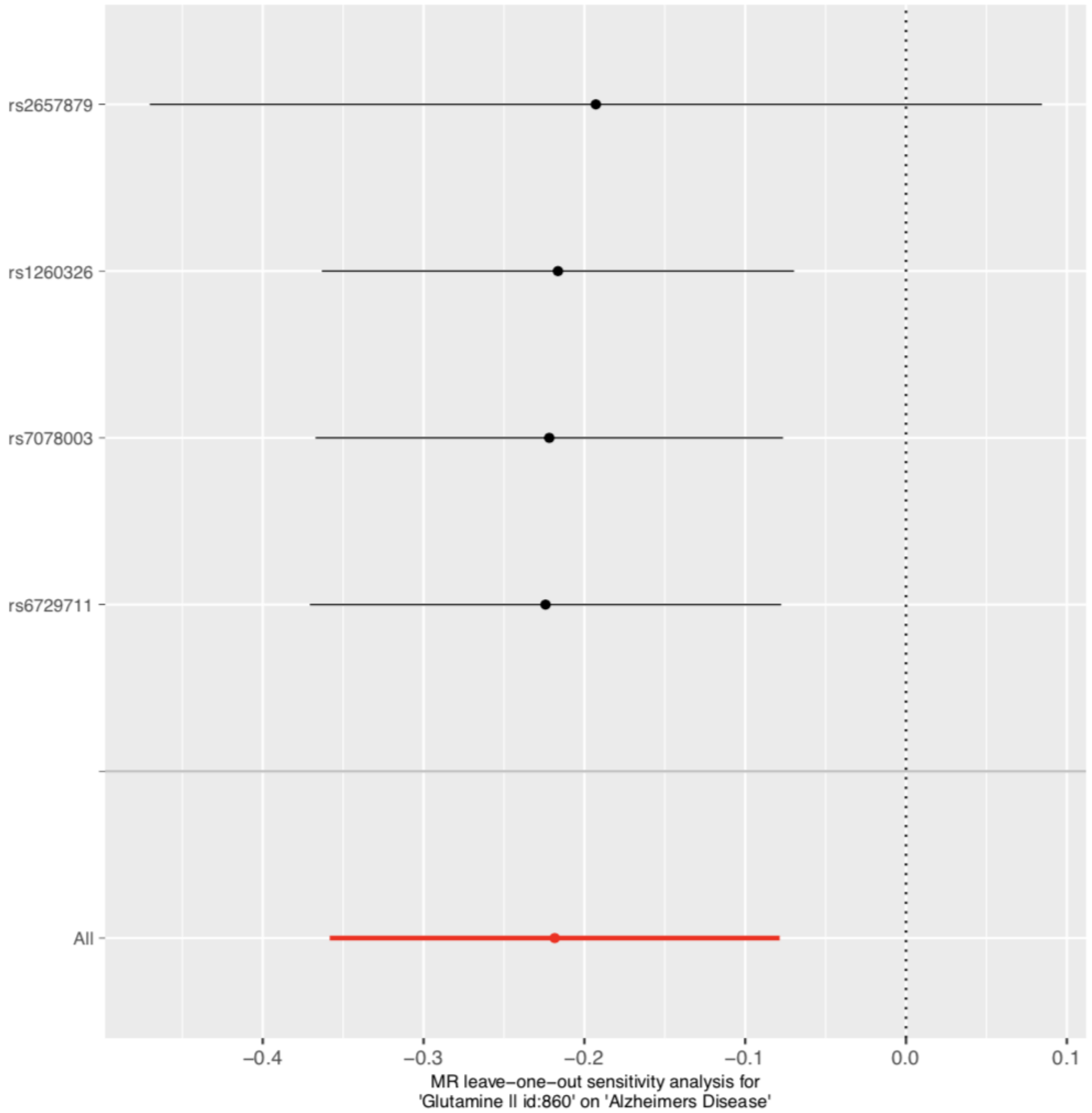


**Supplementary Figure S12.** Forest plot for leave-one-out analyses when Glutamine set as exposure and AD as outcome. The plot illustrates that removal of rs2657879 results in causal estimates crossing the null (vertical dotted line).

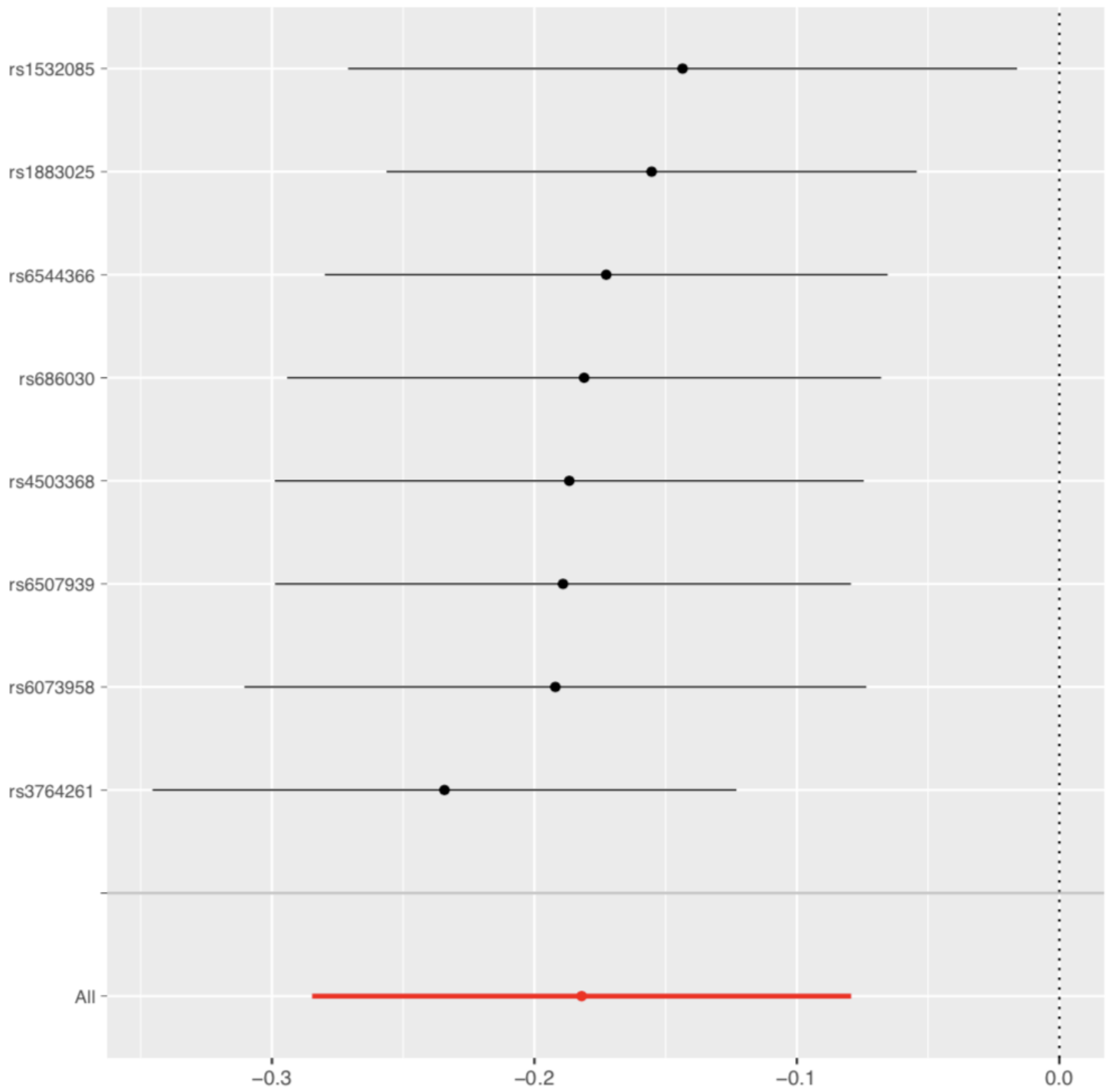


**Supplementary Figure S13.** Forest plot for leave-one-out analyses when XL.HDL.FC set as exposure and AD as outcome. The plot illustrates that no single-SNP removal results in estimates crossing the null. The greatest attenuation is observed with SNP rs1532085 (top).

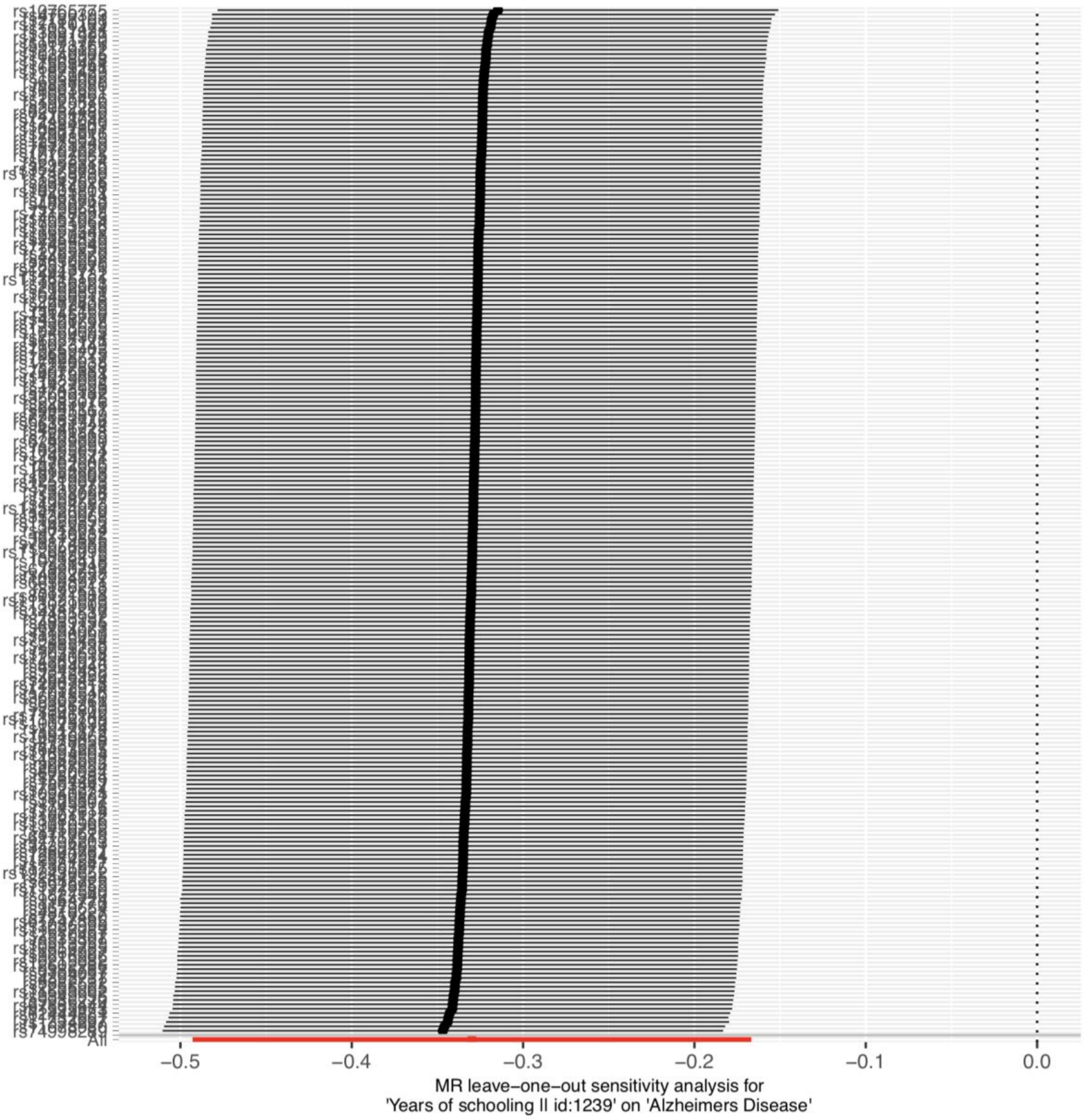


**Supplementary Figure S14.** Forest plot for leave-one-out analyses when education set as exposure and AD as outcome.

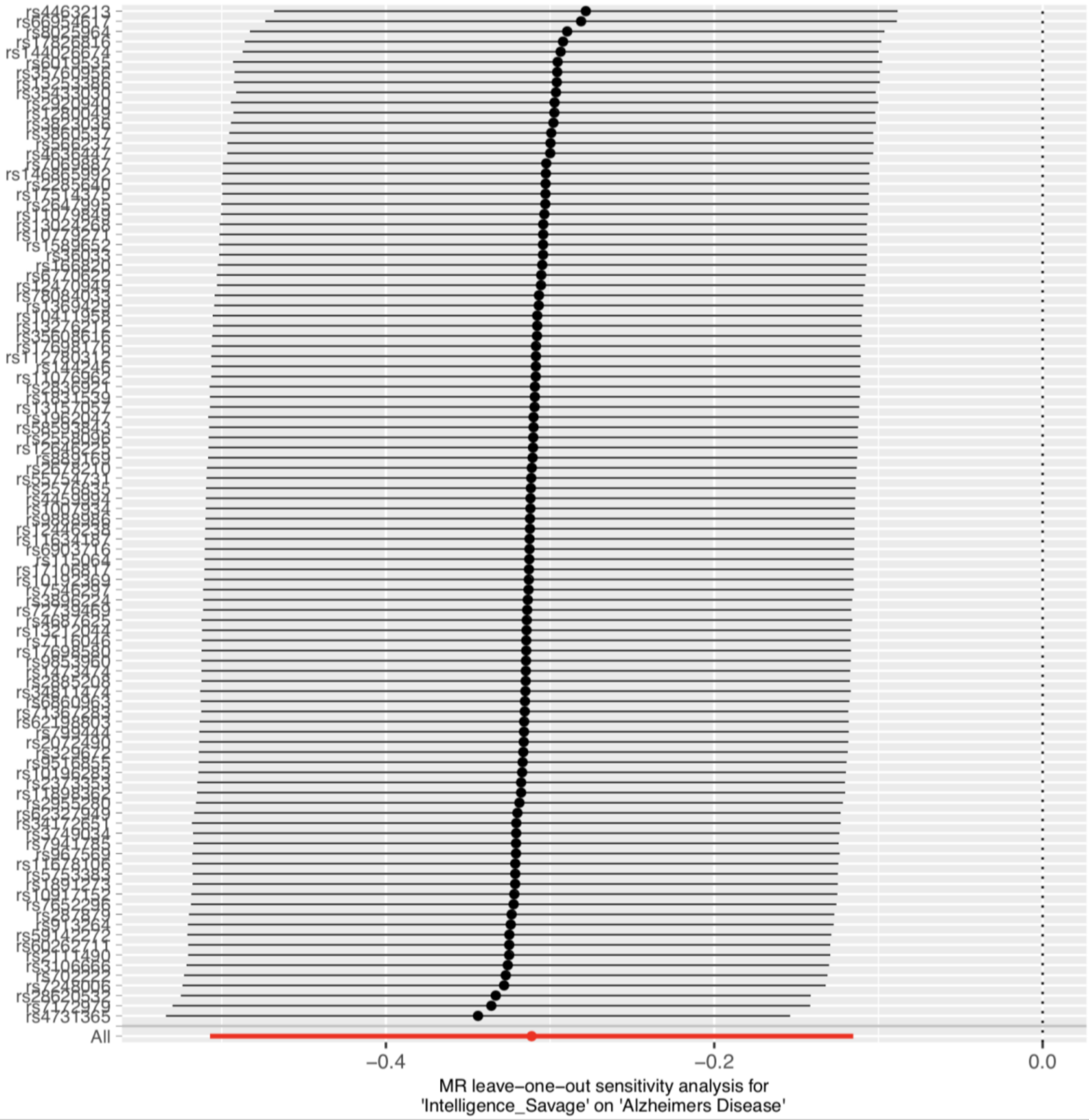


**Supplementary Figure S15.** Forest plot for leave-one-out analyses when cognition set as exposure and AD as outcome.

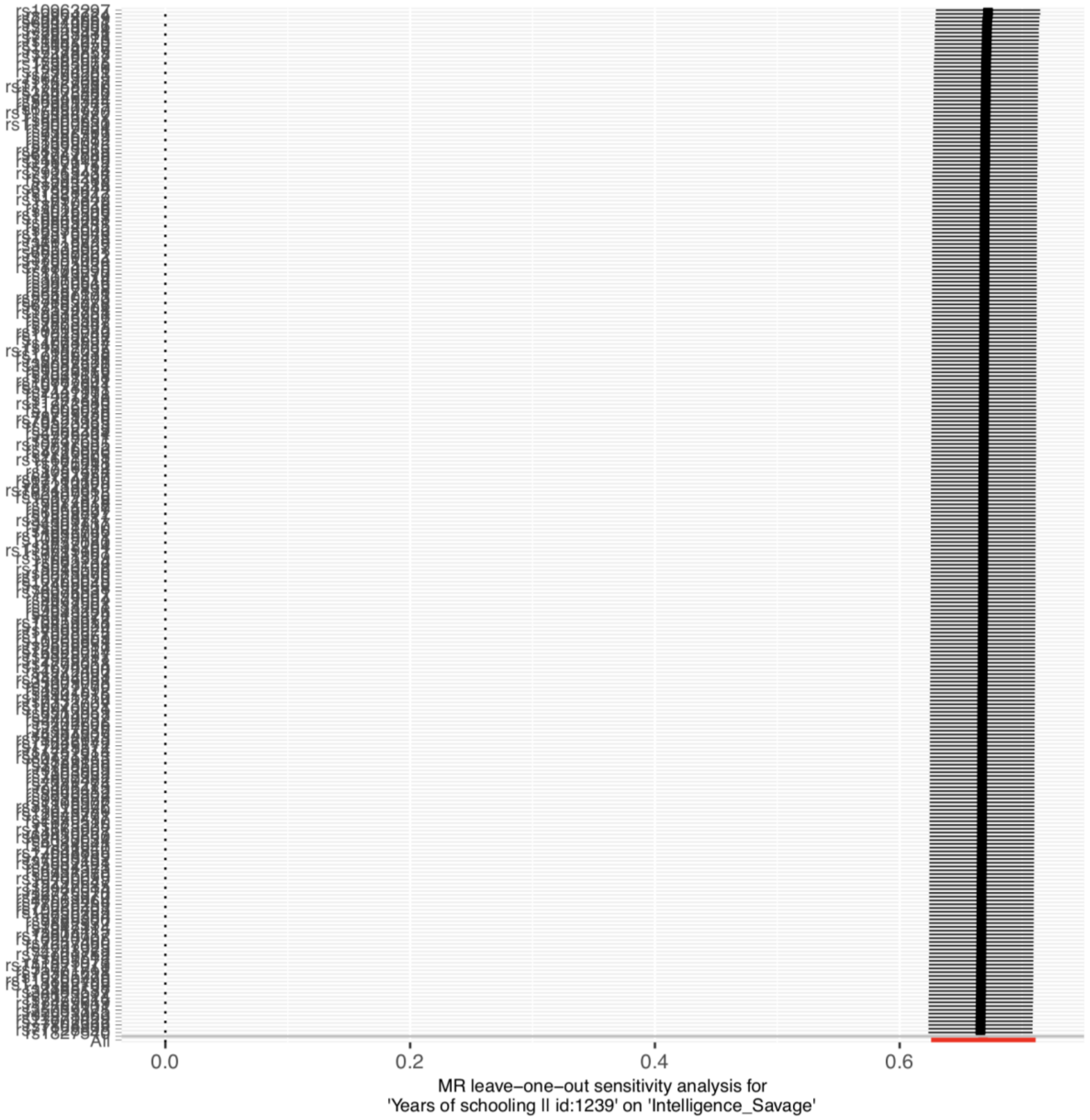


**Supplementary Figure S16.** Forest plot for leave-one-out analyses when education set as exposure and cognition as outcome.

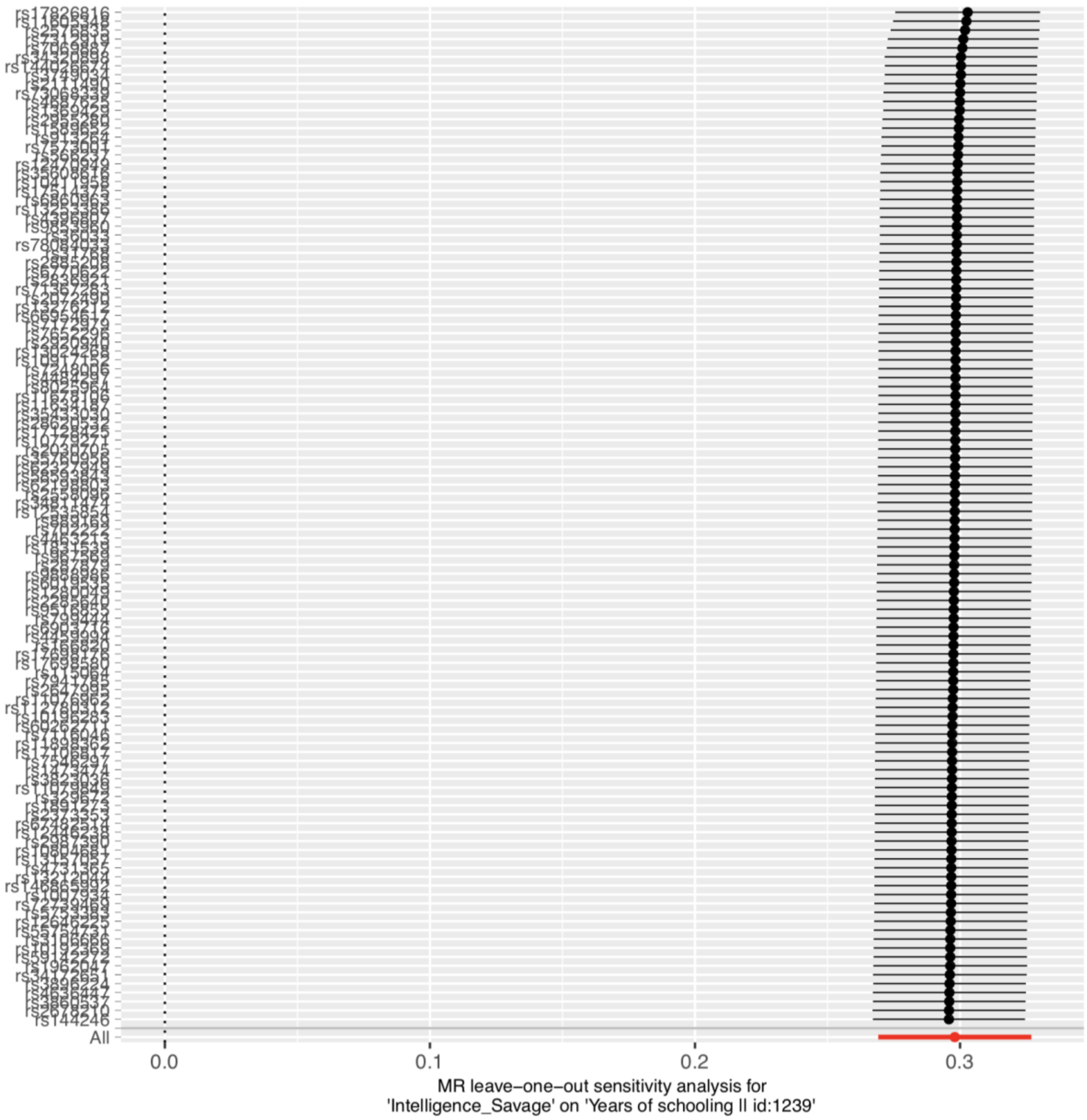


**Supplementary Figure S16.** Forest plot for leave-one-out analyses when cognition set as exposure and education as outcome.

### References

1. Burgess S, Davey Smith G, Davies NM, Dudbridge F, Gill D, Glymour MM, et al. Guidelines for performing Mendelian randomization investigations. Wellcome Open Res. 2019;4:186.

2. Kettunen J, Demirkan A, Würtz P, Draisma HHM, Haller T, Rawal R, et al. Genome-wide study for circulating metabolites identifies 62 loci and reveals novel systemic effects of LPA. Nat Commun. 2016 Mar 23;7:11122.

3. Kunkle BW, Grenier-Boley B, Sims R, Bis JC, Damotte V, Naj AC, et al. Genetic meta-analysis of diagnosed Alzheimer's disease identifies new risk loci and implicates Aβ, tau, immunity and lipid processing. Nat Genet. 2019 Mar;51(3):414–30.

4. Lee JJ, Wedow R, Okbay A, Kong E, Maghzian O, Zacher M, et al. Gene discovery and polygenic prediction from a 1.1-million-person GWAS of education. Nat Genet. 2018 Aug;50(8):1112–21.

5. Savage JE, Jansen PR, Stringer S, Watanabe K, Bryois J, de Leeuw CA, et al. Genome-wide association meta-analysis in 269,867 individuals identifies new genetic and functional links to cognition. Nat Genet. 2018 Jul;50(7):912–9.

6. Lord J, Jermy B, Green R, Wong A, Xu J, Legido-Quigley C, et al. Deciphering the causal relationship between blood metabolites and Alzheimers Disease: a Mendelian Randomization study. medRxiv. 2020 Apr 30;2020.04.28.20083253.

7. Bulik-Sullivan B, Finucane HK, Anttila V, Gusev A, Day FR, Loh P-R, et al. An Atlas of Genetic Correlations across Human Diseases and Traits. Nat Genet. 2015 Nov;47(11):1236–41.

8. Sattlecker M, Kiddle SJ, Newhouse S, Proitsi P, Nelson S, Williams S, et al. Alzheimer's disease biomarker discovery using SOMAscan multiplexed protein technology. Alzheimers Dement J Alzheimers Assoc. 2014 Nov;10(6):724–34.

9. Bowden J, Davey Smith G, Burgess S. Mendelian randomization with invalid instruments: effect estimation and bias detection through Egger regression. Int J Epidemiol. 2015 Apr;44(2):512–25.

10. Bowden J, Davey Smith G, Haycock PC, Burgess S. Consistent Estimation in Mendelian Randomization with Some Invalid Instruments Using a Weighted Median Estimator. Genet Epidemiol. 2016 May;40(4):304–14.

11. Zuber V, Colijn JM, Klaver C, Burgess S. Selecting likely causal risk factors from high-throughput experiments using multivariable Mendelian randomization. Nat Commun. 2020 Jan 7;11(1):29.

12. Wilson PW, Garrison RJ, Castelli WP, Feinleib M, McNamara PM, Kannel WB. Prevalence of coronary heart disease in the framingham offspring study: Role of lipoprotein cholesterols. Am J Cardiol. 1980 Oct 1;46(4):649–54.

13. Ouimet Mireille, Barrett Tessa J., Fisher Edward A. HDL and Reverse Cholesterol Transport. Circ Res. 2019 May 10;124(10):1505–18.

14. Bardagjy AS, Steinberg FM. Relationship Between HDL Functional Characteristics and Cardiovascular Health and Potential Impact of Dietary Patterns: A Narrative Review. Nutrients. 2019 Jun;11(6):1231.

15. Button EB, Robert J, Caffrey TM, Fan J, Zhao W, Wellington CL. HDL from an Alzheimer's disease perspective. Curr Opin Lipidol. 2019 Jun;30(3):224–34.

16. Zhang Q, Sidorenko J, Couvy-Duchesne B, Marioni RE, Wright MJ, Goate AM, et al. Risk prediction of late-onset Alzheimer's disease implies an oligogenic architecture. Nat Commun. 2020 23;11(1):4799.

17. Group TNS. Effect of a 12-mo micronutrient intervention on learning and memory in well-nourished and marginally nourished school-aged children: 2 parallel, randomized, placebo-controlled studies in Australia and Indonesia. Am J Clin Nutr. 2007 Oct 1;86(4):1082–93.

18. Robinson JG, Ijioma N, Harris W. Omega-3 fatty acids and cognitive function in women. Womens Health Lond Engl. 2010 Jan;6(1):119–34.

19. Benefits of omega-3 supplementation for schoolchildren: review of the current evidence on JSTOR [Internet]. [cited 2020 Nov 8]. Available from: https://www.jstor.org/stable/27896109?seq=1

20. Taylor R, Connock M. Effects of oily fish/omega-3 fatty acids on the behavioural, cogitative and educational outcomes of normal school children: A systematic review. :41.

21. Calder PC. Omega-3 fatty acids and inflammatory processes: from molecules to man. Biochem Soc Trans. 2017 Oct 15;45(5):1105–15.

22. Bäck M, Hansson GK. Omega-3 fatty acids, cardiovascular risk, and the resolution of inflammation. FASEB J Off Publ Fed Am Soc Exp Biol. 2019;33(2):1536–9.

23. Kiecolt-Glaser JK, Belury MA, Andridge R, Malarkey WB, Glaser R. Omega-3 supplementation lowers inflammation and anxiety in medical students: a randomized controlled trial. Brain Behav Immun. 2011 Nov;25(8):1725–34.

24. Wei MY, Jacobson TA. Effects of eicosapentaenoic acid versus docosahexaenoic acid on serum lipids: a systematic review and meta-analysis. Curr Atheroscler Rep. 2011 Dec;13(6):474–83.

25. Bradberry JC, Hilleman DE. Overview of Omega-3 Fatty Acid Therapies. Pharm Ther. 2013 Nov;38(11):681–91.

26. Skulas-Ray AC, Wilson PWF, Harris WS, Brinton EA, Kris-Etherton PM, Richter CK, et al. Omega-3 Fatty Acids for the Management of Hypertriglyceridemia: A Science Advisory From the American Heart Association. Circulation. 2019 17;140(12):e673–91.

27. Arca M, Borghi C, Pontremoli R, De Ferrari GM, Colivicchi F, Desideri G, et al. Hypertriglyceridemia and omega-3 fatty acids: Their often overlooked role in cardiovascular disease prevention. Nutr Metab Cardiovasc Dis NMCD. 2018;28(3):197–205.

28. Backes J, Anzalone D, Hilleman D, Catini J. The clinical relevance of omega-3 fatty acids in the management of hypertriglyceridemia. Lipids Health Dis. 2016 Jul 22;15(1):118.

29. Tada H, Nohara A, Inazu A, Mabuchi H, Kawashiri M-A. Remnant lipoproteins and atherosclerotic cardiovascular disease. Clin Chim Acta Int J Clin Chem. 2019 Mar;490:1–5.

30. Valdes-Marquez E, Parish S, Clarke R, Stari T, Worrall BB, METASTROKE Consortium of the ISGC, et al. Relative effects of LDL-C on ischemic stroke and coronary disease: A Mendelian randomization study. Neurology. 2019 12;92(11):e1176–87.

31. Imamura T, Doi Y, Arima H, Yonemoto K, Hata J, Kubo M, et al. LDL cholesterol and the development of stroke subtypes and coronary heart disease in a general Japanese population: the Hisayama study. Stroke. 2009 Feb;40(2):382–8.

32. Zeljkovic A, Vekic J, Spasojevic-Kalimanovska V, Jelic-Ivanovic Z, Bogavac-Stanojevic N, Gulan B, et al. LDL and HDL subclasses in acute ischemic stroke: prediction of risk and short-term mortality. Atherosclerosis. 2010 Jun;210(2):548–54.

33. Proitsi P, Kuh D, Wong A, Maddock J, Bendayan R, Wulaningsih W, et al. Lifetime cognition and late midlife blood metabolites: findings from a British birth cohort. Transl Psychiatry. 2018 Sep 26;8(1):1–11.

34. Kovács KR, Bajkó Z, Szekeres CC, Csapó K, Oláh L, Magyar MT, et al. Elevated LDL-C combined with hypertension worsens subclinical vascular impairment and cognitive function. J Am Soc Hypertens JASH. 2014 Aug;8(8):550–60.

35. Gencer B, Mach F. So low… so far so good: neurocognitive impact of lowering LDL-C levels with PCSK9 inhibitors. Eur Heart J. 2018 01;39(5):382–4.

36. de Oliveira J, Hort MA, Moreira ELG, Glaser V, Ribeiro-do-Valle RM, Prediger RD, et al. Positive correlation between elevated plasma cholesterol levels and cognitive impairments in LDL receptor knockout mice: relevance of cortico-cerebral mitochondrial dysfunction and oxidative stress. Neuroscience. 2011 Dec 1;197:99–106.

37. Zanchetti A, Liu L, Mancia G, Parati G, Grassi G, Stramba-Badiale M, et al. Blood pressure and LDL-cholesterol targets for prevention of recurrent strokes and cognitive decline in the hypertensive patient: design of the European Society of Hypertension-Chinese Hypertension League Stroke in Hypertension Optimal Treatment randomized trial. J Hypertens. 2014 Sep;32(9):1888–97.

38. Stobart JL, Anderson CM. Multifunctional role of astrocytes as gatekeepers of neuronal energy supply. Front Cell Neurosci. 2013;7:38.
